# Interactions with polygenic background impact quantitative traits in the UK Biobank

**DOI:** 10.1101/2025.11.14.25340263

**Authors:** Lino A. F. Ferreira, Sile Hu, Robert W. Davies, Simon R. Myers

**Affiliations:** Centre for Human Genetics, University of Oxford; Department of Statistics, University of Oxford; Genomics Ltd

## Abstract

Association studies have linked many genetic variants to a variety of phenotypes but understanding the biological mechanisms underlying these signals remains a major challenge. Since genes operate within complex networks, statistical interactions between genetic mutations that reflect biological pathways are expected to exist. However, their discovery has been hampered by the vast search space of variant combinations and the multiplicatively small expected effect sizes of interactions. To increase power, we created a test for interaction between single-nucleotide polymorphisms (SNPs) and *groups* of other variants with a direct effect on a phenotype aggregated in a polygenic score (PGS) which can be performed for any quantitative trait. In realistic simulations, this method avoids false positives and is well powered to find interaction networks. We apply it to 97 quantitative phenotypes in European samples in the UK Biobank and identify 144 independent interactions affecting 52 different traits, including important disease risk variants at genes such as *APOE*, *FTO* or *TCF7L2*. We develop approaches to refine identified signals and detect 38 pairwise interactions between SNPs. These include known interactions between *ABO*, *FUT2* and *TREH* affecting alkaline phosphatase levels, which are shown to be part of a larger network including *PIGC* and *FUT6*, as well as an interaction for eosinophil levels between *IL33* and *ALOX15*, two genes whose functional interaction has recently been implicated in asthma. Finally, we propose a method to partition PGSs according to the binding sites of more than 1100 transcription factors using the HOCOMOCO motif database and test for interactions involving functionally partitioned scores. We identify 12 interactions affecting eight traits, two of which directly reflect known regulatory relationships such as that between *TCF7L2* (a key regulator of glucose metabolism) and the transcription factor *KDM2A*, which are known to interact functionally within the Wnt signalling pathway, affecting glycated haemoglobin levels. This work substantially extends the set of known epistatic effects for human phenotypes and shows how statistical interactions can reflect biological interdependencies between genes.

## Main

Genome-wide association studies (GWASs) have identified hundreds of thousands of associations between genetic and phenotypic variation across a wide range of phenotypes [1]. These have provided a better understanding of genetic architecture [2], improved knowledge of biological mechanisms [3] and made possible the genetic prediction of disease risk through polygenic scores (PGSs) [4].

The overwhelming majority of genetic associations discovered for human phenotypes involve the direct effects of single-nucleotide polymorphisms (SNPs) [5]. Because many of these variants likely operate within overlapping biological pathways relevant to these phenotypes, statistical interactions between these variants seem *a priori* likely to occur. However, efforts to find such interactions (where the combined effect of two or more variants is different from the sum of their effects when present in isolation, known as ‘epistasis’) have proved challenging [6, 7], with few robust effects so far identified [8–13]. Further, evidence suggests that the genetic architecture of most human phenotypes is well approximated by an additive model [14, 15], which implies that modelling epistasis is unlikely to improve PGS performance or account for ‘missing heritability’ [16]. One explanation for this is that, even if interactions are widespread, the small effect sizes of most associations imply that interaction terms, which can be modelled as multiplicative effects between variants, would be predicted to be tiny, so that power to detect interactions is low, and phenotypic variance would be captured to a large degree by additive estimates of direct genetic effects [17].

Nevertheless, identification of epistasis remains an important unsolved problem because of its potential to associate genetic findings with biological networks and so mechanisms. Examples of how identifying instances of epistasis affecting particular phenotypes can improve our understanding of the biological pathways that underlie them include the interaction between *ABO* and *FUT2* affecting alkaline phosphatase (ALP) levels, which reflects a known functional interdependence between ABH secretor status, ABO blood type and intestinal ALP [10, 18, 19], or that between an *ERAP1* variant and the *HLA-B27* allele for ankylosing spondylitis risk, informative about disease mechanism since the ERAP1 enzyme is involved in peptide trimming for HLA presentation [9]. Given the longstanding challenge of deciphering the mechanisms underlying GWAS associations, and the important role attributed to gene regulation [20] (which operates through functional interdependencies, e.g., between transcription factors (TFs) and their binding targets), finding and understanding interactions could help bridge the large gap that remains between statistical associations and their biological interpretation.

As well as small effect sizes, another significant challenge to the study of interactions is the vast search space: the number of possible pairwise interactions to test grows quadratically with the number of variants in a dataset, requiring substantial computational resources and stringent multiple-testing corrections [21]. Previous approaches to circumvent these obstacles include high-performance computing–based methods [22] (although these still suffer a loss of power due to multiple testing), two-stage approaches testing only a subset of potential pairwise interactions [23] and, more recently, methods that test for interactions between a variant and all other variants in the genome under a variance components framework (‘marginal epistasis’) [24–26]. Finally, a challenge in identifying interactions is the definition itself, because transformed phenotypes may behave non-linearly with genetic influences, technically generating interactions between all pairs of variants. Whether such interactions are ‘real’ is debated [27] and partly a question of definitions, but in any case they are less likely to lead to genuine biological understanding than effects that differ between SNPs impacting some trait, and in this work we focus on identifying such differing effects. Overall, the success of methods to find interactions has so far been limited, and the extent to which statistical interactions are present and able to shed light on biology therefore remains an open question.

Here we propose a new method that tests for interactions between individual genetic variants (primarily SNPs but also insertions and deletions) and a PGS that captures some fraction of the additive genetic effects underlying a phenotype. Beyond enabling the discovery of new interactions with only modest computational cost, and avoiding the usual multiple testing correction burden, the primary motivation for this method is to identify an underexplored type of interaction, i.e., that between a single genetic variant and *groups* of variants with a direct effect on a trait. Such an interaction could reflect, for example, the disruptive action on a pathway of a single mutation at a master regulator gene, and therefore yield important functional insight. We also develop approaches to refine identified signals to detect particular sets of interacting SNPs. We validate our method through extensive simulations and apply it to 97 quantitative phenotypes in the UK Biobank (UKB) [28], finding 144 independent interactions for 52 different traits. We then identify more localised signals driving these interactions: we detect 38 pairwise (SNP×SNP) interactions for 21 traits, the great majority of which are novel, and 12 interactions with components of the PGS defined according to TF binding sites affecting eight traits. This work substantially increases the set of statistical interactions (including of novel types) known to affect human phenotypes, provides further evidence of how interactions can closely reflect functional interdependencies such as gene regulatory processes, and suggests promising avenues for future research.

## Results

### Overview of methods

Our approach centres around a linear regression model of a quantitative phenotype on a SNP, a PGS and their interaction:

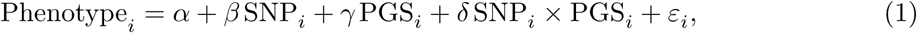

where *i* indexes individuals in the sample, SNP_*i*_ represents the count (or dosage) of minor alleles and ε_*i*_ is an error term. We assume that relevant covariates (especially age, sex and a measure of ancestry to control for population structure, for which we use Ancestry Components [29]) have been regressed out from the phenotype in advance, which simplifies model fitting in practice. While any PGS could in principle be used, for this application we developed a novel PGS construction algorithm that exhausts the additive signal for all SNPs that comprise it, aiming to increase power to detect non-additive effects (Methods).

Given a sample for which a phenotype of interest, a PGS and a set of genotypes are available, the model above can be fitted for each SNP one-by-one, similarly to the usual GWAS framework. We are interested in SNPs whose statistical interaction with the PGS is non-zero: the genotype at such a SNP can be thought of as modulating the effect of the SNPs included in the PGS or, perhaps more intuitively, the SNP’s effect on the trait is estimated to depend on where in the empirical distribution of the PGS an individual lies (Fig. 1a). In this model, the SNP of interest is fitted as interacting with all SNPs in the PGS with the same scaling factor. However, we note that we still expect some power to identify interactions between the SNP and only a subset of SNPs in the PGS (see later simulations), provided these are sufficiently strong, because these form part of the wider PGS. We note also that this test is sensitive only to gene–gene (rather than gene–environment) interactions, though these might be mediated rather indirectly, including via their impacts on other, potentially unmeasured, phenotypes.

**Fig. 1:**
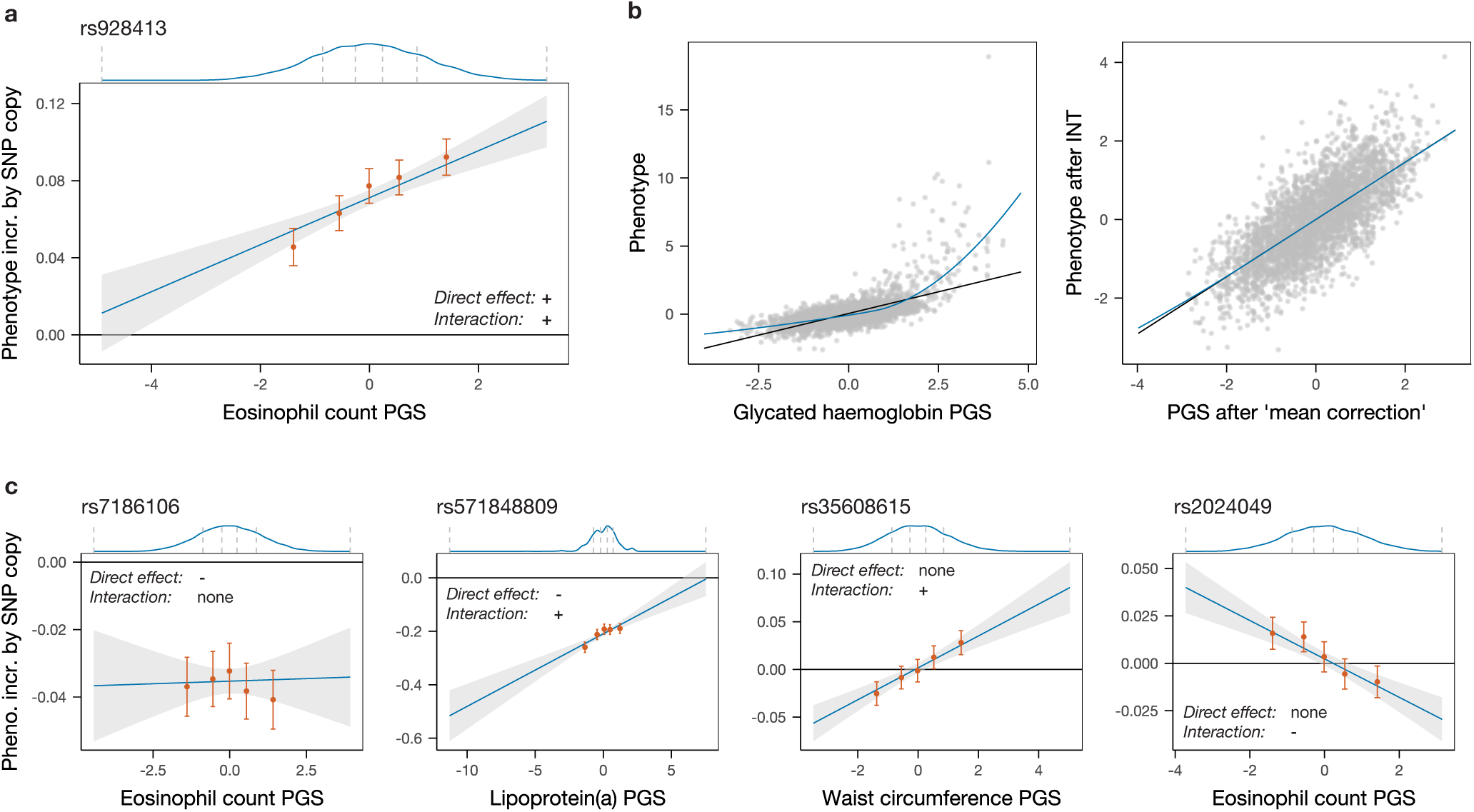
Examples of types of interaction captured and detail on how global non-linear relationships are modelled. **a** | Positive interaction between rs928413, a SNP with a positive direct effect on eosinophil count, and the PGS for this trait. This effect can be interpreted in two equivalent ways: the average effect of the SNP on the phenotype increases with the PGS; or the slope of the increasing line linking PGS with phenotype increases with each copy of the SNP’s minor allele. The blue line in the main panel shows the estimated effect of the SNP on the phenotype as a function of the PGS, based on a regression model of the phenotype on the SNP, the PGS and their product; the grey band represents a 95% confidence interval. The five orange points show the estimated effect of the SNP in a standard GWAS regression model (without any interaction) in each of five equally sized sample groups partitioned by the quintiles of the PGS; the vertical bars represent 95% confidence intervals. The blue curve atop the main panel is a Gaussian kernel density estimate of the distribution of the PGS; the vertical dashed grey lines show the boundaries of the five quintiles. **b** | A global non-linear relationship between the PGS and the phenotype (left plot, glycated haemoglobin; the black line shows linear regression fit, the blue line a LOESS curve) could lead to false-positive findings. We correct by first applying a rank-based inverse normal transformation to the phenotype (after regressing out covariates) and then applying a ‘mean correction’ to the PGS (Methods), obtaining an approximately linear relationship (right plot). **c** | Additional real-world examples of the SNP effect types identified. From left to right: a SNP with a direct association with the phenotype but no significant interaction with the PGS; significant SNP×PGS interaction signals may occur in the opposite direction to a direct association; or occur (positively or negatively, respectively) despite no significant direct association with the trait.

A chief concern is avoiding false-positive findings, which have been a challenge for epistasis detection in the past (e.g., ref. [30]). To avoid known issues, we first ensure that the PGS used in the regression does not contain SNPs in linkage disequilibrium (LD) with the focal SNP being tested for interaction by removing from the PGS all SNPs on the same chromosome as this SNP (obtaining a ‘leave-one-chromosome-out’ PGS). Provided that population structure and the possible long-range LD it can generate have been well accounted for through the use of Ancestry Components [29], by also removing the possibility of local LD we make the method robust to ‘phantom epistasis’ (where a putative interaction is merely tagging an unobserved variant with an additive effect [31]).

We then ensure that possible global non-linear transformations of the additive genetic component of the phenotype (where the genotype–phenotype map can be approximated by, for example, a sigmoidal function of the sum of additive genetic effects) do not bias our inference. Previously termed ‘higher-order’ or ‘global epistasis’ [27, 32], such effects are of scientific interest but the resulting detected interactions depend only on SNP effect sizes, so do not yield insights about individual variants relative to standard GWAS, and cause a definition of interactions that is dependent on phenotypic scaling. To render the PGS–phenotype map linear and generate results insensitive to phenotypic transformations, we first apply a rank-based inverse normal transformation (INT) to the phenotype (after regressing out covariates), then sort and bin individuals by their PGS, and finally replace each individual’s inferred PGS with the linearly interpolated value of the average phenotype in their bin for that individual’s PGS (Methods), thereby approximating any possible global function linking the PGS and the average phenotypic value (we refer to this transformation as ‘mean correction’; Fig. 1b).

While simple, the model in equation (1) captures a range of interaction effects in real data. As seen in Fig. 1c, in addition to the most common case where a SNP associated with a phenotype in a plain GWAS does not interact with the PGS (first panel), we identify SNPs whose effect is modulated by the PGS by either decreasing (second panel) or increasing in absolute value with this score, and also SNPs whose average effect is null but which have non-zero effects in the lower and upper quantiles of the PGS distribution (third and fourth panels). This last category represents SNPs that would not be linked to the trait by a straightforward GWAS, yet impact it via modulation of effect sizes at *other* SNPs. Unlike, e.g., testing approaches that identify variance-associated markers [33–37], our approach does not give a signal in the setting of gene–environment interactions, making these approaches complementary.

Interactions between polygenic background and individual variants have been examined in recent years for individual rare variants in localised regions known to affect the risk of different diseases such as breast, ovarian and colorectal cancers [38–40], type 2 diabetes and various monogenic metabolic diseases [41–44], obesity, fatty liver disease [43] and heart disease [39, 45–48]. A recent study also investigated interactions with polygenic scores for all variants with additive or variance effects affecting eight lipid traits and coronary artery disease risk [13]. We believe the present study is, however, the first to propose a rigorously tested pipeline for performing such analyses at scale and to examine these effects genome-wide (for variants with frequency of at least 0.1%) for a wide array of quantitative phenotypes. This enables detection of interacting variants regardless of their direct impact on a trait, allowing us to examine the overlap between signals in the original GWAS and the SNP×PGS GWAS, while retaining a manageable multiple-testing burden equivalent to that for standard GWAS.

### Simulations

We undertook extensive simulations to assess the inferential properties of our method. To evaluate the false-positive rate under a null model, we first simulated 72 phenotypes without interactions that span a broad range of genetic architectures using genetic data from the UKB. Simulation parameters are chosen to cover a realistic spectrum while focusing in some cases on settings expected to be more challenging for the method. The simulated phenotypes vary along five dimensions: (i) number of causal SNPs (100, 1000 or 10,000, reflecting the range of polygenicity from molecular to anthropometric traits [2]); (ii) positioning of these SNPs (either placed at random, or half are clustered in regions of 1000 kb with five causal SNPs per region on average to emulate observed instances of multiple independent causal variants in a single locus [49, 50]); (iii) effect size adjustment by allele frequency (none, or adjustment following the LDAK model to reduce the effect size of more common SNPs, where the two possible parameters represent the range of best fit for the model across a variety of phenotypes [51]); (iv) non-linear transformation (none, or scaled sigmoid applied to the combined additive genetic effects either before or after random noise is added to achieve a desired additive heritability – such non-linear transformations could be expected to lead to false positives and we therefore aim to test our method’s robustness to them); and (v) heritability (30% or 60% – these are higher than existing estimates for most traits, accounting for the inclusion of rare variants (MAF ≥ 0.1%) leading to greater heritability [51, 52]) (Fig. 2a; Methods).

**Fig. 2:**
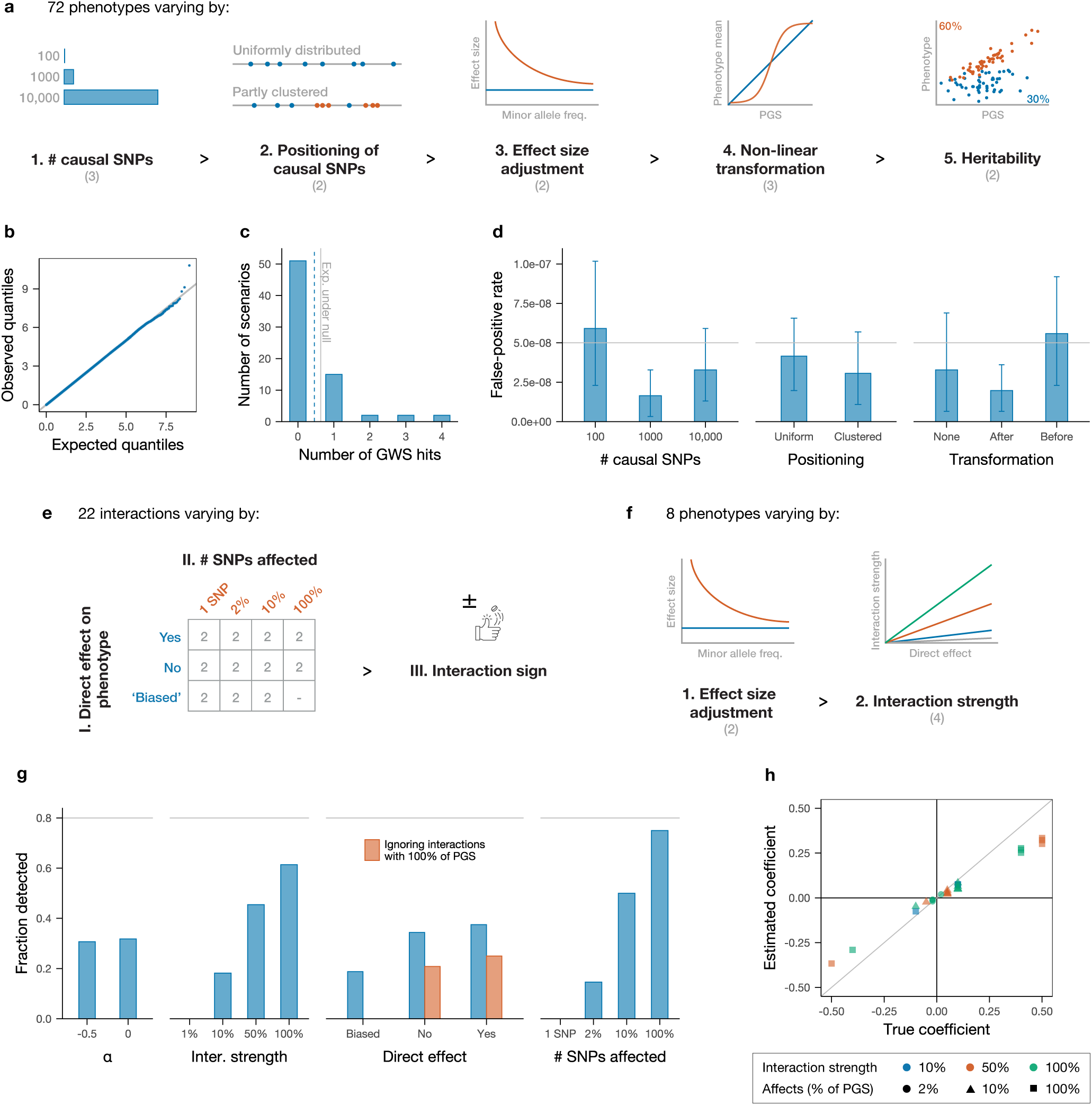
Simulations: setup and results. **a** | Simulated phenotypes without interactions vary in their genetic architecture along five dimensions. The numbers in parentheses under each dimension represent the number of possibilities, yielding 72 phenotypes in total. **b** | Combined Q-Q plot for all 72 traits of the observed p-values for the SNP×PGS interaction term in the interaction GWAS compared with the expected null distribution. Q-Q plots for traits grouped by the different parameters in **a** are shown in Fig. S3. **c** | Histogram showing the number of simulated phenotypes (scenarios) with different numbers of genome-wide significant hits in the interaction GWAS. The vertical dashed blue line represents the average number of genome-wide significant hits across all scenarios and the vertical solid grey line the expectation under the null. **d** | Combined false-positive rate across groups of simulated traits organised by number of causal SNPs (number 1 in **a**, left), their positioning (number 2 in **a**, middle) and whether and how a global transformation is applied (number 4 in **a**, right). The horizontal solid grey lines indicate the expected value under the null. Error bars represent 95% confidence intervals estimated through block bootstrapping. Fig. S4 shows corresponding plots for numbers 3 and 5 in **a**. **e** | Simulated interactions are driven by 22 focal SNPs. These vary in whether they have a direct effect on the phenotype in addition to their interactions; how many SNPs they affect; and the sign of these interactions. **f** | Simulated phenotypes with interactions vary along two dimensions, yielding eight different traits with the same set of causal SNPs. **g** | Fraction of interactions detected across all phenotypes, grouped by effect size adjustment (number 1 in **f**, first plot from the left), interaction strength (number 2 in **f**, second), presence and type of direct effect (number I in **e**, third) and number of SNPs affected (number II in **e**, fourth). Fig. S5 shows corresponding plot for number III in **e**. Overall, 55 of the 176 interactions are recovered. **h** | Scatter plot of estimated vs. true coefficients under blue scenario in panel **a**, number 3 (α = 0; Fig. S6 shows orange scenario with α = −0.5), with colour/shape indicating interaction strength/number of SNPs affected; diagonal line shows *y* = *x*.

After simulating phenotypes, we analysed each simulation exactly as we analysed real data: by performing a standard GWAS and constructing a PGS, then using this to test each SNP for interactions (Methods). Across all 72 phenotypes, the distribution of p-values for the interaction term in equation (1) is well calibrated under the null (Fig. 2b), and the average number of genome-wide significant hits (defined as p ≤ 5 × 10^−8^) is at or below the expected rate (Fig. 2c) in all settings. Analysis of false-positive rates by simulation parameters show only non-significant trends (Figs. 2d and S3).

We then simulated a variety of interactions to evaluate power and understand the type of epistatic effects that we can expect to detect in real data. Starting from a null architecture with 1000 causal SNPs placed at random, which comprise the (true) additive ‘PGS’ for the new simulated trait, we selected 22 focal ‘left-hand side’ (LHS) SNPs to interact with different subsets of this PGS. These 22 SNPs vary along three dimensions: (i) whether they have a direct effect on the trait (eight SNPs do as they are chosen from among the PGS SNPs; the remaining 14 SNPs do not, but six of these will interact primarily with SNPs in the PGS with a positive direct effect, and so are likely to have a positive estimated direct effect (Methods)); (ii) how many SNPs in the PGS they interact with (one, 20, 100 or all 1000 causal SNPs); and (iii) the sign of the interaction (each LHS SNP affects all PGS SNPs with which it interacts with the same sign, either increasing or decreasing their effect in absolute value; this is set at random with equal probability) (Fig. 2e).

Having defined the set of additive effects (the PGS) and interactions (driven by the 22 LHS SNPs), we generated eight different phenotypes with the same set of causal SNPs (including interactions) whose architecture varies along two dimensions: (i) whether the effect sizes of both additive and epistatic effects are adjusted by allele frequency as for the null simulations; and (ii) the magnitude of the interactions (set to 1%, 10%, 50% or 100% of the effect size of the interacting SNPs in the PGS) (Fig. 2f).

After applying our data analysis pipeline to these simulated traits, we unsurprisingly find that interactions with larger effect sizes and which affect a greater fraction of PGS SNPs are more likely to be recovered (Fig. 2g). In real data, we do not expect to recover interactions between only two SNPs unless their effect is very strong, and expect to mostly recover interactions involving a sizeable fraction of PGS SNPs. Comparing the inferred coefficients with their predicted ‘true’ values (after accounting for the ‘attenuation’ of inferred coefficients for interactions with only a fraction of the PGS) shows that this approach is quantitatively accurate, with only slight underestimation of true effect sizes (Fig. 2h), presumably due to imperfect construction of the PGS. Together, these simulations suggest our approach to be overall robust, well calibrated, and able to detect gene–gene interactions of sufficient strength.

### SNP×PGS interactions

We applied our approach to identify interactions involving 97 quantitative phenotypes (Table S1) in a sample of approximately 344,000 individuals of ‘White British’ ancestry in the UKB. Fig. 3a uses triglyceride levels as an example to compare the results of a standard GWAS with those for the SNP×PGS interaction GWAS. The associations from the standard GWAS are spread genome-wide, but interaction signals are much more specific, with only three significant peaks, mapping near the *APOA5*, *LIPC* and *PNPLA3* genes. After filtering through iterative regression (Methods), the *LIPC* peak was found to comprise two independent signals as listed in Fig. 3b. Comparing interaction strength as measured through the log-transformed p-value for each SNP in the original vs. the interaction GWAS for a representative set of traits (Figs. 3b–c; Fig. S7), there are clear differences between the two so that interaction signals cannot be predicted from additive associations, and the strongest interactions do not simply correspond to the strongest direct association signals. Interestingly, this holds not only for unlinked loci in different genomic regions but also in many cases within regions nearby interaction signals (Fig. S8). This suggests that interactions might reflect highly specific functional mechanisms that are specific to individual hits (for example, perhaps reflecting perturbed binding of individual TFs or gene expression in certain tissues), rather than being driven more simply by the genes they are nearby. Below, we highlight related findings whereby distinct coding mutations in the same gene, *APOE*, are associated with interactions for different traits.

**Fig. 3:**
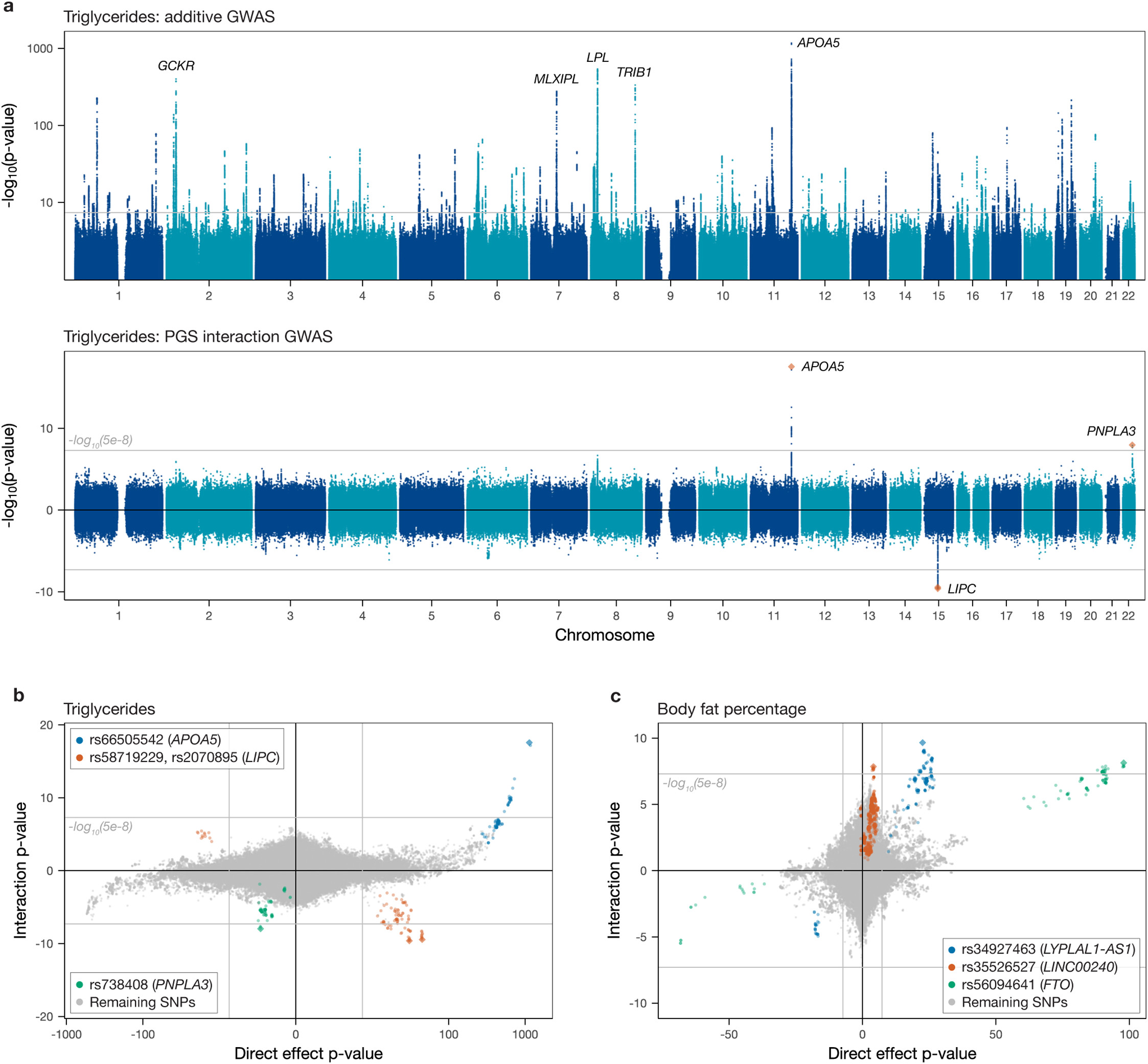
Comparison of standard (additive) and interaction GWAS results. **a** | *Top:* Manhattan plot of standard additive GWAS results for triglycerides in blood. Genes of the top five peaks are shown; the horizontal grey line represents the genome-wide significance threshold of 5 × 10^−8^. *Bottom:* Miami plot of interaction GWAS results for the same trait, with signs given by the product of the signs of the coefficient estimates in the standard and interaction GWASs. Genes of the three independent interaction peaks are shown. **b** | Scatter plot of signed (by the sign of the coefficient) log-transformed p-values in the standard (vertical axis) and interaction (horizontal axis) GWASs for triglycerides. SNPs in LD (squared Pearson correlation between unphased genotypes) of 30% and higher with, and within 1 Mb of, each of the three independent hits are coloured as indicated in the legend. The solid grey lines correspond to the genome-wide significance threshold. **c** | As **b** for body fat percentage.

Overall, we observed 144 independent genome-wide significant (p ≤ 5 × 10^−8^) associations for 52 of the 97 traits tested (Table S6 and Fig. 4, with the latter highlighting sharing across phenotypes after collapsing SNPs in strong LD as documented in Table S7; of these 144 signals, 81 remain significant at a stricter threshold of 5 × 10^−8^/97). Interestingly, most discovered interactions involved molecular phenotypes (primarily biochemistry markers circulating in blood and counts of blood cell types, as shown in Figs. 4a and 4b, respectively), with additional signals for various measures of body fat (Fig. 4c), something we discuss further below. Of our SNP×PGS GWAS associations, 38 did not show an initial GWAS signal for the same trait, including SNPs in *GREB1*, *PNPLA3*, *TAOK1* and the *HLA* region.

**Fig. 4:**
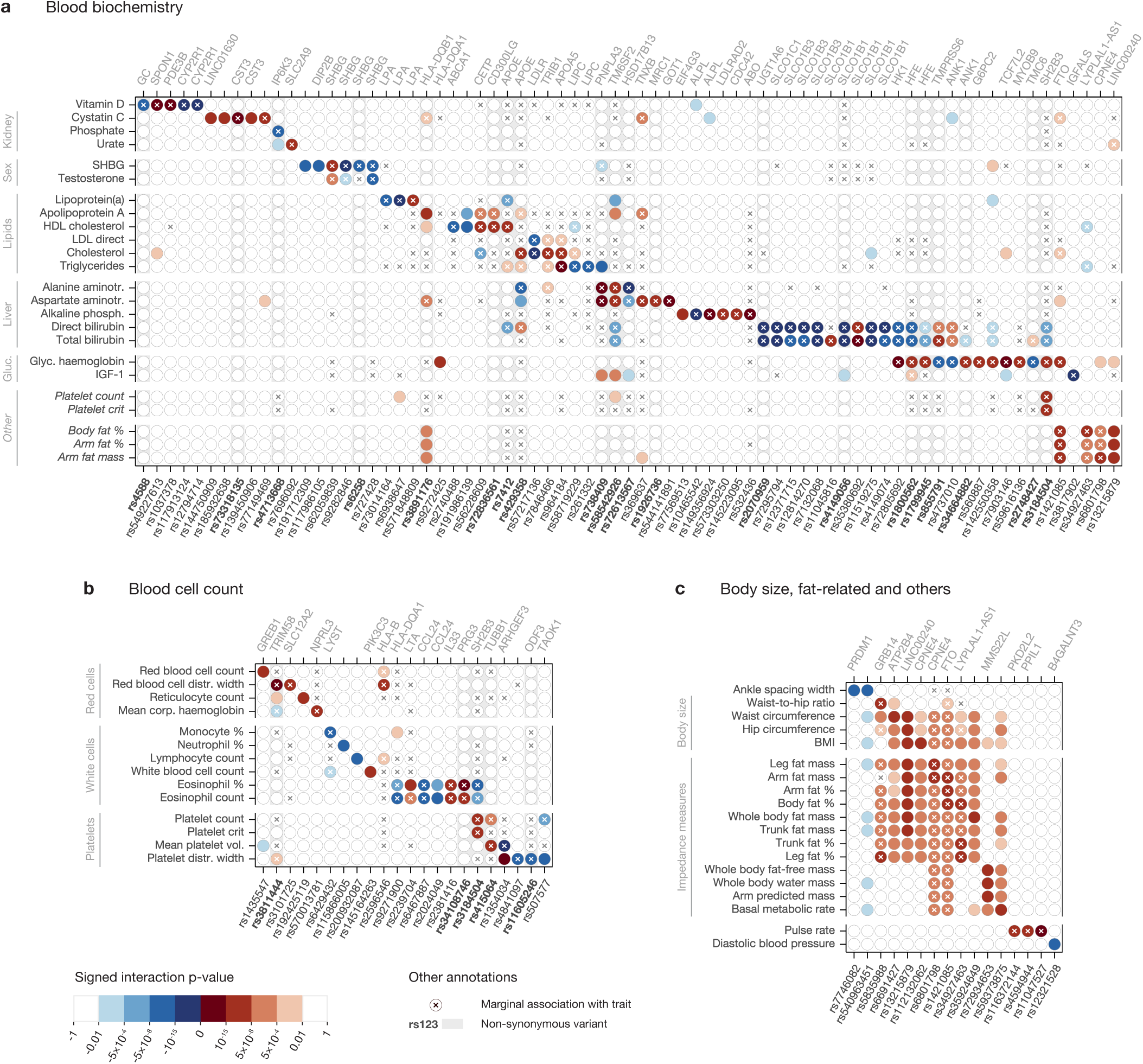
Overview of all 144 independent SNP-by-PGS interaction hits identified for 52 phenotypes. **a** | Interaction p-value (signed by interaction coefficient) of all the independent interaction hits across all phenotypes (among the 52 with at least one interaction hit) in the blood biochemistry trait category. Genome-wide significant marginal associations are denoted with a cross; non-synonymous variants have a boldface label and a grey background in the corresponding column. Five traits not related to blood biochemistry are included as the five bottom rows to highlight overlap with traits in that category. **b, c** | Equivalent plots for blood cell count (**b**), and body size, fat-related and remaining traits (**c**). The signed interaction p-values plotted in this figure are listed in Tables S8a–c.

Non-synonymous mutations are well represented among associations, with 33 of 144 of this type (Table S6; Methods), and there are several cases where apparently independent coding and non-coding associations localise within the same gene (Fig. 4). Interactions include the genes directly encoding components of measured phenotypes for ALP (*ALPL*), cystatin C (*CST3*), lipoprotein lipase (*LPA*), sex hormone–binding globulin (*SHBG*), total and LDL cholesterol (*LDLR*) and triglycerides (*LIPC*). Multiplicative impacts for coding variants seem natural in cases such as these where many or all pathways to generate the observed molecular phenotype operate via the indicated polymorphic gene.

Given a genome-wide significant interaction signal for a specific phenotype, examining the strength of interaction for the remaining 96 traits reveals in several cases a weaker but suggestive signal that would be significant at a more lenient Bonferroni-corrected threshold (0.05/96 ≈ 5 × 10^−4^; Fig. 4 shows such signals in lighter colours for subsets of related traits with at least one genome-wide significant hit; results for all 97 traits are listed in Table S9), and we observe structure among traits (Fig. 4), despite a high level of overall diversity in interactions seen. For example, HDL cholesterol shows little overlap with either LDL or total cholesterol, but the latter two measures have correlated signals among themselves and with triglycerides, which in turn shares hits with levels of alanine and aspartate aminotransferases. Extensive overlap occurs for body fat–related traits, the majority of which identify a significant interaction involving *LINC00240* as well as signals at *CPNE4* and *FTO*, among others. The interacting *FTO* SNP (rs1421085) is noteworthy as it was one of the first polymorphisms to be linked to obesity [53, 54]: the fact that it interacts with the PGS for several obesity-related traits therefore suggests that its impact is partly mediated through polygenic background. Among other SNPs, rs3184504, a missense variant of *SH2B3*, shows an interaction signal for the largest number of non-fat-related traits, including some that are not obviously closely connected (eosinophil percentage, platelet count and crit, and glycated haemoglobin). This gene is a negative regulator of intracellular cytokine signalling, is involved in haematopoiesis and has been linked to several autoimmune and cardiovascular diseases [55]. Other overlaps across less obviously connected traits include between glycated haemoglobin and bilirubins (at *HK1*, *HFE* and *TMPRSS6* in particular), and body fat percentage, arm fat percentage and arm fat mass (*FTO*, same SNP as above).

We compared our findings with those of a previous study of SNP×PGS interactions for lipid traits [13]. This study provides a natural comparison to our results, although we note testing was restricted to candidate variants showing marginal effects on traits (mean or variance – employing a lower significance threshold accordingly) and used an approach that did not incorporate the corrections we apply to linearise the PGS–phenotype relationship, employed a distinct PGS construction method and did not regress out some of our covariates. In that study, 16 significant hits were identified for traits that we also analysed, in a joint analysis of Icelandic, British UKB and Danish samples; however, only two of these reached our significance threshold (p ≤ 5 × 10^−8^) using the UKB samples alone, so we expect limited overlap. Interestingly, we identify a larger number (15) of interactions for these traits. Three of these (or a closely associated tag variant) were identified by Snaebjarnarson *et al.* [13] for the same phenotype (Table S10), while another was identified in that study for different lipid traits, suggesting they are real. The differences in approach described above likely explain the remaining non-overlapping hits: for example, an *HLA-DQB1* variant for apolipoprotein A, which lacks a marginal association signal in our results, also did not show either a mean or a variance effect in their results and hence was not tested; while, e.g., associations for cholesterol near *LDLR* and *TRIB1* and for triglycerides at *APOA5* are identified as having variance or marginal epistasis impacts in other studies [26, 37] (Table S10), suggesting (along with our simulation results) that they represent genuine interactions.

We also compared our results with those of different, but related, approaches testing for marginal epistasis [26] and variance effects [34–37, 56–58] applied for overlapping phenotypes. Comparing with testing for marginal epistasis, we recover five of 16 interactions found in a study of 53 quantitative traits in the UKB [26] (Table S10). We also see overlap with the results of tests for genetic effects on phenotypic variance. While we do not observe an effect for rs7202116 impacting body mass index (BMI) as reported by Yang *et al.* [34], we detect this region as interacting with the PGS for the related traits of body fat percentage (rs56094641), arm fat mass and arm fat percentage (rs1421085 in both cases). Similarly, we detect two of the seven loci reported to affect BMI dispersion by Young *et al.* [35] (those in *LYPLAL1* and *FTO*) as SNP×PGS hits for traits related to BMI. Across the remaining five studies, we find that 38 of our 144 hits match variance effect signals for the same phenotypes, including 32 hits reported for 15 biomarkers in the UKB by Lyon *et al.* [37] (Table S10; some of our hits are seen in more than one previous study). This overlap of variance-influencing mutations with our associations suggests that some (though likely not all) previous hits indeed reflect the gene–gene interactions that our approach is designed to specifically identify, and adds confidence in our results.

### SNP×SNP interactions

Having identified a considerable number of independent SNP×PGS interactions, we then leveraged these signals to find SNP×SNP interactions by running a GWAS of pairwise interaction for each SNP×PGS interaction hit. As this required running only a few GWASs for each of the 52 phenotypes for which we had identified SNP×PGS interactions, the number of statistical tests performed for each phenotype was of the same order of magnitude as a standard GWAS, therefore incurring only modest computational cost and requiring the usual Bonferroni correction for multiple testing. This procedure identified 38 independent genome-wide significant pairwise interactions for 21 traits for which both SNPs have a frequency of at least 1% (Table S11), and an additional 45 interactions with an interaction frequency (defined analogously to the minor allele frequency for a single SNP) of at least 0.1% (Table S12). Six of the 38 common-variant interactions have been previously reported in the literature [10, 11, 13, 26, 37, 59–61] (Table S13) while the remainder are novel to our knowledge.

Fig. 5a shows the results of the pairwise interaction GWAS for rs635634 (a SNP near *ABO*) for ALP levels in serum, a clinical biomarker commonly used in the diagnosis of liver and bone diseases. Of the five significant peaks, the signals at the *PIGC*, *TREH*, *FUT6* and *FUT2* genes were shown to be driven by a single SNP each through iterative regression while the *MIS18A* hit was discarded as it did not meet either frequency threshold. Fitting a regression model including all possible pairwise interactions between these five SNPs (Fig. 5b) confirms that the four interactions identified through the GWAS are jointly significant while also detecting a significant interaction between *TREH* and *FUT2* and a suggestive interaction between *PIGC* and *FUT2* significant at the 10^−4^ level. All five SNPs in this interaction network are non-synonymous or closely tag such a SNP (Fig. 5b).

**Fig. 5:**
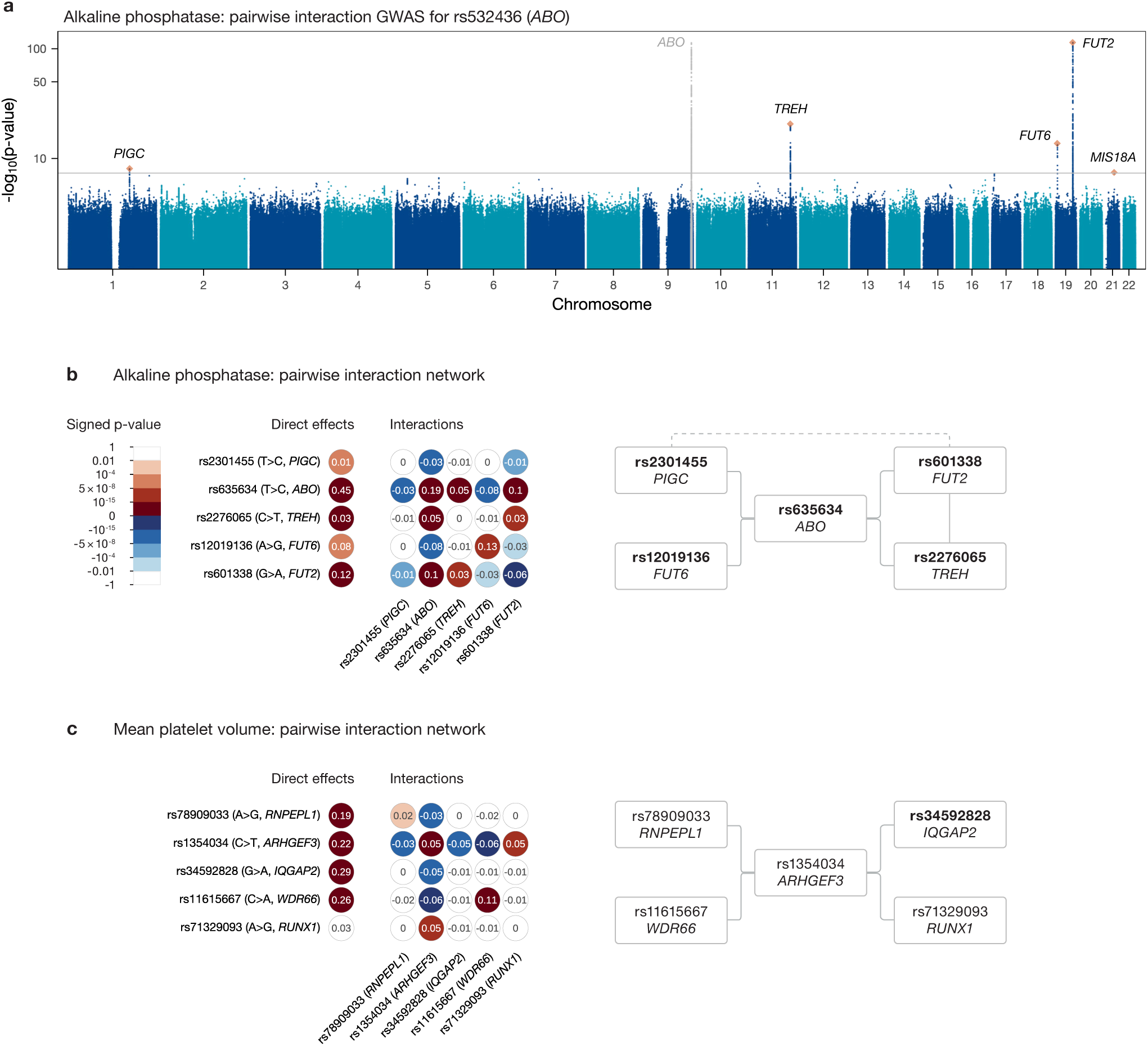
Example of pairwise interaction GWAS and resulting interacting networks of SNPs and genes. **a** | Manhattan plot of pairwise interaction GWAS between rs532436 and every other SNP for ALP. SNPs within 1 Mb of this SNP are grey; these interaction signals are potential false positives driven by LD. Genes at the five interaction peaks are displayed, with the independent SNPs at each peak shown as orange diamonds. **b** | *Left:* Coefficients (numbers within each circle) and p-values (colours, signed by coefficient) from joint linear regression model with main effects (left-most column; effect allele was chosen to give positive main effects), squared allele count/dosage terms to account for possible dominance or recessiveness (diagonal matrix elements), and pairwise interactions (off-diagonal elements; we use rs601338 as the *FUT2* SNP here because it is the accepted causal mutation for European non-secretor status [62]). *Right:* Corresponding network of pairwise interactions: solid lines show strongly evidenced interactions (p ≤ 5 × 10^−8^), dashed lines p ≤ 10^−4^. All SNPs are in bold, signifying they are non-synonymous or tag (*r*^2^ ≥ 80% within 1 Mb) a non-synonymous variant. **c** | Corresponding pairwise interaction network for mean platelet volume. On the right, bold type again denotes non-synonymous SNPs.

The interaction between rs635634 (*ABO*) and rs601338 (*FUT2*), first identified in the Japanese population [10] and more recently replicated in the UKB [37], highlights the potential for epistatic signals to reflect relevant biological mechanisms. Circulating ALP consists of tissue-nonspecific ALP (encoded by *ALPL* and produced in the liver and bones), which accounts for more than 80% of the total [63], and intestinal ALP (encoded by *ALPI* and produced by the intestinal epithelium). Intestinal ALP (IALP) levels in serum have long been known to depend on ABO blood group (determined by *ABO*) and ABH secretor status (release of ABO antigens into bodily fluids, determined by *FUT2*): they are higher for B and O types than for A or AB, and for secretors than non-secretors [64], with the latter having much lower (by 80% [65]) levels of IALP (tissue-nonspecific ALP does not exhibit these dependencies [65]). While the differences across blood types operate in the same direction regardless of secretor status, they are significantly more pronounced for secretors than non-secretors [19]: this is thought to reflect a blood group–dependent mechanism for elimination of IALP from serum where, for secretors, red blood cells of groups A and AB bind to IALP to a much greater extent than those of groups B and O, leading to more rapid elimination [18, 19]. The main effects of, and statistical interaction between, rs635634 (*ABO*) and rs601338 (*FUT2*) in a regression model that includes only these SNPs and their interaction (Table S11) track these differences: rs635634 accurately predicts A vs. non-A blood type, with the minor allele T being associated with A types^1^ [66] and therefore having a negative direct effect on ALP; similarly, rs601338 G>A is a stop-gain variant causing non-secretor status and the wildtype allele G therefore has a positive effect; finally, their interaction has a negative effect, reflecting the fact that the magnitude of the effect of rs635634 is larger for non-secretors.^2^ Interactions between *ABO* and *FUT2* mutations or between the ABO blood group and secretor phenotypes more generally have also been reported to affect serum lipids [13, 67], the levels of 55 proteins in a recent proteomics study of the UKB [68] and risk of various diseases including Crohn’s disease [69], early childhood asthma, *Streptococcus pneumoniae* [70], pancreatic cancer [71] and coronary artery disease [13], underlining the functional importance of this biological pathway.

This epistatic network also includes previously reported interactions between *ABO* and *TREH* and between *TREH* and *FUT2* [37], as well as novel interactions of *ABO* with *PIGC* and *FUT6*, and a suggestive interaction between *PIGC* and *FUT2*. These three additional genes appear functionally relevant in this context: IALP is made in gut epithelial cells in which *TREH* is specifically expressed; both are tethered to the brush border of epithelial cells through GPI anchoring, and *PIGC* is involved in the biosynthesis of these lipid anchors; finally, *FUT6* is tightly linked with *FUT3* (the primary gene determining Lewis phenotype, which is connected with ABH secretor status and has also been linked to ALP levels in a similar manner [72]) and itself influences Lewis phenotype, with these blood group antigens also being manufactured in gut epithelial cells. In the case of the *ABO*–*FUT2* interaction, the functional knowledge necessary to interpret its significance had been discovered prior to the identification of the epistatic signal; for the remaining interactions, further research is necessary to understand the mechanisms underlying the statistical signals.

Another pairwise interaction network occurs for mean platelet volume (MPV; Fig. 5c). It is centred around *ARHGEF3* and involves genes and variants with functional relevance. *ARHGEF3* is a master regulator of megakaryopoiesis, being involved in vasculogenesis and erythropoiesis [73]; the interacting SNP, rs1354034, is a common intronic variant previously associated with various platelet-related traits (as well as with other blood cell types including reticulocytes and lymphocytes) in both European and Asian populations [74–77]. The *RNPEPL1* SNP, rs78909033, has been linked to platelet and red blood cells [75, 78], while rs34592828, an *IQGAP2* variant, has been associated with platelets and haemoglobin [78]. The fourth gene involved, *WDR66*, was identified in one of the first platelet GWAS as being associated with MPV [79], and rs11615667, the SNP in question, has also been linked to platelets [80–82]. Finally, *RUNX1* plays a crucial role in haemotopoiesis and mutations at this gene have been connected to a variety of haematological malignancies, including leukemia; the variant rs71329093 has been found to associate with platelet count and crit [77]. Interestingly, both *ARHGEF3* and *WDR66* are part of an interaction network for platelet distribution width (Fig. S9b), strengthening the evidence for a meaningful functional role for these interactions.

Network plots for additional traits for which we identified interactions involving more than two SNPs are shown in Fig. S9. A noteworthy interaction is that between rs2381416 (*IL33*) and rs34210653 (*ALOX15* missense variant) for eosinophil percentage (the same interaction involving a different but highly correlated *IL33* variant is also seen for eosinophil count). *IL33* encodes a cytokine expressed in various tissues including the lung. It plays an important role in exacerbating inflammation in allergic diseases through eosinophil activation [83], with *IL33* levels found to be elevated in patients with allergic airway diseases [84]. The specific variant in this interaction has been associated with asthma risk [85]. *ALOX15*, its interacting counterpart, encodes 12/15-Lipoxygenase, present in eosinophils and epithelial cells and important in inflammatory response, having been implicated in multiple inflammatory diseases [86]. The interacting SNP, rs34210653, has been associated with eosinophils [75, 78]. Of particular relevance is the fact that a functional interaction has been found recently between these proteins in the context of a murine model of asthma: genetic deletion of mouse *ALOX15* exacerbated IL33-induced eosinophilic airway inflammation [87], and it was found that *ALOX15*-expressing eosinophils are responsible for attenuating inflammation [88]. This novel statistical interaction may therefore be explained by conservation between mice and humans of this functional interaction between the two proteins, both of which have been proposed as potential therapeutic targets for asthma [83, 88].

A pair of interactions involving *APOE* occurs for alanine aminotransferase (ALT) and lipoprotein(a) (LPA) levels. For ALT, an enzyme produced primarily in the liver whose levels in serum are used to assess hepatic health, we identify an interaction with positive sign between *APOE* missense variant rs429358 and rs2176521, an intergenic variant in chromosome 3. For LPA, a lipoprotein whose function is still poorly understood but which is associated with cardiovascular disease risk [89] and whose components (apolipoprotein(a) and apolipoprotein B-100) are assembled in the liver [90], we report a negative interaction between a different *APOE* missense variant, rs7412, and rs571848809, an intronic SNP of the *LPA* gene. These two *APOE* SNPs are well known as they jointly determine which of three isoforms the APOE protein takes. They are also the strongest genetic risk factors found for Alzheimer’s disease (AD), with rs429358 increasing risk and rs7412 being protective [91]. The interactions identified between these two major AD genetic factors and liver-related proteins are suggestive in light of a proposed link between liver function and this disease [92].

### Interactions with partitioned PGSs

Through our SNP×PGS interaction pipeline, we identified a large number of such effects for a variety of phenotypes, and only a minority of these possess clear SNP×SNP pairwise interactions (previous section) that might partially describe their effects. While the remaining loci might genuinely interact with the entire PGS, another possibility, highlighted by our simulations, is interactions mainly involving only particular functional components of the PGS, and identifying these could strengthen the estimated signals and facilitate understanding of the underlying biology.

We therefore explored an initial approach to divide each trait’s PGS into functionally defined components for downstream testing. Given the likely importance of gene regulation in phenotypic architecture, and the natural connection between regulatory networks and interactions, we used data on TF binding to deconstruct each PGS into components, with each component including those SNPs that potentially disrupt the binding sites of a particular TF. Information on TF binding preferences was obtained from the HOCOMOCO v13 database, comprising 1611 binding motifs for 1120 unique human TFs [93]. As there are no established methods, we developed a new statistical approach to perform this task weighting binding motif information. Given a PGS comprising a set of independently associated SNPs and corresponding effect sizes, we identify (Methods) probabilities for each possible causal variant using their association signal as well as information on each variant’s functional severity (using predicted consequences from Ensembl VEP [94]) and overlap with H3K4me1 and H3K4me3 methylation marks in a relevant tissue for the phenotype in question (likely marking enhancers and promoters, respectively, in that tissue; we use ChIP-seq peaks obtained from the Blueprint ChIP-seq Consortium via the International Human Epigenome Consortium Data Portal [95]). Then we sum over potential causal variants to obtain, for each TF, a score estimating how likely each potential causal SNP is to fall within an active (i.e., bound) motif for that TF. This score is defined by first computing a binding score for the motif in a narrow window of 60 bp around each SNP using a standard approach. An initial quantile-based score is then scaled to reflect the odds of finding a score of this value for SNPs within enhancers vs. SNPs not in regulatory regions (Methods), assuming that enrichment reflects functional enhancer copies in order to estimate, separately for each motif/TF, which scores might truly correspond to bound motif copies. Summing scores weighted by causal probabilities over all SNPs results in a collection of TF-specific PGSs upweighting those SNPs plausibly bound by each respective TF. Our resulting score for each TF is unlikely to perfectly reflect true TF binding, but nonetheless testing using these scores vs. the PGS as a whole tests for different behaviours of some SNPs within the PGS vs. others. Leave-one-chromosome-out versions are made for all scores and used in interaction testing as before. The regression model also includes the regular (non-partitioned) PGS and its interaction with the SNP (Methods).

Testing for interactions with TF-partitioned PGSs (TF-PGSs) for each of the 144 SNP–trait pairs for which a SNP×PGS interaction had been previously identified yielded 12 independent associations for eight different traits (Table S14) significant at a conservative Bonferroni-corrected threshold of 10^−5^ (Methods), and some in each direction relative to the original interaction. Thus, although this is just one illustrative initial approach to partition PGSs and others (e.g., based on nearby gene ontology) should be explored in future, for at least some cases our genetic interactions possess significantly stronger effects for identifiable subsets of variants within the PGS. One interaction of interest involves a TF known to regulate the interacting SNP’s associated gene in a trait-relevant context, supporting the possibility of interaction between the two genes. We identify an interaction between a *TCF7L2* intronic SNP and PGS SNPs altering sites bound by the KDM2A TF for glycated haemoglobin (HbA1c) levels, a biomarker of blood glucose widely used in the diagnosis and management of diabetes. TCF7L2, which is also a TF, is a key regulator of glucose metabolism and an important component of the Wnt signaling pathway [96], which is also involved in glucose homeostasis and plays a key role in the aetiology of diabetes [97]. KDM2A has also been shown to (negatively) regulate genes within this pathway both by direct binding to DNA targets [98], and by physically interacting with key Wnt components including TCF7L1/2 and other *TCF* /*LEF* genes, and nuclear Beta-catenin. The interacting SNP at *TCF7L2*, rs7903146, is the most strongly associated type 2 diabetes risk variant in the GWAS Catalog [99–101]. KDM2A binds to unmethylated CpG islands through its DNA-binding CXXC zinc finger domain, and then demethylates H3K36me1/2 residues and acts as a negative regulator of gluconeogenic gene expression [102, 103]. A recent study found that KDM2A interacts physically with TCF7L2 through this same CXXC zinc finger domain, promoting its degradation [104]. Our observations might be explained by the fact that these TFs operate in the same individual cells and signalling pathway simply multiplying their impacts, or may reflect stronger connections such as competition for binding sites and/or these physical interactions between them. We also find an interaction affecting levels of cystatin C between a SNP near the *CST3* gene (which encodes cystatin C itself) and targets of the EGR1/EGR3 TFs. This is interesting given reports of strongly reduced *Cst3* expression following *Egr1* elevation in *Egr1*-transfected mice neurones [105], and following overexpression of *Egr1* in the amygdala of transgenic mice [106], suggesting *CST3* itself is a target of the EGR1 TF, at least in the mouse brain [105, 106].

## Discussion

Statistical interactions between genetic variants have been hypothesised since the early days of quantitative genetics but their identification has been hampered by multiple technical challenges and their role in the architecture of human phenotypes remains a topic of intense debate. On the one hand, it is well established that additive models of genetic effects are sufficient to capture a large fraction of genetically driven phenotypic variation in human populations, and this apparent additivity is unsurprising, due to the fact that most discovered variants have very small effect sizes. Moreover, despite extensive methodological efforts and empirical studies, to date few epistatic signals have been robustly identified for human phenotypes. On the other hand, even subtle interaction signals, by reflecting interdependencies between different functional loci, could shed light on biological mechanisms and aid in the functional understanding of GWAS signals, which remains a major challenge. Some examples of interactions so far identified (such as those affecting ALP levels and ankylosing spondylitis risk mentioned in the introduction) lend strength to this ambition. Overall then, we do not expect incorporation of interactions to strongly improve genetic predictions, but the extent to which identified additive associations partially also reveal underlying epistatic signals and, if identifiable, how helpful these interactions might be in understanding biology remains an open question.

In this work, we sought to make progress towards answering these questions by conducting a large-scale search specifically targeting gene–gene (i.e., not pure gene–environment) interactions, for which we might hope to have statistical power: between single genetic variants and scores aggregating additive genetic effects on a phenotype. We developed a simple statistical method for this task and evaluated it through extensive simulations. Application to 97 quantitative traits for a set of White British individuals in the UKB identified 144 independent SNP×PGS interactions for 52 different traits, revealing that identifiable signals exist for most traits. These SNP×PGS hits allowed us to identify 38 SNP×SNP interactions for 21 traits, revealing functional relationships among them and significantly increasing the known set of such effects. We also found an additional 12 associations between SNPs and groups of variants within the PGS overlapping binding sites for particular TFs.

A first conclusion from our work is that interactions involving sets of SNPs aggregated into scores appear common, extending previous research that explored these effects in more restricted settings and demonstrating that non-linear epistasis signals ‘hidden’ within GWAS data are quite commonplace. These signals differ strongly from those captured by standard direct association tests in both strength and direction. For example, interactions are identified (e.g., at the *HLA* or at *PNPLA3*) that involve SNPs showing no direct marginal association with the focal trait, while many strong directly associated variants do *not* show interaction evidence. Given that it may be applied to any quantitative trait GWAS dataset, with only the same multiple-testing burden as the initial direct GWAS, we suggest that performing a secondary SNP×PGS interaction GWAS might represent a worthwhile and largely risk-free routine endeavour. It also complements existing methods that search for genetic modulators of phenotypic variance [33–37], which might reflect both gene–gene and gene–environment interactions. Although it is not always straightforward to identify the biological basis of interactions, our results suggest that SNP×SNP testing as well as partitioning of genome-wide PGSs into specialised components determined according to prior biological information can be successful as targeted downstream analyses. For the latter, we used primarily one type of information (TF binding motifs) but our testing approach is agnostic. Future research could therefore leverage other sources of functional knowledge (such as additional epigenetic marks or gene sets associated with different pathways) for PGS partitioning. More generally, we have shown that statistical interactions can capture meaningful biological relationships, a key hypothesis that drove our work and motivates further research.

Given that only a minority of trait-impacting variants show interaction signals, a natural question is whether they show particular properties. One clear signal is that non-synonymous coding variants appear overrepresented in both SNP×PGS and SNP×SNP interactions, explaining over 20% of our signals. This seems natural for molecular phenotypes measuring the product of a single or small number of genes, and indeed several of our cases are of this type: we might expect multiplicative effects operating on many or all upstream regulatory variants impacting expression of the corresponding gene. For more complex phenotypes, interactions might instead occur at a subset of markers regulating the corresponding gene, so deconstruction of these cases might be of particular interest. More generally, because non-synonymous mutations impact the corresponding protein product in all tissues, it seems natural that they are especially likely to be involved in interactions. This might explain also why non-synonymous site hits sometimes show interaction signals across a range of our studied phenotypes (e.g., *SH2B3*, *TM6SF2*, *PNPLA3*, *HLA-DQA1*). Despite this, the majority of our signals are non-coding, and so likely to be regulatory in nature.

Another clear pattern is that we tend to find a large number of interactions for ‘simpler’ molecular phenotypes such as blood biochemistry biomarkers and counts of different blood cell types, and few (often none) for complex phenotypes such as height, though with some notable exceptions for body-composition traits such as waist circumference. This occurs despite these complex traits possessing many direct GWAS signals, and we suggest that it does not reflect a fundamental lack of interactive pathways impacting these traits, but rather a lack of statistical power to find them. Since the architecture of highly polygenic phenotypes (such as height) likely aggregates the effects of many molecular ‘intermediate’ traits, there might be many interactions, but with each involving only a small fraction of the overall set of loci represented within the PGS. Our simulations confirmed limited power to identify such signals, because we test the entire PGS against SNP genotype. A highly promising avenue for future research is therefore to perform functional partitioning of PGSs for complex traits and to perform interaction testing using these components.

We note some limitations of our work. Most importantly, our current approach and analysis are restricted to quantitative phenotypes. Developing and implementing a statistical approach for binary case/control phenotypes would be a valuable direction for future work. Our ‘linearisation’ of the relationship between the PGS and the trait is important to prevent false positives, and would require extending in the setting of binary traits to instead predict probabilities. A second limitation is that we cannot currently test for interactions in *cis* because this risks false positives (which led us to build leave-one-chromosome-out PGSs for interaction testing). This challenge is not specific to our approach, however [30], and recent methodological developments may allow for such testing in the future [107]. Lastly, as noted above we have limited power to detect interactions for more polygenic phenotypes. These limitations do not cast doubt on the reliability of our current findings but rather point to promising opportunities for further research.

## Methods

### Data

#### Phenotypes

We analyse 97 quantitative phenotypes (Table S1), including the majority of traits available from the blood assays (biochemistry and blood cell counts), physical measurements (body size, body fat measured through impedance, blood pressure and handgrip strength) and urine assays categories, as well as a selection of traits of general interest from other categories.

#### Samples

Our main dataset consists of 343,969 samples of ‘White British’ ancestry as defined in the original UKB publication [28].

#### Variants

We use the autosomal genotype dosages obtained through imputation based primarily on the Haplotype Reference Consortium provided by the UKB. We apply four filters (minor allele frequency ≥ 0.1%; genotype missing rate ≤ 5%; Hardy-Weinberg equilibrium p ≥ 10^−10^; imputation quality score ≥ 0.8) to obtain a subset of 12,690,793 variants for analysis.

#### External annotations

We use annotations on functional consequence and associated gene for each variant from the Ensembl VEP [94], RefSeq Gene [108] and Gencode [109] databases.

### Testing for SNP×PGS interactions

We present below a brief summary of the main steps in our method; a detailed description with motivating considerations is provided in the Supplementary Information.

#### Controlling for covariates and applying a Normal transformation

We begin by regressing out the following covariates from each phenotype: age at recruitment and its square; sex; age×sex and age^2^×sex; genotype measurement batch; assessment centre attended; and Ancestry Components to correct for population stratification [29]. We then apply INT to the resulting residuals to avoid potential false-positive associations driven by phenotypic outliers.

#### PGS construction

We first build a PGS using a simple Clumping & Thresholding (C+T) approach with a very loose threshold of 90% for LD (squared Pearson correlation) between neighbouring variants within a 1 Mb window in the LD Clumping step, and with a p-value threshold determined based on predictive performance on a held-out set of samples. This initial C+T approach achieves our goal of identifying a very broad set of variants associated with the phenotype but incurs the cost of including many highly correlated neighbouring variants. To correct the coefficients of neighbouring variants for ‘double counting’, we develop and apply an iterative algorithm that adjusts their effects to approximate the result of fitting all variants simultaneously in a multiple regression model. This two-step approach produces a PGS that accounts for all the additive signal of the large set of variants it contains: if we regress out the score from the phenotype and then test each variant in the PGS for association with the resulting residuals, none of these will be genome-wide significant by construction. In accounting for all the direct/additive signals of a large set of associated variants for each trait, we aim to increase power to detect interaction/non-additive signals in downstream testing.

#### Leave-one-chromosome-out PGS to prevent LD-driven false positives

An important consideration when testing for epistatic effects is whether, rather than representing true departures from an additive model, putative interactions might merely reflect the (additive) effect of an unobserved third variant. Termed ‘phantom epistasis’, this source of confounding has been examined theoretically [31] and found to be problematic also in practice [30, 110]. Since it is driven by LD between the two variants tested for interaction and a third variant (which is not considered in the test), this issue is of concern primarily when the variants in question are in close proximity. To avoid detecting false-positive interactions driven by correlations between the variant tested for interaction with a PGS and other variants included in that PGS, we build 22 leave-one-chromosome-out (LOCO) versions of each PGS and use these in all our tests.

#### Adjusting for a global non-linear functional form of the genotype–phenotype map

Research into the genetic architecture of a wide range of complex human phenotypes has convincingly shown that – for the purpose of explaining phenotypic variance due to genetic variation using a statistical model – their functional form is well approximated by a linear, additive combination of weighted genetic effects [14, 15]. However, for this additive form to be appropriate, it may be necessary to apply a non-linear transformation (such as a sigmoid) to the additive combination of genetic factors to render them onto the scale of the phenotype. Such generalised non-linear transformations, termed ‘higher-order’ or ‘global epistasis’, are of scientific interest (and have been examined previously [27, 32]) but are not the focus of this project and, more importantly, may produce spurious interactions if not addressed (for example, if the transformation is approximately quadratic, this will generate an interaction term for every pair of variants). In particular, ‘interaction’ terms generated this way are simple functions of the effect sizes of each variant, so are unlikely to lead to insights about underlying trait biology not provided by the initial GWAS, and so we regard such signals as false positives here and attempt to avoid their occurrence. To prevent false-positive interactions driven by global non-linearities, after obtaining a PGS we transform it using a function built in a data-driven manner to approximate a possible non-linear transformation of the combined additive genetic effects. We begin by sorting individuals by their PGS in increasing order and divide them into bins of 1000 (with 20 smaller bins of 50 at either end of the distribution); for each bin, we compute the median of the PGS and the mean of the phenotype (after regressing out covariates and applying INT); finally, we perform linear interpolation of these values (the PGS median and the phenotype mean in each bin providing the X and Y coordinates, respectively) and replace each individual’s score with the interpolated value of the true phenotype for their PGS.

### SNP×PGS interaction GWAS

Having obtained a LOCO PGS, we proceed to run our main GWAS by fitting a simple multiple linear regression model for each SNP separately. The dependent variable is the residuals obtained above by regressing out covariates and applying INT and from which we additionally regress out the PGS (non-LOCO, to account for as much additive signal as possible) and its interactions with assessment centre and Ancestry Components variables (to account for possible interactions with aggregate genetic measures of population stratification). The regressors are an intercept term, the minor allele count/dosage of the SNP, the PGS and the product of the SNP’s allele count/dosage and the PGS, which is the primary term of interest (equation (1)).

#### Identifying independent interactions that are not explained by dominance effects

We conclude the testing for SNP×PGS interactions by distilling the results of each GWAS into a set of independent genome-wide significant hits to prioritise for further investigation. We run the following iterative regression procedure: sort all genome-wide significant SNPs in decreasing order of significance; beginning with the most significant SNP, run the same SNP×PGS interaction test as before but now add a term for the square of the SNP’s minor allele count/dosage; if the interaction remains genome-wide significant, assign this SNP to the set of independent hits and otherwise remove its terms (main effect and its square, and interaction with the PGS) from the model; add the second most significant SNP to the model and proceed analogously until all genome-wide significant SNPs have been considered. In including a term for the square of each SNP’s genotype, we are controlling for possible dominance or recessiveness effects which might otherwise confound our estimates of interactions.

The list of independent SNP×PGS interactions we identified in this manner is provided in Table S6. For each interaction hit, we include annotations from Ensembl VEP, RefSeq Gene and Gencode as well as an indicator of whether it is non-synonymous. We consider a hit to be non-synonymous if either the SNP itself or one of its tags (with *r*^2^ ≥ 80% within a 1 Mb window) has a predicted consequence at least as severe as ‘protein altering variant’ in the VEP database. In addition to the results of a regression model including the SNP’s minor allele count/dosage, its square, the PGS and the interaction term, we also list in Table S6 the results of direct association testing (i.e., a simple GWAS) for each variant–trait pair as well as those of an interaction model in which we do not regress out from the phenotype the PGS and its interactions with Ancestry Components and assessment centre variables, and also scale the PGS to have zero mean and unit variance, to obtain an interpretable scale for all coefficients.

### Identifying the targets of interaction signals (I): pairwise interactions

SNPs found to interact with a PGS through the approach just described may in fact interact with only a (potentially small) subset of SNPs in that PGS if this interaction is strong enough. If that is the case, it would be relevant to identify the narrower set of SNPs driving the interaction as this would make biological interpretation and further investigation easier.

We begin our search for more specific ‘targets’ of SNPs interacting with a PGS by searching for pairwise interactions with other SNPs. Given the low cost (both computational and in terms of the multiple-testing correction required) of performing pairwise tests for the relatively small set of SNPs identified through the method above (fewer than 150 in our results), we test for a pairwise interaction between each SNP found to interact with the PGS for a given phenotype and all other SNPs in the genome (rather than simply those in the PGS). The dependent variable in these regressions is the original trait from which demographic variables are regressed out and INT is then applied (we do not regress out the PGS and its interactions with Ancestry Components and assessment centre variables as in the SNP×PGS interaction GWAS). We run an iterative regression procedure similar to the one previously described to isolate independent genome-wide significant pairwise hits. We apply further filtering based on frequency to the SNPs surviving iterative regression to eliminate potential false positives: the main set of interactions we report consists of pairs of SNPs which are both required to have frequency of at least 1% (Table S11); we additionally report pairs with interaction frequency (defined as the mean product of the allele counts/dosages of the two SNPs divided by two in analogy with the frequency definition for single variants) of at least 0.1% (Table S12); remaining pairs are discarded.

Compared with the strategy of exhaustively testing for all possible pairwise interactions, this approach narrows down the search space by fixing one of the interacting SNPs. Since these SNPs interact with the PGS, they are plausibly more likely to interact with single SNPs either in that PGS or elsewhere in the genome.

### Identifying the targets of interaction signals (II): partitioning the PGS

To further explore possible targets of the SNPs found to interact with the PGS, we take the natural step of partitioning this PGS into different components in a biologically informed manner. We use information on the functional consequences of different SNPs, on their overlap with methylation marks and, most importantly, on whether they disrupt the binding sites of a large number of TFs.

In this final analysis, for conceptual consistency and computational efficiency, we employ a simpler PGS with fewer variants than the one with which we tested for SNP×PGS interactions: we use a PGS built using C+T with a standard low threshold for LD between variants of 10%. A LOCO version of this PGS will be used in interaction testing as explained below, while the genome-wide significant SNPs resulting from the LD Clumping procedure form the basis of the partitioned scores.

We begin by taking the independent genome-wide significant SNPs for each phenotype (obtained through LD Clumping in the process of building the new PGS) and, to account for the fact that the most strongly associated SNP in an LD block is not necessarily ‘causal’, we extend it to include the tags of each ‘hit’ (or ‘driver’) SNP (*r*^2^ ≥ 75% within a 1 Mb window in our sample). We then make the simplifying assumption that there is exactly one causal SNP in each of the regions defined by a hit SNP and its tags, and set a weight for each tag by computing an approximate Bayes Factor and rescaling them so that they sum to one for each region. Each partitioned score will be obtained by multiplying these Bayes Factors by a weight determined by biological information (as described below) to obtain a final weight that is then multiplied by each SNP’s GWAS coefficient.

#### Partitioning the PGS using functional consequence and methylation mark annotations

We take information on the functional consequences of all SNPs in the PGS from Ensembl VEP [94] and build a binary indicator (denoted 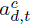, where *d* indexes regions and *t* the potential causal SNPs within each region) of whether each SNP’s predicted consequence is in the top 24 of categories ranked by severity.^3^ For methylation marks, we obtain data on H3K4me1 and H3K4me3 histone marks from the Blueprint ChIP-seq Consortium via the International Human Epigenome Consortium Data Portal [95] for different tissues and cell types (264/306 different datasets for H3K4me1/H3K4me3, respectively); for each dataset and methylation mark, we make a binary indicator of overlap with a ChIP-seq peak for each SNP (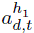 and 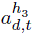, respectively, dataset not indexed).

For each binary annotation, the weights *w*_*d*,*t*_ for each tag SNP are defined as:

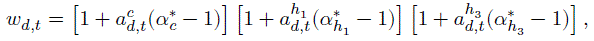

for SNPs with that annotation and set to zero otherwise. These weights represent a model for the prior probability a SNP is causal, under the setting where annotations operate multiplicatively. The 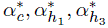 parameters (which we estimate) represent how each annotation either increases or decreases the weight of each tag that has that annotation: the baseline weight for each type of annotation is 1; if the annotation is present (*a* = 1), a value of α > 1 increases the tag’s weight in the PGS and vice-versa for α < 1. An annotation may therefore inform the PGS by either increasing or decreasing the weights of SNPs with that annotation, and we allow these three annotations to operate multiplicatively.

The three α parameters are defined as determining the prior probability of causality for each tag SNP (under the assumption that exactly one tag is causal for each hit as noted above) as follows, and suppressing the indicators *c*, ℎ_1_ and ℎ_3_ for clarity:

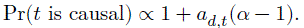

They are then estimated by maximum likelihood in an overall model of which SNPs are causal for the trait. For the histone marks, for which we have multiple datasets (from different tissues and cell types) as annotation sources, we fit the model separately for each dataset and choose the dataset and corresponding parameter that provides the best fit to the data (i.e., the highest likelihood).

#### Partitioning the PGS by TF binding sites

For determining the binding sites of different TFs, we use information on DNA binding motifs for 1120 TFs (1611 unique binding motifs in total) provided in the form of Position Count Matrices in the HOCOMOCO v13 database [93].

We begin by assuming that we have available, for each tag and motif, a score (denoted OR_*d*,*t*_, motif not indexed, again for simplicity) which is equal to one if the SNP has no possibility of falling in an ‘active’ motif for this TF and greater than one otherwise, with a magnitude reflecting the likelihood of this (where ‘active’ means that the TF binds to this motif, causally generating the association observed).

We then extend the approach used above by modelling the prior probability for each tag as being proportional to:

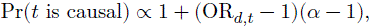

i.e., we substitute the quantitative score in place of the binary indicator of an annotation being present, and otherwise use the same construction. We fit the parameter α for the corresponding TF by maximum likelihood estimation, as above, and define the weights (again, proportional to the modelled probability of being causal) as:

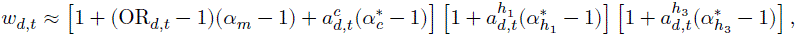

jointly modelling all four types of annotation. Previously, we set the weights of SNPs without the annotation in question to zero; since motif annotations are not binary, we now instead multiply all weights by a quantity approximating the probability that the motif is active. Note that, in the first term above, we assume that the ‘coding’ and ‘TF’ possibilities are biologically distinct and so approximate them as acting additively on the prior.

Finally, we define the scores OR_*d*,*t*_ for each tag and motif by first computing the best binding score for the motif in question by calculating a raw score based on the motif’s highest Position Count Matrix, for potential motif start positions overlapping that SNP. We then rescale this score to provide an odds ratio. We calculate the odds of such a strong match when sampling random SNPs in the genome falling within H3K4me1 sites (we used H3K4me1 sites to represent the overall sequence context of enhancers in the relevant tissue type) vs. SNPs in sites with neither H3K4me1 nor H3K4me3 marks (representing potentially non-functional DNA). Thus, scores corresponding to strong motif matches typically seen much more often in H3K4me1 sites than in SNPs as a whole are given the largest weights. The aim of this approach is simply to identify how ‘good’ a motif match is needed to see such enrichment. Because these weights are later multiplied by α and this is fitted to the data, a relative scaling rather than an absolute enrichment level is what we hope to fit using this approach. See the Supplementary Information for a detailed description of the method.

#### Interaction testing and identifying independent interactions

The regression models used to test for interactions with these partitioned PGSs also include the original (non-partitioned and LOCO) PGS and its interaction with the SNP under consideration:

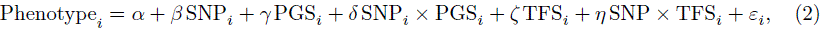

where TFS_*i*_ denotes the TF-PGS. The dependent variable is the same as for the pairwise interaction tests (after regressing out demographic variables and subsequently applying INT, but without additionally regressing out the PGS and its interactions with Ancestry Components and assessment centre variables) while the non-partitioned PGS is a simple C+T score as noted above. Importantly, we therefore only identify an interaction signal for a particular partitioned PGS if it shows a signal that is distinct from that for the PGS as a whole. A stepwise regression procedure is applied to the results of the GWAS to obtain independent hits. Since fewer than 750 TF-PGSs are tested for interaction with each SNP×PGS hit on average – as not all scores are available for all phenotypes (Supplementary Methods) – we set a conservative Bonferroni-corrected threshold of 0.01/1000 = 10^−5^.

Having obtained an initial set of TF-PGS interactions, we finally perform a test of whether any of these signals are in fact explained by a simple pairwise interaction with one (i.e., any) of the driver SNPs in the TF-PGS and/or its tags. This would be undesirable, and potentially suggest a spurious functional interpretation, because it would cast what may be a simple pairwise interaction as an interaction with a wider set of SNPs defined by the binding of a TF, because that interacting SNP by chance significantly contributes to the TF-PGS. For this test, we remove any SNPs significant in the pairwise interaction GWAS and their tags from the TF-PGS and rerun the interaction test (Supplementary Methods). Only those interactions remaining significant after this procedure are reported in Table S14.

### Simulation details

#### Null simulations

We simulate traits without interactions that vary along five dimensions:

- *Number of causal SNPs (3 possibilities):* 100, 1000 or 10,000;
- *Positioning of causal SNPs (2 possibilities):* they are either placed uniformly at random anywhere on the genome; or half are placed into regions of physical length 1 Mb at an average density of five per region, while the rest are placed at random;
- *Effect sizes (2 possibilities):* drawn independently from standard Normal distributions. These are then either transformed by multiplying each variant’s effect size by [2*p*(1−*p*)]^−0.25^, where *p* is the variant’s minor allele frequency, to model a realistic relationship between effect size and allele frequency [51], or simply kept untransformed;
- *Global transformation (3 possibilities):* the combined additive genetic effects are either transformed using a scaled sigmoid (which can be applied either before or after random Normal noise is added to achieve a desired level of heritability), or left untransformed;
- *Heritability (2 possibilities):* set to either 30% or 60%.

In total, 72 different traits are simulated and processed through the same data analysis pipeline as the real phenotypes.

#### Simulations with interactions

To build simulated traits with interactions, we start from one of the simplest simulated architectures without interactions (an intermediate number of 1000 causal loci, SNPs randomly positioned along the genome, and no global transformation) and extend it by adding interaction terms.

We first choose 22 ‘left-hand side’ (LHS) SNPs with minor allele frequency greater than or equal to 5% to interact with different sets of SNPs as described below. When evaluating our method’s power on these simulated traits, we will assess whether these 22 SNPs with varying characteristics are detected. Having defined the 22 LHS SNPs, we then choose their ‘right-hand side’ (RHS) targets. Finally, we fix the resulting set of LHS–RHS SNP pairs and create *eight different traits* that vary only in whether the effect sizes (of both additive effects and interactions) are transformed using allele frequency as in the third bullet point in the previous section and in the magnitude of the effect sizes of the interactions.

Interactions within each trait vary along three dimensions:

- *Presence of a direct effect on the trait:* of the 22 LHS SNPs, eight are randomly chosen from among the 1000 SNPs with a direct effect on the trait; eight are randomly chosen from the SNPs without an effect on the trait; and six are also chosen from those SNPs without a direct effect on the trait but interact primarily (at a four-to-one proportion) with SNPs with a positive direct effect (‘biased interaction’);
- *Number of RHS SNPs that each LHS SNP interacts with:* one SNP among the set of 1000 causal SNPs, 2% of these SNPs, 10%, or the full set of causal SNPs;
- *Sign of the interaction:* each of the 22 LHS SNPs will interact with all their RHS targets with the same sign, drawn with equal probability. If it is positive, it will increase (in absolute value) the main effect of the SNPs with which it interacts, and vice-versa if it is negative;

while traits (all of which share the same additive effects and interactions driven by the 22 LHS SNPs) will vary along two axes:

- *Magnitude of interaction effect sizes (4 possibilities):* the interaction pairs will have an effect that is 1%, 10%, 50% or 100% of the effect size *of the RHS SNP*. Interactions can therefore be thought of as rotating the increasing line representing the correlation between PGS and trait;
- *Possible transformation of effect sizes (2 possibilities):* the effect sizes of the 1000 causal SNPs with an additive effect and of the interactions are either transformed by multiplying by [2*p*(1 − *p*)]^−0.25^ (where the frequency *p* is that of the RHS SNP in the case of interactions) or left untransformed.

## Data availability

UK Biobank data is available for use by approved researchers (https://www.ukbiobank.ac.uk/). Ensembl VEP annotations can be accessed at https://www.ensembl.org. RefSeq Gene and Gencode annotations can be accessed through Annovar (https://annovar.openbioinformatics.org). The H3K4me1 and H3K4me3 ChIP-seq datasets used can be obtained from the International Human Epigenome Consortium Data Portal (https://epigenomesportal.ca/ihec/). TF binding motifs are available through https://hocomoco13.autosome.org/. PGS coefficients and association testing results generated in this project will be made available upon publication.

## Code availability

All data processing and analysis scripts used in this project will be released under an open-source license upon publication. They are currently available from the first author upon request.

## Acknowledgements

This research has been conducted using the UK Biobank Resource under Application Numbers 27960 and 103076. We thank all participants of the UK Biobank study. We thank the Wellcome Trust for funding through grants [222334/Z/21/Z] to L.A.F.F. and [212284/Z/18/Z] to S.R.M. L.A.F.F. was partially supported by National Institutes of Health grant HG011432 to Molly Przeworski. Computation used the Oxford Biomedical Research Computing (BMRC) facility, a joint development between the Wellcome Centre for Human Genetics and the Big Data Institute supported by Health Data Research UK and the NIHR Oxford Biomedical Research Centre. Financial support was provided by the Wellcome Trust Core Award [203141/Z/16/Z]. The views expressed are those of the authors and not necessarily those of the NHS, the NIHR or the Department of Health. For the purpose of Open Access, the authors have applied a CC BY public copyright licence to any Author Accepted Manuscript version arising from this submission.

## Footnotes

1 In the subset of UKB individuals used for the regression in Fig. 5b for which ABO blood type information is available, 77.9% of individuals homozygous for C at this SNP have non-A blood types; in contrast, 99.2% of heterozygous or homozygous for T individuals have A types.

2 Note that by including all five SNPs and their interactions in the model in Fig. 5b we have flipped the sign of the effect of rs601338 compared with a model with only the *ABO* and *FUT2* SNPs and their interaction.

3 These are all categories with ‘high’, ‘moderate’ or ‘low’ IMPACT rating, as well as the top two categories with ‘modifier’ rating (‘coding sequence variant’ and ‘mature miRNA variant’).

