## Supplementary information for "Interactions with polygenic background impact quantitative traits in the UK Biobank"

29 December 2025

**References** **46**

**Supplementary tables** **49**

**Supplementary figures** **56**

### 1 Data

#### 1.1 Phenotype selection

In selecting the phenotypes for this analysis, our aim is to have as large and diverse a selection of quantitative traits as possible while ensuring adequate power to detect effects of interest. To this end, we begin by selecting the majority[1] of traits from the following categories in the UKB data:

- Blood assays:
  - Blood biochemistry;
  - Blood count;
- Physical measures:
  - Blood pressure;
  - Body-size measures;
  - Hand-grip strength;
  - Impedance measures;
- Urine assays.

In addition to the above, we also select a smaller list of traits of general interest (excluding technical measurements related to sample collection, for example) from the following more specialised categories:

- Bone densitometry of heel;
- Fluid intelligence;
- Numeric memory;
- Memory tests (pairs matching card game);
- Prospective memory (recollection of shapes and colours);
- Reaction time;
- Spirometry;

summing to a total of 103 quantitative phenotypes. We then filter this combined list to obtain a final set of phenotypes.

Given the central role played by polygenic scores (PGSs) in our method, as will be explained in detail below, we choose traits for which we expect to be able to build a PGS with reasonable predictive power. To assess this, we use estimates of each phenotype's genetic heritability (specifically, additive common single-nucleotide polymorphism (SNP) heritability estimated through LD Score Regression [1] for many phenotypes in the UKB by Benjamin M. Neale and collaborators [2]) and set a threshold of 15%.

In addition to choosing traits that are moderately heritable, we also restrict our analysis to traits available for a large number of individuals. We retrieve the sample size available for each phenotype from the heritability dataset[2] and apply a threshold of 100 000 individuals.

[1] When multiple closely related traits are available (for example, measurements from both left and right arms or legs), we keep only one.

[2] The full set of samples considered in the Neale Lab's heritability analysis is comprised of approximately 361 000 people of European ancestry. This is similar to, but slightly larger than, the set of unrelated White British individuals that we use for our analysis.

In selecting the final list of traits, we allow a lower heritability value to be compensated for by a larger sample size and vice-versa (e.g., if a trait has a heritability estimate that is 5% greater than 15% but a sample size 5% smaller than 100 000, we consider it to (exactly) satisfy our threshold).

This selection procedure yields 84 quantitative traits from the following six main categories:

- Blood biochemistry;
- Blood count;
- Blood pressure;
- Body size measures;
- Impedance measures;
- Urine assays;

and eight traits from these six more specialised categories which we group into an 'Other traits' category in our results:

- Bone-densitometry of heel;
- Fluid intelligence score;
- Hand grip strength;
- Pairs matching;
- Reaction time;
- Spirometry.

To these 92 traits, we add five others that we compute based on existing traits:

- Waist-to-hip ratio;
- Leg length (computed as the difference between standing and sitting heights);
- Relative leg length and sitting height; and
- FEV1/FVC ratio (forced expiratory volume in 1 second divided by forced vital capacity).

In total, we analyse 97 traits. These are listed in Table S1 with their heritability estimates and sample sizes.

##### 1.2 Covariates

We account for the following individual covariates in our linear regression models:

- Age (at recruitment) and its square;
- Sex;
- Age $\times$ Sex and Age$^2$ $\times$ Sex;
- Genotype measurement batch (encoded as a set of 105 binary variables);
- Assessment centre attended (encoded as a set of 21 binary variables);
- 'Ancestry Components'.

Ancestry Components (ACs) were designed as an alternative to Principal Components (PCs) when correcting for population stratification in GWAS analyses [3]. They are a set of 127 variables corresponding to different regions within the UK, Europe and the world that form a partition of an individual's ancestry. Using ACs has been shown to achieve better control for stratification in GWASs of quantitative traits in the UKB when compared to PCs [3] and, unlike linear mixed model–based statistical methods, can be employed by simply regressing out these variables from the phenotype of interest, which makes them well suited to our statistical approach.

##### 1.3 Sample selection

We begin by identifying all individuals for whom all covariates as well as imputed genotype data are available; there are 487 255 such individuals. We then filter these into three subsets as we now describe.

###### 1.3.1 Main set of unrelated White British individuals

All our primary analyses are performed on a set of 343 969 unrelated individuals of 'White British' (WB) ancestry. To identify these individuals, we rely on a previously defined set used in the statistical analyses in the main UKB publication [4] and which we obtained directly from the authors. To define this subsample, Bycroft et al. identify a maximal set of individuals who satisfy the following conditions:

- Report their ethnic background as 'British';
- Have very similar ancestral backgrounds according to Principal Component Analysis (using PCs 1 to 6);
- Have imputed genotype data available;
- Have an inferred sex that matches their self-reported sex;
- Are unrelated (i.e., with no 3$^{rd}$-degree relative or closer as inferred through estimation of kinship coefficients).

We take the intersection of this set and the full set above to obtain 343 969 individuals.

###### 1.3.2 Validation and test sets

While our main analyses are undertaken on the set of 343 969 unrelated WB individuals just discussed, for parameter tuning and evaluation of PGS performance it is important to have separate sets of observations who are similar to those in the main sample but which are not directly used to construct the scores.

For this purpose, and following standard practice in the literature, we define 'validation' and 'test' sets of 10 000 and 9071 individuals, respectively.

To define these two subsets of samples, we first identify all individuals in the full sample whose ancestry (as measured through ACs) is at least 90% from Great Britain (we exclude Ireland) but who are *not* part of the subset of unrelated WB individuals above (52 729 individuals).

Then, using the estimates of kinship coefficients[3] between pairs of individuals related to the 3$^{rd}$ degree or closer provided in the UKB, we filter out individuals related to the 2$^{nd}$ degree or closer[4] with someone in the WB set above (this gives us 32 148 individuals).

Finally, we remove individuals from this set to ensure that they are also unrelated to one another while aiming to keep as many individuals as possible. For this, we begin by removing

[3] These were computed with the software package KING [5].

[4] This corresponds to a kinship coefficient of at least 0.0884.

individuals with the highest number of relatives until each person appears only once in the list of related pairs, and then exclude half of these at random.

This procedure gives us 19 071 individuals from which we randomly sample 10 000 to make the validation set while the remaining 9071 are assigned to the test set.

##### 1.4 Variant quality control and filtering

We consider only autosomal genetic variation and use the first imputed genotype calls provided by the UKB (unphased dosages based on the Haplotype Reference Consortium reference panel as well as the UK10K and 1000 Genomes Project datasets) for 93 095 623 variants.[5] To this set of variants, we apply the following filters:

- Minor allele frequency (MAF) $\geq 0.001$;
- Genotype missing rate $\leq 0.05$;
- Hardy-Weinberg equilibrium p-value $\geq 10^{-10}$;
- 'INFO' imputation score $\geq 0.8$.

The four filtering variables above are standard in the literature (see, e.g., refs. [4, 6, 7]). The first filter ensures that the SNPs used are not very rare; this is important as rare SNPs can lead to false positive associations, as we discuss below. With the second and third filters, we aim to remove SNPs with potential genotyping errors. Finally, the imputation score filter corresponds to a filter on effective sample size.[6]

Applying the filters above to the set of imputed variants[7] yields a total of 12 690 793 variants which are used in all analyses.

##### 1.5 Functional annotations

When reporting association results, we include information on the likely function of each variant and the gene within which it lies (or which it is predicted to affect) obtained from three distinct sources (all in assembly GRCh37/hg19 to match the UKB imputed data):

- The Ensembl Variant Effect Predictor (VEP) database (release 113) [8];
- The RefSeq Gene database (last updated 17/08/2020) [9];
- The Gencode Basic collection (v46, last updated 13/05/2024) [10];

where the RefSeq and Gencode datasets were obtained and processed through Annovar [11].

[5]Note that multi-allelic sites are recorded as separate variants.

[6]Bycroft et al. [4] report that an information score of 0.3 corresponds to an effective sample size of approximately 150 000. Our imputation score filter is therefore stringent in requiring the imputation quality to equate to a large effective sample size.

[7]We compute MAF, genotype missing rate and Hardy-Weinberg equilibrium p-values for our main set of WB individuals only rather than relying on estimates for the whole UKB sample. The INFO imputation score is taken directly from the variant quality-control information supplied with the dataset.

### 2 Methods

#### 2.1 A linear model of a quantitative phenotype

The statistical framework underlying our approach is a linear model for a quantitative trait which is decomposed as the sum of additive, dominant/recessive and possible epistatic genetic effects, the contributions of multiple covariates (these are primarily demographic – age, sex and a measure of genetic ancestry) and a zero-mean stochastic error term.

Consider a sample of $N$ individuals indexed by $i = 1, \dots, N$ and a set of $K$ positions along the genome indexed by $k = 1, \dots, K$, and denote by $x_{ik}$ the genotype (minor allele count[8]) of individual $i$ at position $k$. Denoting the (mean-centred) phenotype by $y_i$, we can write this model as follows:

$$y_i = \sum_{k=1}^{K} \beta_k x_{ik} + \sum_{k=1}^{K} \gamma_k \mathbb{I}(x_{ik} = 1) + \sum_{k<k'} \delta_{k,k'} x_{ik} x_{ik'} + \\ + \eta a_i + \theta b_i + \sum_{m=1}^{M} \iota_m c_{im} + \varepsilon_i, \qquad (1)$$

where:

- $\beta_k$ is the 'additive' or 'direct' effect of SNP $k$;
- $\mathbb{I}(\cdot)$ is the indicator function and $\gamma_k$ a possible dominant/recessive effect of SNP $k$;
- $\delta_{k,k'}$ is the epistatic effect – which may be zero – of the interaction between SNPs $k$ and $k'$;
- $\eta$ and $\theta$ represent the effects of age and sex, respectively;
- the set of $M$ variables indexed by $m = 1, \dots, M$ with effect $\iota_m$ represents additional covariates (including a measure of ancestry); and
- $\varepsilon_i$ is a zero-mean stochastic error term which is independent and identically distributed (i.i.d.) across individuals.

We note that a trait may need to be scaled or transformed (e.g., through a logarithmic function) for this model to be appropriate – this will be discussed in more detail below.

We also note that it is possible for a SNP to affect the phenotype only by modulating the effect of another SNP (e.g., it is possible for $\delta_{k,k'} \neq 0$ while either $\beta_k = 0$ or $\beta_{k'} = 0$), and we do find examples of such SNPs in our analysis of real data.

Lastly, we have not included gene-environment interactions in the model above as we will not examine such non-linear effects in this work but these may be relevant for the phenotypes analysed and are worthy of further study.

#### 2.2 The simplest approach to interaction testing

The most immediate approach to testing for interactions between $P$ SNPs is to test for a possible interaction between each pair of SNPs. The computational cost of such a method is quadratic in

[8]We note that all the derivations below, including the results in Section 2.4.1, are also valid for a continuous genotype variable representing the 'dosage' of a SNP inferred through imputation, as will be the case for most of the variants we analyse in real data.

the number of SNPs considered, as with $P$ SNPs one would need to perform $\binom{P}{2} = P(P-1)/2$ tests.

There are two important drawbacks to exhaustively testing for all possible pairwise interactions in this way. First, given the large number of SNPs which are routinely available in genetic datasets, carrying out $P(P-1)/2$ tests can be very expensive computationally. However, this challenge can be partly overcome through the use of modern computing infrastructure – by parallelising operations using graphical processing units (GPUs) or a cluster of central processing units (CPUs) – or computational methods that leverage high-efficiency data structures and operations, such as bitwise computing [12].

The second drawback is the low statistical power that methods that rely on such a large number of statistical tests will necessarily have. This challenge is more severe than that posed by their computational burden because it is inherent to any statistical approach that undertakes a large number of independent statistical tests, and so cannot be solved with more computational resources or greater efficiency in computation.

##### 2.3 Testing for SNP × PGS interactions

We propose a new approach to detecting interactions that does not require exhaustive pairwise testing, is computationally tractable and achieves adequate power.

Consider a PGS for a trait $y$ denoted by $\hat{S}_i$ for individual $i$ which is defined as a weighted sum of mutation counts at a set of $L$ positions along the genome indexed by $l = 1, \ldots L$, i.e., $\hat{S}_i = \sum_{l=1}^{L} \hat{\beta}_l x_{il}$ for coefficients $\hat{\beta}_l$ for each of $L$ positions. Ideally, this PGS would correspond exactly to the set of additive effects in the 'true model' given in equation (1) by $\sum_{k=1}^{K} \beta_k x_{ik}$. In practice, however, we will need to rely on an empirically derived PGS for which both the SNPs included and their coefficients may be different from the set of SNPs and coefficients in the true PGS.[9] We will discuss below how this will affect our inference and the steps we take to minimise its impact.

Given an inferred PGS $\hat{S}_i$ for each individual, we propose to test for statistical interactions genome-wide between each SNP $x_{ij}$ under consideration and the PGS. More specifically, we fit the following model once for each SNP in the genome present in the data:

$$\begin{aligned} y_i = \beta_j x_{ij} + \kappa \hat{S}_i + \lambda_j x_{ij} \hat{S}_i + \\ + \eta a_i + \theta b_i + \sum_{m=1}^{M} \iota_m c_{im} + \varepsilon_i, \end{aligned} \tag{2}$$

where the first three terms correspond to the additive effect of SNP $x_{ij}$, the direct effect of the estimated PGS $\hat{S}_i$ and an interaction term between the SNP and the PGS, respectively, and the remaining terms are as before in equation (1).[10]

[9] We denote the 'true' PGS by $S_i$ and the estimated PGS by $\hat{S}_i$; similarly, $\beta_l$ represents a true coefficient and $\hat{\beta}_l$ its estimated counterpart.

[10] In practice, we will first 'regress out' the covariates from the phenotype and then run regressions with only the first three terms as this greatly reduces computational cost. The practical aspects of our computational implementation are discussed below in Section 2.6.

Our motivation to look for interactions between single genetic variants and a PGS is two-fold. First, this approach requires only $P$ tests and is therefore a possible solution to the computational challenges and low power of exhaustive approaches.

More importantly, this approach has greater power than standard methods to *find genetic variants that interact with multiple other variants.* We consider this to be a significant advantage as we expect such interactions to exist: given the importance of gene regulatory networks – where a SNP that impacts the expression of one gene can have downstream effects on many others – we expect some SNPs to interact statistically with mutations in multiple loci along the genome and have designed a method that targets this type of effect.

##### 2.4 Potential sources of type I and II errors

Our aim is to identify statistical interactions between a variant and one or more other variants that are significantly associated with a quantitative phenotype after taking into account possible confounders – we want to detect interactions that reflect true biological interdependencies between genetic mutations rather than spurious effects driven by statistical artefacts.

With this in mind, we now proceed to consider our method's propensity to lead to inferential errors of either type I or type II, i.e., to false-positive or false-negative findings, respectively. We beging by examining two potential sources of false negatives before considering four factors that may lead to false positives. In the next section we will discuss how we address these different sources of error.

###### 2.4.1 Asymptotic properties of the OLS estimator of the interaction term

We consider potential sources of false negatives by deriving the bias of the ordinary least-squares (OLS) estimator of coefficient $\lambda_j$ in equation (2) in two simplified settings. We again consider a sample of $N$ individuals indexed by $i$ for whom we observe a quantitative phenotype $y_i$. We assume the following simple true model for $y_i$ which contains both additive and interaction genetic effects:

$$y_i = S_i + \lambda_j x_{ij} R_i + \varepsilon_i, \tag{3}$$

where $S_i$ is the PGS (i.e., an aggregation of all additive effects), $x_{ij}$ is a SNP, $R_i$ is part of the PGS $S_i$ as explained next, and $\varepsilon_i$ is a zero-mean error term with finite variance which is i.i.d. across individuals. For simplicity and without loss of generality, all variables are assumed to have mean zero and $x_{ij}$ is also assumed to have variance one.

We further assume that the PGS $S_i$ can be decomposed as follows:

$$S_i = Q_i + R_i, \tag{4}$$

where

$$Q_i := \sum_{o=1}^{O} \beta_o x_{io} \tag{5}$$

$$R_i := \sum_{p=1}^{P} \beta_p x_{ip}. \tag{6}$$

Under this model, therefore, $y_i$ is driven by an additive genetic component $S_i$ as well as by an *interaction* between a single SNP $x_{ij}$ and a fraction $R_i$ of the PGS.

As noted previously, in real applications we do not observe the true PGS $S_i$ and instead have to rely on an approximation or estimate $\hat{S}_i$. Here we will assume that this is observed with an *additive and independent* measurement error relative to the true score, i.e.:

$$\hat{S}_i = S_i + \zeta_i, \tag{7}$$

with $\zeta_i$ a zero-mean, finite-variance error which is i.i.d. across individuals.

Equations (4) and (7) imply the following expression for $R_i$, the part of the PGS that interacts with SNP $x_{ij}$:

$$\begin{aligned} R_i &= S_i - Q_i \\ &= \hat{S}_i - \zeta_i - Q_i. \end{aligned} \tag{8}$$

Finally, we make the following assumptions regarding independence relationships and correlations between variables. First, the general error term $\varepsilon_i$ is assumed to be *independent* of the explanatory variables $x_{ij}$, $S_i$ and their product, as well as of $\zeta_i$, which implies independence from $\hat{S}_i$. Second, the variant $x_{ij}$ is assumed to be *uncorrelated* with all the variables related to the PGS and its components and any product of these variables (i.e., $\mathbb{E}(x_{ij}Q_i) = \mathbb{E}(x_{ij}R_i) = \mathbb{E}(x_{ij}S_i) = \mathbb{E}(x_{ij}S_i^2) = \mathbb{E}(x_{ij}\zeta_i) = \mathbb{E}(x_{ij}\hat{S}_i) = \mathbb{E}(x_{ij}\hat{S}_i^2) = 0$, etc.), and similarly for $x_{ij}^2$.

The first assumption is required for the OLS estimator to be unbiased.[11] The second assumption is more interesting as it has biological meaning: assuming that the SNP and the PGS are uncorrelated is equivalent to assuming that they are not in LD. One way to achieve this is by making sure that the PGS does not contain any variant present in the same chromosome as the SNP being tested for an interaction, i.e., building a 'leave-one-chromosome-out' (LOCO) PGS, as we will discuss in Section 2.5.3.[12]

Our goal is to recover the interaction coefficient $\lambda_j$. To this end, we fit the following linear regression model, estimating its coefficients through OLS:[13]

$$y_i = \alpha_0 + \alpha_1 \hat{S}_i + \alpha_2 x_{ij} \hat{S}_i + u_i.$$

We now proceed by formally deriving basic properties of our estimator under different scenarios. We will focus on large-sample properties (in particular, consistency, i.e., convergence in probability[14] of the estimator to the true value of the parameter as the sample size tends to infinity[15]) as these are the most relevant to our application in a setting with many thousands of

[11] For consistency, it is sufficient to assume no correlation between the error term and the explanatory variables.

[12] We ignore long-range LD (which can have multiple sources such as relatedness between individuals in the sample or population admixture) as our sample is of homogeneous ancestry and good care has been taken to remove related samples and correct for population structure.

[13] Not including a main term for $x_{ij}$ in the regression model will greatly simplify the derivations below. When running regressions in practice, however, we always include main terms for all variables.

[14] Consider a sequence of estimators $\hat{X}_1, \hat{X}_2, \dots, \hat{X}_n$, where $\hat{X}_j$ is computed using the first $j$ observations. Convergence in probability of this sequence of estimators to a value $a$ means that, for any $\varepsilon > 0$, $\lim_{n\to\infty} \Pr(|\hat{X}_n - a| < \varepsilon) = 1$, which we denote more concisely by $\hat{X}_n \xrightarrow{\text{p}} a$. See, e.g., ref. [13, Section 5.2].

[15] See, e.g., ref. [13, Section 6.3].

samples. We will begin with a trivial example (Case 1) in which we assume that the interaction in the true model involves the *full PGS* which we *observe perfectly* – this will serve as a baseline for subsequent derivations. We will then relax these two assumptions in turn: first, we will consider a scenario in which the interaction is with only part of the PGS as above (Case 2); then, we will assess the consequences of additive measurement error in the PGS when the interaction again involves the full PGS for simplicity (Case 3).

*Case 1: interaction with the full PGS which is perfectly observed:*

We begin by considering the OLS estimator of $\lambda_j$ under the simplest possible setting: we assume that the SNP $x_{ij}$ interacts with the full PGS and that we observe the true value of this PGS without error, i.e., $R_i = S_i = \hat{S}_i$.

The true model is therefore:

$$y_i = S_i + \lambda_j x_{ij} S_i + \varepsilon_i \tag{9}$$

and we fit the regression model through OLS:

$$y_i = \alpha_0 + \alpha_1 S_i + \alpha_2 x_{ij} S_i + u_i. \tag{10}$$

**Proposition 2.1.** *The OLS estimator $\hat{\alpha}_2$ of the coefficient $\lambda_j$ in the true model given in equation* (9) *obtained from the regression model in equation* (10) *is unbiased, efficient*[16] *and consistent.*

This is presented without proof as these properties follow from classic results for least-squares estimators of linear models under standard assumptions (see, e.g., ref. [14, Sections 1.3 and 2.3]). The crucial assumption underlying this result is that of *independence* between the error term $\varepsilon_i$ and the two explanatory variables in the model, $S_i$ and $x_{ij} S_i$.

*Case 2: interaction with only part of the PGS which is perfectly observed:*

We now return to the setting introduced at the start of this section (equation (3)), assuming that the interaction is with only part of the PGS, $R_i$. However, in contrast with that initial model we now assume that we observe the full PGS perfectly; this will simplify our derivations while still enabling us to draw relevant conclusions on the effect of this 'partial interaction'.

The true model is now:

$$y_i = S_i + \lambda_j x_{ij} R_i + \varepsilon_i \tag{11}$$

but as we do not know $R_i$ we fit the following regression:

$$y_i = \alpha_0 + \alpha_1 S_i + \alpha_2 x_{ij} S_i + u_i. \tag{12}$$

We begin by noting that the least-squares estimator of $\alpha_2$ in the simple two variable linear model $y_i = \alpha_0 + \alpha_1 x_{1i} + \alpha_2 x_{2i} + u_i$ can be shown to be [15]:

$$\hat{\alpha}_2 = \frac{\sum_i x_{1i}^2 \sum_i x_{2i} y_i - \sum_i x_{1i} x_{2i} \sum_i x_{1i} y_i}{\sum_i x_{1i}^2 \sum_i x_{2i}^2 - \left(\sum_i x_{1i} x_{2i}\right)^2}.$$

[16] Efficiency means that it achieves the lowest variance within the class of linear unbiased estimators.

The estimator for the model in equation (12) is therefore:

$$\hat{\alpha}_2 = \frac{\sum_i S_i^2 \sum_i x_{ij} S_i y_i - \sum_i S_i^2 x_{ij} \sum_i S_i y_i}{\sum_i S_i^2 \sum_i x_{ij}^2 S_i^2 - \left(\sum_i S_i^2 x_{ij}\right)^2}. \tag{13}$$

**Proposition 2.2.** *The OLS estimator $\hat{\alpha}_2$ of the coefficient $\lambda_j$ in equation* (11) *converges in probability according to the following expression:*

$$\hat{\alpha}_2 \xrightarrow{\text{p}} \lambda_j \frac{Cov(R_i, S_i)}{Var(S_i)}.$$

*Proof.* See Appendix 2.A. □

Proposition 2.2 shows formally that our ability to detect an interaction between a SNP and the PGS will depend on the *correlation between the fraction of the PGS with which the SNP interacts and the full score:* the estimated coefficient will be *attenuated* (i.e., biased towards zero) with a degree of attenuation inversely proportional to the fraction of the PGS affected.

It is worth noting that $R_i$ and $Q_i$ represent the *weighted* contribution of the SNPs contained in them to the PGS, with weights given by each variant's $\beta$ coefficient. Therefore, if a SNP interacts with a small fraction of the SNPs that comprise the PGS but the SNPs with which it interacts have large effect sizes, this will reduce the degree of attenuation.

In summary, this result implies that our approach will have greater power to detect associations that involve either a large fraction of the SNPs in the PGS, or especially impactful SNPs within that PGS.

*Case 3: interaction with the full PGS which is estimated with additive error:*

We proceed by considering the slightly more complex case under which the PGS is estimated with additive error (i.e., we observe $\hat{S}_i = S_i + \zeta_i$) while, for simplicity, the SNP $x_{ij}$ again interacts with the full PGS as in Case 1.

The true model is:

$$y_i = S_i + \lambda_j x_{ij} S_i + \varepsilon_i \tag{14}$$

but we now fit a regression model with an estimated PGS $\hat{S}_i$:

$$y_i = \alpha_0 + \alpha_1 \hat{S}_i + \alpha_2 x_{ij} \hat{S}_i + u_i. \tag{15}$$

This setting is reminiscent of the classical errors-in-variables model in the statistics and econometrics literatures (see, e.g., [16, Section 9.14], [17, Section 1.1] or [18, Section 8.8.1]). However, unlike in those models, in which the error is purely additive for all the variables observed with error, here the error in our variable of interest, $x_{ij}\hat{S}_i$, is multiplied by the allele count of the SNP under consideration.

We will now derive the impact of this error on the estimator of $\lambda_j$ and show that it will again be attenuated, with the strength of attenuation increasing as the signal-to-noise ratio in the PGS decreases. This is again similar to the basic errors-in-variables model.

Adapting equation (13), we see that the estimator for the model in equation (15) is:

$$\hat{\alpha}_2 = \frac{\sum_i \hat{S}_i^2 \sum_i x_{ij}\hat{S}_i y_i - \sum_i \hat{S}_i^2 x_{ij} \sum_i \hat{S}_i y_i}{\sum_i \hat{S}_i^2 \sum_i x_{ij}^2 \hat{S}_i^2 - \left(\sum_i \hat{S}_i^2 x_{ij}\right)^2}. \tag{16}$$

**Proposition 2.3.** *The OLS estimator $\hat{\alpha}_2$ of the coefficient $\lambda_j$ in equation* (14) *converges in probability according to the following expression:*

$$\hat{\alpha}_2 \xrightarrow{\text{p}} \lambda_j \frac{Cov(\hat{S}_i, S_i)}{Var(\hat{S}_i)}.$$

*Proof.* See Appendix 2.B. □

We see first from Proposition 2.3 that, in the case of no noise in the estimated PGS (i.e., $\zeta_i = 0$ for all $i$), the OLS estimator converges to the true parameter value $\lambda_j$ as we had seen previously in Case 1. What is new in this setting is that, as the expected noise fraction in the PGS increases, the estimate of the interaction coefficient is attenuated towards zero. This implies that the less accurate our PGS, the lower the power we will have to detect interaction signals.

The two simple models above allowed us to assess the impact of two factors – an interaction with only a fraction of the PGS, and a lack of accuracy in the inferred PGS – on the estimates of our parameter of interest. We saw that both factors lead to an attenuation of the estimate of the $\lambda_j$ coefficient and therefore to reduced statistical power to identify a true interaction.

While both of these factors have similar effects on the estimator of the interaction term, it is important to note that they differ in whether or not they can be addressed in practice. The attenuation bias caused by the fact that we test for an interaction with the full PGS while many interactions may involve only a small subset of this PGS is a natural and unavoidable consequence of our testing strategy – it implies that this approach will be more likely to detect interactions with a substantial proportion of the PGS, as we noted above in Case 2. By contrast, the attenuation bias caused by additive error in the inferred PGS can be reduced by using a more 'accurate' PGS. We will discuss below how we may define accuracy in this context and how we will seek to maximise it. For now, having first considered possible sources of false positives, we turn to examining four potential causes of false negatives.

##### 2.4.2 LD as a source of false positives in interaction testing

We begin by discussing the possible pernicious effects of unobserved variants in LD with those being tested for interactions. Consider the toy example of genetic variation for a sample of three haploid individuals $a$, $b$ and $c$ given in the figure below. We assume:

- Three segregating sites in the sample at relative positions 0.2, 0.4 and 0.7 in the genome;
- The genealogical history shown in the figure, which includes a recombination event at position 0.3.

The recombination event means that different parts of individual $b$'s genome coalesce first with either individual $a$ (the left part of $b$'s genome) or individual $c$ (the right part), and leads to $b$ having all three mutations as shown in the genotype table.

| | Position | | |
|---|---|---|---|
| Ind. | 0.2 | 0.4 | 0.7 |
| $a$ | 1 | 0 | 0 |
| $b$ | 1 | 1 | 1 |
| $c$ | 0 | 0 | 1 |

Genotype table

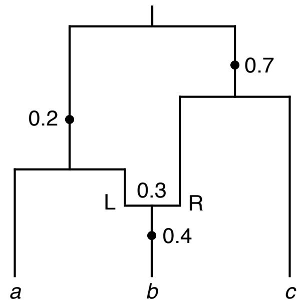

Ancestral recombination graph

**Figure:** Example of genetic variation at three sites with genealogical history.

We define three variables $x_{1i}$, $x_{2i}$ and $x_{3i}$ for individuals indexed by $i \in \{a, b, c\}$ corresponding to mutations at positions 0.2, 0.4 and 0.7, respectively. These variables take the value 1 if a mutation is present for an individual at that site and the value 0 otherwise. Note that, in this sample, we have the following relationship between the three genotypes:

$$x_{2i} = x_{1i} x_{i3} \quad \text{for all } i. \tag{17}$$

We now consider a quantitative phenotype $y$ and assume the following model:

$$y_i = \alpha_1 x_{1i} + \alpha_2 x_{2i} + \varepsilon_i, \tag{18}$$

where $\alpha_1, \alpha_2$ are the coefficients of SNPs $x_{1i}, x_{2i}$ and $\varepsilon_i$ is an error term as before.

Using equation (17), the model in equation (18) can be equivalently rewritten as:

$$y_i = \alpha_1 x_{1i} + \alpha_2 x_{1i} x_{3i} + \varepsilon_i.$$

From this we see that, even though there is no interaction in the original model given in equation (18), a test for an interaction between $x_{1i}$ and $x_{3i}$ could nevertheless suggest an interaction between these two variants. However, this interaction is not 'real' in the sense that it can be equivalently accounted for by a single variant, $x_{2i}$, and results from the particular genealogical history of this sample. Note that this false-positive signal of interaction is enabled by the close pattern of LD between the three variants, each of which is correlated with the other two.

The potential for an unobserved third variant with a purely additive effect on the phenotype to lead to the spurious inference of non-existent interactions has been previously explored in the literature, where it is known as 'phantom epistasis' [19]. Far from being a merely theorical possibility, this phenomenon has in fact led to the erroneous report of false-positive interactions affecting gene expression levels [20–22]. As discussed in ref. [19], and in line with the toy example above, phantom epistasis is more likely to be an issue when the loci being tested for interaction are in LD. In designing our interaction testing pipeline, therefore, we will ensure that each SNP and PGS are uncorrelated; we will achieve this by building LOCO PGSs, as briefly hinted at above and further discussed below.

##### 2.4.3 Dominance effects

The concepts of dominance and recessiveness were introduced by Gregor Mendel in reference to discrete phenotypic traits when reporting on his experiments with peas [23] and were later generalised to refer to alleles rather than phenotypes and to apply to quantitative traits as well as discrete ones [24]. In modern usage, an allele is considered dominant if the effect of a single copy of it on a phenotype is larger in magnitude than the additional effect of a second copy [25]. In fact, the second copy of the allele may have no effect at all.

To illustrate two relevant points relating to dominance in the context of modelling epistasis, we write as an example the following linear model of a quantitative phenotype $y_i$:

$$y_i = \alpha\,\mathbb{I}(x_{ij} = 1) + \beta\,\mathbb{I}(x_{ij} = 2) + \boldsymbol{\theta}^\intercal \boldsymbol{x}_{i,-j}, \tag{19}$$

where $x_{ij}$ denotes a variant whose value is 0, 1 or 2 depending on whether 0, 1 or 2 copies of the allele in question are present for individual $i$, $\boldsymbol{x}_{i,-j}$ is a vector of genotypes at locations other than $j$ that have a standard additive effect on the phenotype and $\boldsymbol{\theta}^\intercal$ is a transposed vector of their effects. Based on this simple model, we can define the allele whose number of copies the variable $x_j$ counts as dominant (which makes the alternative allele at that position recessive) if and only if $|\alpha| > |\beta|/2$.

We note first that representing dominance for a genetic variant requires two parameters. This is because the standard assumption of the second copy of an allele having the same effect as the first, which is implicit in linear models that do not consider dominance, does not hold. Moreover, the model in equation (19) could equivalently be written as:

$$y_i = \gamma x_{ij} + \delta\,\mathbb{I}(x_{ij} = 1) + \boldsymbol{\theta}^\intercal \boldsymbol{x}_{i,-j}$$

or as:

$$y_i = \zeta x_{ij} + \eta x_{ij}^2 + \boldsymbol{\theta}^\intercal \boldsymbol{x}_{i,-j}. \tag{20}$$

As long as there are two free parameters available to represent the distinct effects of the first and second copy of the mutated allele, dominance effects can be fully and equivalently captured by these different parameterisations of the model.

Second, we see from equation (20) that dominance may be thought of, and represented as, a *self-interaction.* This is important in the context of this project because it suggests that dominance could potentially be detected by our method to test for interactions. While dominance is certainly worthy of study, it is not the focus of this work and has already been explored in depth for over 1000 phenotypes in the UKB [25]. Because of this, we will treat dominance as a possible confounder and seek to prevent our method from detecting these non-linearities as potential interactions, as described in Section 2.5 below.

##### 2.4.4 Non-linear functional form of the genetics–trait mapping

When introducing the general statistical model in equation (1), we noted that it may be necessary to transform the trait in question for this simple linear model to be appropriate. By this we mean that the most appropriate model for a particular trait may take the following form:

$$y_i = f\Bigg(\sum_{k=1}^{K}\beta_k x_{ik} + \sum_{k=1}^{K}\gamma_k \mathbb{I}(x_{ik}=1) + \sum_{k<k'}\delta_{k,k'}x_{ik}x_{ik'} + \eta a_i + \theta b_i + \sum_{m=1}^{M}\iota_m c_{im}\Bigg) + \varepsilon_i, \quad (21)$$

where $f : \mathbb{R} \to \mathbb{R}$ is a function that non-linearly transforms the linear genetic and environmental effects (e.g., $f$ can be a quadratic, exponential or sigmoid function) to generate the trait on the scale in which it is observed.

In general, there is no biological or mathematical reason to expect genetic variation, even if it can be additively combined, to translate linearly onto the scale of the trait. This is especially true for traits whose scale is in a sense artificial by construction – for example, the body mass index (BMI), which does not measure a directly observable phenotype but rather is a non-linear combination of two different phenotypes.

Such 'global' non-linearities involving all SNPs relevant for a trait are interesting in themselves and have been studied before (see, e.g., Otwinowski et al. [26], who refer to this non-linear mapping as 'global epistasis'). However, they are not our object of focus and, importantly, may produce spurious statistical interactions between smaller sets of SNPs if unaccounted for. For example, if $f$ is quadratic then even in the absence of dominance effects and pairwise interactions (i.e., $\gamma_k = 0$ for all $k$ and $\delta_{k,k'} = 0$ for all $k : k < k'$), and ignoring the effects of covariates, by the Binomial Theorem the expression $\left(\sum_{k=1}^{K}\beta_k x_{ik}\right)^2$ will result in pairwise interaction terms for every possible pair of mutations.

##### 2.4.5 Phenotypic outliers

We conclude this section on sources of type I and II errors by considering the potential effects of phenotypic outliers.

In its simplest form, association testing using genotyping array data (where allele counts take values $0, 1$ or $2$ rather than being an imputed dosage taking values in $[0, 2]$) is a test for the differences in the mean phenotypic value across the three discrete groups of samples defined by the allele count. Since means are notoriously sensitive to outlying values, association tests for the effect of a variant can be affected by observations with extreme phenotypes, which may lead to false-positive associations.

Consider for example a rare variant (with MAF $< 0.001$, for example), as these are usually the most likely to yield false-positive associations. Since the frequency of this mutation is very low, the group of samples who are homozygous for this mutation will also be very small. Because of this, if a few observations in that group happen to have a very low or very high value for their phenotype, this can sufficiently decrease or increase the mean phenotypic value in that group such that the null hypothesis of no association is rejected – while it would not be if the observations in question were removed from the sample. These associations may be spurious because they are driven by a small number of observations, and so their significance is sensitive to departures from modelling assumptions such as normality. This problem is even more severe when testing for the effects of interactions, since the relevant frequency will then be the product of the frequencies of the two individual variants, and this will be very small for two rare variants.

Due to the potential for rare variants to be incorrectly classified as being associated with a phenotype, it is common to filter out variants with very low frequency before undertaking association testing – we also perform such filtering, as discussed in Section 1.4 above. Moreover, it is also standard practice to pre-process observations with extreme phenotypic values to ensure that these do not carry a disproportionate weight in the analysis, a procedure which we will also undertake, as described below.

##### 2.5 Addressing sources of type I and II errors

In Section 2.4, we identified five factors that may reduce our power to identify true interactions or lead to false-positive associations.[17] We list them here in the order in which they will be addressed in our data analysis pipeline, which we describe in detail in Section 2.6:

1. Observations with extreme phenotypic values;
2. Errors in the inferred PGS;
3. LD between variants being tested for interaction;
4. Global non-linearities in the genetics–trait mapping; and
5. Genetic dominance.

We now consider each of these factors in order and describe our strategies to minimise their impact.

###### 2.5.1 Inverse Normal transformation of the phenotype

Outliers are common in real-world data and as such different methods have been developed in the statistics literature to correct for their potentially harmful effects. We use the standard approach in statistical genetics of transforming the phenotype so that its distribution is Normal by applying a rank-based inverse Normal transformation (INT) to the residuals we obtain after regressing out the covariates listed in Section 1.2.

INT consists of ranking the values of the variable in question and mapping those to the quantiles of a standard Normal distribution, thus ensuring that the transformed variable is Normally distributed. More precisely, if $r_i$ is the rank of the value of $y_i$ for subject $i$ of $N$, we can compute a transformed value $y_i'$ as follows [27, 28]:

$$y_i' = \Phi^{-1}\left(\frac{r_i - c}{N - 2c + 1}\right), \quad c \in [0, 1/2],$$

where $\Phi(\cdot)$ is the cumulative density function of the standard Normal distribution.

Common values for $c$ include 3/8, 1/2 and 0, with different values considered unlikely to have an impact on the results of the analysis [27]. In our pipeline, we will use the value 1/2 as this is the default adopted in the `R` package `bestNormalize` [29, 30] that we employ for this purpose.

[17]A sixth factor, the attenuation bias for interactions with only part of the PGS, is unavoidable, as noted above.

##### 2.5.2 An iterative linear regression algorithm for PGS construction

Our approach to testing for interactions relies on first obtaining an inferred PGS with which interactions involving individual mutations are then tested. We now consider the question of how to build a suitable PGS for this application.

In Section 2.4, we explored the consequences of *additive error* in the PGS on the asymptotic behaviour of the OLS estimator of the coefficient of the SNP $\times$ PGS interaction term. While assuming a purely additive error term is a simplification, it nevertheless allowed us to see how errors in the inferred PGS reduce power to detect true interactions by attenuating the inferred coefficient. We begin by considering the following question: in what ways can a PGS be 'wrong' or poorly estimated?

Our goal is to estimate the additive genetic component of the trait given by the term $\sum_{k=1}^{K} \beta_k x_{ik}$ in equation (1) as accurately as possible. We note first that a PGS is defined and computed based on two vectors:

- A vector of the SNPs included in the PGS; and
- A vector of the same length with the coefficients of each of these SNPs.

Each of these vectors is a source of potential error: the SNPs that we choose to include in the PGS will not always be the true causal SNPs (they may only imperfectly tag a causal variant, for example, or even miss it completely) and their effect sizes will be imperfectly estimated. Therefore, when inferring a PGS for interaction testing, our aim will be to minimise both sources of error.

Our approach involves two steps which we first summarise and then explain in greater detail:

1. We will first build an 'initial PGS' which is designed to account for as great a fraction of the SNPs with a direct (i.e., additive) effect on the trait as possible.

   To achieve this, we include in our PGS a large selection of SNPs found to have an effect of some significance on the phenotype in a simple GWAS. In this first stage, the SNPs' effect sizes are those obtained in that initial GWAS;

2. The inclusion of a large number of SNPs in the PGS – many of which will be in close physical proximity with one another – with effects obtained through a (univariate) GWAS will lead to a 'double counting' of the effects of neighbouring variants.

   To address this, in a second stage we employ an iterative algorithm that calibrates the effect sizes of SNPs in close proximity to account for the LD between them. The goal of this algorithm – which is also its stopping condition – is that none of the SNPs in the PGS remain significant after running it. This means that, if we regress out the final PGS from the trait and perform a GWAS on the resulting residuals, none of the PGS SNPs should be genome-wide significant (i.e., have a p-value $\leq 5 \times 10^{-8}$).

Through this two-step approach we are able to obtain a PGS which includes most of the relevant SNPs for the trait in question while also exhausting the additive signal for these SNPs. Exhausting the additive signal can be seen as an attempt to minimise the additive error $\zeta_i$ in the derivation above, thus reducing the attenuation bias caused by this error.

*Training, validation and test sets:*

Our PGS construction method requires, in addition to a main dataset which we will term the 'training set', two other smaller datasets for calibrating parameters (the 'validation set') and testing the final performance of the scores (the 'test set'). We described how we constructed these three separate datasets in Sections 1.3.1 and 1.3.2 above.

*Step 1: Building an initial PGS with lax Clumping and Thresholding:*

To build an initial PGS which includes as many variants with a direct effect on the trait as possible, we use Clumping and Thresholding (C+T) with a very *loose threshold for the correlation between nearby SNPs.* An advantage of employing C+T is that it is an easily interpretable approach: we can justify precisely why certain variants were or were not included in the PGS.

C+T is based on the LD Clumping procedure which we now describe for completeness. Given a set $\mathcal{A}$ of SNPs of interest with corresponding GWAS summary statistics, begin by ordering SNPs in decreasing order of significance. Also initialise an empty set $\mathcal{B}$ to contain the filtered set of SNPs at the end of the procedure. Then, iteratively remove the most significant SNP from $\mathcal{A}$ (denote it $x$) and add it to $\mathcal{B}$; remove also from $\mathcal{A}$ all SNPs within a window of size $2\varpi$ around $x$ whose LD with $x$ is greater than a threshold $\rho$ (these are discarded rather than added to $\mathcal{B}$ as they are considered to be part of the same 'clump' as $x$). Continue in this manner until no SNPs are left in $\mathcal{A}$. The set of filtered or 'clumped' SNPs is $\mathcal{B}$.

To obtain a PGS through C+T for a given p-value threshold $\tau$ and given a set of SNPs with summary statistics, begin by removing those SNPs with a p-value greater than $\tau$ and then apply LD Clumping to that subset. This gives the set of SNPs in the PGS; their coefficients are simply those obtained in the GWAS.

For each trait, we first run a GWAS in the *training set* to obtain estimated effect sizes and p-values for each SNP. We then run C+T with a threshold $\rho = 0.9$ for the squared correlation between nearby SNPs and a window size of 1 Mb (i.e., $\varpi = 500$ kb) for 1000 different p-value thresholds $\tau$ equally spaced on the logarithmic scale between $10^{-9}$ and 0.05. For each value of the p-value threshold, we compute the PGS/estimated phenotype in the *validation set*, thus obtaining 1000 PGSs.

We evaluate the performance (measured through the coefficient of determination from a linear regression of the trait – after regressing out covariates as we will describe in Section 2.6.1 – on the PGS in the *validation set*) of the PGS resulting from each of the 1000 p-value thresholds and select the best-performing threshold.

Finally, we compute this optimal 'initial PGS' (using the coefficients from the original GWAS as before) for the full set of samples (training, validation and test sets). This gives us a starting set of SNPs and associated coefficients which we will improve upon through the iterative approach we now describe.

*Step 2: Calibrating the coefficients of neighbouring SNPs with an iterative algorithm:*

C+T achieves our first goal of obtaining a PGS with a large selection of relevant SNPs. However, since it includes many SNPs in close proximity and high LD with one another (due to the loose $r^2$ threshold used for LD clumping) and whose coefficients (obtained through univariate linear regressions) do not account for these correlation patterns, it suffers from significant bias due to double counting of neighbouring effects, and this leads to suboptimal performance.

To correct for this, we propose an iterative algorithm that adjusts the effect sizes of each SNP in the PGS *until their additive signal is fully accounted for.* By this we mean that, after running the algorithm, if we regress the PGS out from the trait and perform a GWAS on the PGS SNPs, none of them should be genome-wide significant.

The iterative algorithm is formally described below (Algorithm 2.1) and we now summarise it more informally. We begin by regressing out the initial PGS from the trait and obtaining the resulting residuals – intuitively, these represent the signal in the trait that remains to be explained. We then take these residuals, run a GWAS *only on the SNPs included in the PGS* and build a new score through C+T as above but now with a much more stringent LD threshold of $\rho = 0.1$ so as to only update approximately independent SNPs. We add this new PGS to the existing one (their relative weights are allowed to differ and are computed based on a joint regression of the original phenotype on the current PGS and the new PGS in the training set), regress out this combined PGS from the phenotype and run a new GWAS on the resulting residuals. We continue iterating in this manner until no significant SNPs are left. (Note that the set of SNPs included in the PGS is fixed from the start by the C+T algorithm with which we build the initial PGS – only the coefficients of the included SNPs are updated by the iterative algorithm.)

**Algorithm 2.1:** Iterative PGS algorithm (inputs, outputs and parameters only).

**Input:**

- Initial PGS:
  - Array $\mathcal{L} = (1, \ldots, L)$ of SNPs that comprise the PGS;
  - Initial effect sizes $\hat{\beta}_l^0$ and physical positions $q_l$ for each SNP $l \in \mathcal{L}$;
  - A measure $r^2_{l,l'}$ of LD between neighbouring pairs of SNPs $(l, l') : |q_l - q_{l'}| \leq \varpi$;
  - Genotypes $x_{il}$ for each SNP and each observation $i \in 1, \ldots, N$;
  - Array of initial estimated phenotypic values $\hat{S}_i^0$ for each individual $i$;
- Phenotype $y_i$ for each observation (from which covariates have been regressed out);
- A partition of observations $i \in 1, \ldots, N$ into separate *training* and *validation* sets.

**Parameters:**

- LD Clumping parameters $\varpi$ and $\rho$;
- An array $\mathcal{T} = (\tau_1, \ldots, \tau_T)$ of p-value thresholds for C+T algorithm;
- A threshold of significance $\upsilon$ for termination of the iterative algorithm.

**Output:**

- Updated effect sizes $\hat{\beta}_l^R$ for each SNP after $R$ iterations;
- Array of final estimated phenotypic values $\hat{S}_i^R$ for each individual $i$.

This algorithm is a slightly modified version of the simple boosting algorithm [31, 32] known as Least-Squares Boosting or LS-Boost [33], which in turn is equivalent to gradient descent boosting on a squared-error (or $L_2$) loss function with a linear model as the 'base learner'. LS-Boost differs

**Algorithm 2.1:** Iterative PGS algorithm.

*Initialisation*

$r \leftarrow 0$; `// start iteration counter`

Regress original trait $y_i$ on the initial PGS $\hat{S}_i^0$ in the *training set* and obtain the resulting residuals $\hat{u}_i^0$:

$$y_i = \hat{\alpha}^0 + \hat{\pi}^0 \hat{S}_i^0 + \hat{u}_i^0.$$

Run a GWAS of the residuals $\hat{u}_i^0$ in the *training set* for each SNP $l \in \mathcal{L}$ and obtain coefficients $\hat{\delta}_l^0$ and corresponding p-values $p_l^0$:

$$\hat{u}_i^0 = \hat{\gamma}^0 + \hat{\delta}_l^0 x_{il} + \hat{\varepsilon}_i^0.$$

*Iterative procedure*

**while** $\min_{l \in \mathcal{L}} p_l^r \leq \upsilon$ **do** `// while there are genome-wide sign. SNPs in latest GWAS`

$r \leftarrow r + 1$; `// update iteration counter`

**for** $\tau_t : \tau_t \leq \tau^{r-1}$ **do** `// build intermediate PGS`

Build *temporary 'PGS component'* $\hat{S}_i^{\prime r, \tau_t}$ by applying C+T with p-value threshold $\tau_t$.[a] The p-values and coefficients used for sorting SNPs and building the PGS are those from the most recent GWAS ($p_l^{r-1}$ and $\hat{\delta}_l^{r-1}$, respectively).

Run the following regression in the *training set* to obtain relative weights for the previous PGS and the new temporary PGS component:

$$y_i = \hat{\eta}^r + \hat{\phi}^r \hat{S}_i^{r-1} + \hat{\psi}^r \hat{S}_i^{\prime r, \tau_t} + \hat{\xi}_i^r.$$

Compute an updated PGS as a weighted sum of the previous PGS and the new component and assess its performance in the *validation set*:

$$\hat{S}_i^{r, \tau_t} = \hat{\phi}^r \hat{S}_i^{r-1} + \hat{\psi}^r \hat{S}_i^{\prime r, \tau_t}.$$

**end**

The *new PGS* is $\hat{S}_i^r = \hat{S}_i^{r, \tau^r}$ for the best-performing p-value threshold $\tau^r$. Compute and store also the new coefficients $\hat{\beta}_l^r = \hat{\phi}^r \hat{\beta}_l^{r-1} + \hat{\psi}^r \hat{\delta}_l^{r-1}$, where the weights $\hat{\phi}^r$ and $\hat{\psi}^r$ correspond to the optimal threshold and we set $\hat{\delta}_l^{r-1} = 0$ for the SNPs that do not survive LD clumping at this threshold.

Regress out the new PGS $\hat{S}_i^r$ from the trait in the *training set* ($y_i = \hat{\alpha}^r + \hat{\pi}^r \hat{S}_i^r + \hat{u}_i^r$) and obtain the resulting residuals $\hat{u}_i^r$.

Run a GWAS of the residuals $\hat{u}_i^r$ in the *training set* ($\hat{u}_i^r = \hat{\gamma}^r + \hat{\delta}_l^r x_{il} + \hat{\varepsilon}_i^r$) and obtain coefficients $\hat{\delta}_l^r$ and p-values $p_l^r$.

**end**

**return**

- *Updated effect sizes $\hat{\beta}_l^R$ for each SNP where $R$ is the final iteration;*
- *Array of final estimated phenotypic values $\hat{S}_i^R$ for each individual $i$.*

[a] Note that we force the p-value threshold to stay constant or decrease with every iteration to increase computational speed as the algorithm progresses.

from the algorithm above in two main ways: it begins with all coefficients equal to zero (while we take the initial PGS as the starting point); and it updates only the most significant variable (strictly speaking, the variable that achieves the greatest reduction in squared error) at each step rather than several variables simultaneously as we do.

Despite its simplicity, LS-Boost has appealing theoretical properties: Freund et al. [34] have recently shown that this algorithm converges linearly to the multiple regression OLS solution. This property is appealing in our context as it would guarantee that our algorithm would converge in a finite number of steps and that it would indeed exhaust all additive signal (since this is true by construction for multiple regression solutions as they ensure $\mathbb{E}(x_i\hat{\varepsilon}_i) = 0$ for every regressor $x$, where $\hat{\varepsilon}_i$ are the residuals).

Since our algorithm is slightly different, this theoretical property may not hold. However, our algorithm does converge in practice for all traits (real and simulated) for which we have run it and, since we only update variants that are not in high LD with one another simultaneously, this multiple updating seems unlikely to disrupt convergence (these modifications increase speed considerably in our context with a very large number of SNPs).

More practically, our boosting algorithm achieves effective 'shrinkage' of SNP effects (the final coefficient estimates are 5.8% of their starting values on average across all SNPs and phenotypes), thus performing implicit regularisation while accounting for the correlation of effects of neighbouring SNPs. The performance (measured as the coefficient of determination of a regression of the trait after regressing out covariates on this score in the held-out test set) for each phenotype is reported in Table S1 together with SNP heritability estimates for comparison.

The iterative algorithm we have described achieves our goal of exhausting all the additive genome-wide significant signal for a trait from the large set of SNPs included in the PGS, which in turn allows us to focus on detecting remaining *non-additive* signal for the trait by testing for interactions with this PGS.

###### 2.5.3 Leave-one-chromosome-out PGS to counter LD-driven confounding

To avoid potential confounding caused by LD between the SNP being tested for an interaction with the PGS and neighbouring SNPs in that PGS, after building a PGS we also compute 22 LOCO versions of that score that exclude SNPs from each chromosome in turn. Then, when fitting models like the one in equation (2), for each SNP we use the corresponding LOCO PGS (i.e., the PGS from which all SNPs in the same chromosome as the SNP being tested for an interaction have been removed).

###### 2.5.4 Data-driven adjustment of the functional form of the PGS

We noted previously that our goal is to detect interactions between specific pairs or larger groups of SNPs rather than global non-linearities where the combined additive effect of all relevant SNPs is transformed by some function $f$ to yield the final phenotype (equation (21)).

To account for a possible global transformation of this kind, after obtaining an estimate of the PGS we will transform it by a function $\hat{f}$ with which we aim to approximate $f$. We perform this transformation as follows:

- We begin by sorting all individuals in our training set by their PGS in increasing order;

- We then divide them into bins of 1000 individuals. (To increase granularity at the ends of the PGS distribution, the first and last 1000 individuals are allocated into 20 bins of 50 samples;)
- For each bin, we compute the median of the PGS and the mean of the true phenotype (after regressing out covariates and applying INT) for the individuals in that bin;
- Finally, we perform linear interpolation of these values (with the X and Y coordinates given by the median PGS and average phenotype in each bin, respectively) and replace each individual's PGS with the interpolated value of the average phenotype for their PGS.

The aim of applying this transformation is to account for global non-linear genetic effects on the phenotype through the PGS, and so reduce the likelihood of detecting false-positive interactions driven by such effects, as discussed in Section 2.4.4. In the simulation study that we describe in the next chapter, we will generate phenotypes with global non-linear effects (by applying sigmoidal transformations) and show that our method is robust to this potential source of spurious interactions.

##### 2.5.5 Testing whether dominance explains interaction signals

Our primary interaction testing model, given in equation (2), does not include a dominance term for the SNP being tested. While in principle we could have included such a term, in practice this would increase the computational cost of fitting the model and, more importantly, for rare variants it would lead to a nearly perfect correlation between the allele count $x_{ij}$ and its square, further complicating model fitting.

As an alternative, we test whether an interaction can be explained by a dominance term after we have obtained initial summary statistics for the SNP $\times$ PGS interaction model. We will include dominance terms as we filter these first results down to an independent set of loci as described below in Section 2.6.5.

#### 2.6 Data analysis pipeline

In this final section covering the SNP$\times$PGS interaction GWAS, we describe in more detail the computational implementation of our approach, including the main software packages used. We implemented our data analysis pipeline as a Snakemake workflow [35], with simple statistical operations undertaken in R [36] and most analyses of genetic data run with PLINK 2.0 [37, 38]. All analyses were performed on the Oxford Biomedical Research Computing Cluster (BMRC).

We provide a diagram of the pipeline in Fig. S1 and now proceed to briefly describe the different steps involved.

##### 2.6.1 Pre-processing of traits

Our main regression model includes a large number of covariates previously listed in Section 1.2. Because of this, fitting the model exactly as described in equation (2) would substantially increase the computational cost of the interaction GWAS. Therefore, and as is common practice in

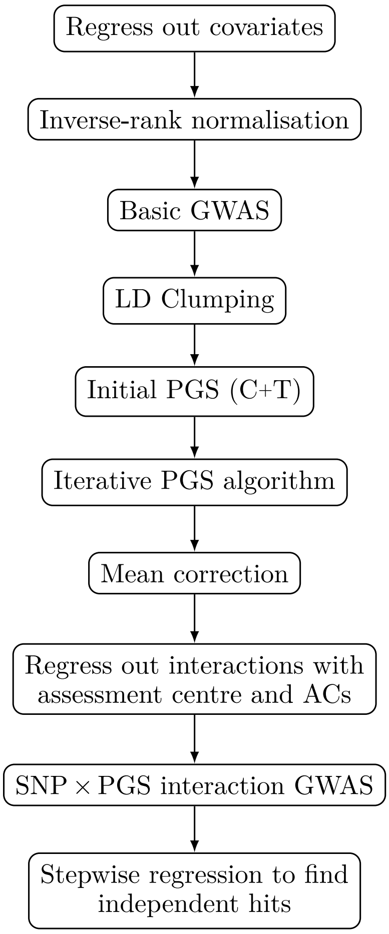

**Figure S1:** Diagram of first part of data analysis pipeline.

the field, we begin by first regressing these out from the trait and then proceed to analyse the resulting residuals.[18]

After regressing out the covariates from each trait, we transform the resulting residuals by applying a rank-based inverse Normal transformation in R using the `orderNorm` function of the `bestNormalize` [29, 30] package with default settings.

##### 2.6.2 GWAS

We continue by running a basic GWAS, i.e., we regress the residuals obtained in the previous step on each SNP in turn, without including any interaction terms or PGSs in these regressions. This first GWAS provides the summary statistics used to build the initial PGS in the next step.

The UKB imputed genetic data is provided in BGEN format [39]. We chose to use PLINK 2.0 [37, 38] for operations involving genetic data whenever possible due to its versatility, stability and speed. Since PLINK cannot directly operate on BGEN files, we first convert the imputed

[18] By the Frisch-Waugh-Lovell Theorem (see, e.g., [18, Section 3.3]), this procedure is equivalent to fitting the larger model with all covariates as long as these are uncorrelated with the explanatory variables of interest – in our case, the allele count of the SNP, the PGS and their product.

genetic data from the original BGEN format into PGEN, PLINK 2.0's native format. The basic GWAS is then run using PLINK 2.0's `--glm` function.

##### 2.6.3 Building the PGS

To build the PGS, we begin by running C+T on the results from the basic GWAS. Since an LD Clumping function had not yet been implemented in PLINK 2.0 at the time of this part of the analysis, we instead use the `bed_clumping` function in the R package `bigsnpr` [40], which can work directly on the original BGEN files.

We run LD Clumping with two alternative thresholds $\rho \in \{0.1, 0.9\}$ for the squared correlation between neighbouring SNPs. The results for $\rho = 0.9$ are used as the starting point for the iterative algorithm as described in Section 2.5.2 while the results for $\rho = 0.1$ give us a set of approximately independent SNPs within each PGS which we will use when incorporating information on transcription factor binding sites below.

Having obtained the LD Clumping results, we then build a series of PGSs on the *validation set* for 1000 different p-value thresholds $\tau$ equally spaced on the logarithmic scale between $10^{-9}$ and 0.05 using PLINK's `--score` function, assess their performance on this sample set and, finally, extend the best-performing score to all samples as described above.

We proceeded by running the second (iterative) step in our PGS construction algorithm. Since this consists of multiple iterative steps each involving GWAS, C+T and intermediate PGS construction, we found PLINK was not fast enough and use the `bigsnpr` and `bigstatsr` R packages [40] instead.

##### 2.6.4 Testing for interactions

To test for interactions between each SNP and the PGS for a trait, we again use PLINK's `--glm` function to fit linear regressions of the phenotype residuals on an intercept, the minor allele count (or dosage) for the SNP, the corresponding LOCO PGS and an interaction term between the SNP and this PGS.

The residuals used in this stage are different from those used for the basic GWAS: from those first residuals (to which INT has been applied) we also regress out interaction terms between the (full, i.e., not LOCO) PGS and each of the assessment centre and ancestry component variables. This is to account for any potential generalised interaction effects between an individual's ancestry (measured directly through the ACs and also correlated with the assessment centre where their data was collected, since these tend to be located in the region where they reside) and their genome-wide additive effects for a trait as aggregated in the PGS.

##### 2.6.5 Identifying independent interaction hits

GWASs often identify association peaks consisting of tens of closely linked SNPs in a single region of the genome that is usually not broken by recombination. It is generally accepted that only one or a few of these SNPs will have a 'causal' effect on the trait (meaning that there is a biological mechanism – such as disruption of a protein or regulation of gene expression, for example – linking that mutation to the phenotype, rather than the SNP simply being highly correlated with one other such SNP) and many methods have been developed that attempt to distil or 'fine-map' these sets of associations into smaller groups of likely causal variants (see refs. [41, 42] for reviews of existing methods).

After obtaining summary statistics from our interaction GWAS, we therefore filter these down to a set of independent associations for each trait. A first use of this filtering is to provide us with a smaller list of associations to interpret and analyse, as well as with an estimate of the number of independent associated loci which is not upwardly biased by LD as described in the previous paragraph. A second benefit of this approach in our setting is that it prioritises specific SNPs for further investigation: below we will describe how we search for subsets of SNPs within the PGS that drive each observed interaction.

When performing fine-mapping in a standard GWAS, it can be helpful to use functional information on associated SNPs to prioritise those which are likely to be causal, possibly integrating these formally into a statistical fine-mapping approach. However, our priority is to increase power to detect loci in the PGS driving the interaction in a subsequent step. Because of this, we did not use functional information or more sophisticated fine-mapping methods to filter loci with statistical evidence of an interaction but rather chose the most significantly associated SNP at each locus as these have the strongest association signal.

In more detail, we employ a stepwise regression procedure of variable selection consisting of the following steps:

1. Sort all genome-wide significant SNPs in decreasing order of significance. Denote this initial set of SNPs by $\mathcal{A}$ and initialise an empty set $\mathcal{B}$ of independent hits;
2. Beginning with the first (most significant) SNP in $\mathcal{A}$, run a regression of the residuals on an intercept, the SNP's minor allele count, its square (to account for a possible dominance effect), the PGS and an interaction between the SNP and the PGS. Remove this SNP from the set $\mathcal{A}$;
3. If the interaction term is genome-wide significant, assign the SNP to the set of independent interaction hits $\mathcal{B}$ and include the three terms related to it (its minor allele count and square, and its interaction with the PGS) in all downstream regression models;[19]
4. Iteratively add to the regression model the remaining SNPs in $\mathcal{A}$ one-by-one and remove them from this set (for each SNP, add a main effect, its square and its interaction with the PGS as before).[20] Those with significant interactions are assigned to $\mathcal{B}$ and remain in the model, while the rest are discarded. Continue until $\mathcal{A}$ is empty.

##### 2.7 Identifying the targets of interaction signals (I): pairwise interactions

The method described above allows us to identify a set of independent interaction signals between a SNP and a PGS for a variety of phenotypes. As previously discussed, identifying interactions involving groups of SNPs rather than only pairs is the primary goal of this project as these can have a particular biological interpretation. However, the PGSs we build contain large numbers of SNPs (indeed, up to a few hundred thousand for highly polygenic traits like standing height) and therefore it may be the case that a SNP×PGS interaction is in fact capturing the modulatory

[19]Note that, once a SNP is added to $\mathcal{B}$, it stays in that set and is not removed at any point in the procedure even if it ceases to be significant once more SNPs are added.

[20]As we consider each remaining SNP in turn, we discard those whose squared correlation with a SNP in $\mathcal{B}$ is strictly greater than 0.9 to increase computational speed.

effect of the SNP on only a small subset of SNPs in the PGS. As the final step in our statistical pipeline, we therefore attempt to identify narrower interaction sets for each independent hit and corresponding phenotype.

We begin with the low-hanging fruit of pairwise interactions. Even though these are not the target of our approach, having identified SNPs that appear to interact with a substantial fraction of the additive genetic component for a trait as represented in a PGS, we are well placed to attempt to find pairwise interactions involving these independent SNP×PGS hits. By restricting ourselves to considering SNP×PGS hits only as the 'left-hand side' SNP in our search for SNP interaction pairs, we narrow down the enormous space of possible pairwise interactions to a very significant degree and are thus able to identify novel interaction pairs without incurring a large penalty for multiple testing.

For most cases, we do not find genome-wide significant 'partners' for the independent SNP×PGS hits through this analysis. This is as expected: it is likely that some SNPs interacting with the PGS do in fact interact with a large group of other SNPs. For such SNPs, it may not be possible – and neither would it be sensible – to reduce the interaction to a pairwise one, or to one involving a small set of SNPs. However, several independent hits can be indeed be linked to specific SNPs in the PGS in this manner, and so we are able to extend the known set of pairwise interactions for human complex traits with a similar computational and multiple testing cost to that of a standard additive GWAS.

We therefore run, for each independent SNP×PGS hit identified in the previous step, a simple GWAS of pairwise interaction with every SNP in the genome (and not only those in the PGS) one-by-one, using the usual genome-wide significance threshold when searching these results for significant associations.[21] We then use stepwise regression again to filter these down to a final set of pairwise interactions.

The stepwise regression procedure used here is analogous to that employed above when filtering SNP×PGS interactions, with a few differences as follows: for each SNP (including both the SNP×PGS hit and each of its interaction pairs), a squared genotype term is only included if the correlation between the genotype and its square is strictly less than 0.999; and, even if the SNP most recently added to the model does not have a significant interaction with the focal SNP×PGS interaction hit, if its main effect is significant we keep it (and its square if available) in downstream regression models.

##### 2.8 Identifying the targets of interaction signals (II): partitioning the PGS

Finally, having considered pairwise interactions involving the independent SNP×PGS hits, we will attempt to partition the PGS into functionally meaningful components determined by transcription factor (TF) binding sites, as well as functional annotations and methylation marks, to identify interactions with these smaller sets of SNPs.

For this final analysis, motivated by computational efficiency as well as conceptual simplicity, we use a simpler PGS than the one employed previously to identify SNP×PGS interactions:

[21]The phenotype used for this GWAS is the same as that used in the basic GWAS, i.e., the residuals obtained from regressing out covariates from the original phenotype and then applying INT. We also include in these regression models the square of the SNP×PGS hit's minor allele count/dosage if the correlation between this count/dosage and its square is strictly less than 0.999.

instead of the 'iterative' PGS computed above, we use a simple C+T PGS with an $r^2$ threshold of 10%, a window size of 1 Mb and a p-value threshold chosen from among 1000 values equally spaced between $1 \times 10^{-9}$ and 0.05 on the logarithmic scale based on performance on the validation set. A LOCO version of this PGS will be used in regression models as explained below, while the set of SNPs that result from this clumping procedure and which are genome-wide significant forms the basis for the TF-specific PGSs.

##### 2.8.1 Making a set of independent GWAS hits and their tags

We begin with the set of genome-wide significant 'hit SNPs' from the C+T PGS. To account for the fact that the most strongly associated SNP in an LD block may not be the 'causal' SNP, we extend this set of hit SNPs to encompass their tags ($r^2 \geq 0.75$ within a window of 1 Mb in the White British samples of the UKB imputed data).[22] By modelling functional and TF-binding information not only for the hit SNPs but also for their tags (with appropriate weighting as described below), we aim to achieve more accurate TF-specific PGSs and so increase our power.

Having found each hit SNP's tags, we use the summary statistics from our basic GWAS to compute an approximate Bayes Factor using Jon Wakefield's approach [43]. For a SNP $k$ with Wald test statistic $z_k$ from a logistic regression model, the Bayes Factor measuring evidence in favour of the *alternative* hypothesis (of non-zero association) can be approximated as follows:[23]

$$\mathrm{BF}_k \approx \frac{1}{\sqrt{1+K}} \exp\left(\frac{z_k^2}{2}\frac{K}{1+K}\right),$$

where $K$ is a scaling factor for the prior distribution for the Bayes Factor. We set $K = 1$, which corresponds to the unit-information prior [43, 44].

Finally, for each independent hit SNP, we divide the tags' Bayes Factors by the sum of Bayes Factors for all tags of that hit SNP so that they sum to one. This rescaled Bayes Factor can be seen as a posterior probability of causality for each tag SNP under the assumption that exactly one of those SNPs is causal for each hit SNP (see, e.g., ref. [41]).

##### 2.8.2 Making a 'sub-PGS' for SNPs with relevant functional consequences or which overlap methylation marks

As an intermediate step to obtaining TF-specific 'sub-PGSs' (TF-PGSs), we first consider annotations of functional consequence and overlap with methylation (H3K4me1 and H3K4me3) marks. We build sub-PGSs for 'functionally significant' and H3K4me1/H3K4me3-overlapping SNPs (which are of interest in themselves and which we will test for interactions) and in the process obtain relevant parameter estimates for the construction of TF-PGSs, as will become clear below.

We begin by making indicator variables of whether each tag SNP has functional relevance and whether it falls in H3K4me1 or H3K4me3 methylation marks in an appropriate cell type:

1. For each tag, check whether it physically overlaps H3K4me1 and H3k4me3 marks separately for each of a range of cell types using data from the Blueprint ChIP-seq Consortium

[22] If a SNP has more than 100 tags, we keep the strongest 100.

[23] This approximation is derived in ref. [43] for binary phenotypes but is likely to hold also for continuous ones.

obtained through the International Human Epigenome Consortium Data Portal [45]. There are 264/306 datasets for H3K4me1/H3K4me3, respectively.

2. For each tag, obtain a functional consequence ('coding') annotation using the Ensembl VEP [8] dataset of whether it belongs to any of the top 24 variant consequence categories ranked by severity.[24]

To compute PGSs specific to these three types of annotation, we first restrict the set of tag SNPs to those with the annotation in question and then multiply their original coefficients (taken from a simple GWAS) by weights that incorporate this annotation. The weights for each tag $t$ in the set $\mathcal{T}_d$ of all tags for driver SNP $d$ are defined as follows:

$$w_{d,t} = \left[1 + a^c_{d,t}(\alpha^*_c - 1)\right] \left[1 + a^{h_1}_{d,t}(\alpha^*_{h_1} - 1)\right] \left[1 + a^{h_3}_{d,t}(\alpha^*_{h_3} - 1)\right], \tag{22}$$

where the indices $c, h_1, h_3$ refer to the coding, H3K4me1 and H3K4me3 annotations, respectively, and $\alpha^*_c, \alpha^*_{h_1}, \alpha^*_{h_3}$ are parameters representing how each type of annotation either increases or decreases the weights in the PGS for the SNPs overlapping that annotation.

To see this more clearly, we consider the first term, which corresponds to 'coding' annotations, and note that 1 is the baseline weight for a tag before taking into account functional information. If the annotation is not present, such that $a^c_{d,t} = 0$, it does not affect the weight for the tag, which remains 1. Conversely, if it is present, the expression simplifies to $\alpha^c_{d,t}$, and therefore the annotation increases the tag's weight in the PGS if $\alpha^*_c > 1$ and decreases it otherwise. Therefore, an annotation may inform the PGS by either increasing or decreasing the weights of SNPs with that annotation, and we allow the three annotations modelled in this way to operate multiplicatively.

We now consider the model under which $\alpha^*_c, \alpha^*_{h_1}, \alpha^*_{h_3}$ are defined and how these parameters are estimated. The object of modelling is which SNPs are causal for this trait; we will construct a model such that this depends on $\alpha$ and then estimate this parameter through maximum likelihood estimation (MLE).

As discussed above, we have a set of regions defined by a 'driver' SNP $d$, each containing a set of tags $\mathcal{T}_d$. We postulate that the causal SNPs for this phenotype are chosen by drawing a number of SNPs equal to the total number of driver SNPs at random, with replacement, from among the set of all tags and drivers in all regions. The probability according to which each tag is drawn is given by a 'prior' which we set to be:

$$\Pr(t \text{ is causal}) = \frac{v_{d,t}}{\sum_{d \in \mathcal{D}} \sum_{t \in \mathcal{T}_d} v_{d,t}},$$

where

$$v_{d,t} = 1 + a_{d,t}(\alpha - 1).$$

[24] As of this writing, these are (in decreasing order of severity): transcript ablation; splice acceptor variant; splice donor variant; stop gained; frameshift variant; stop lost; start lost; transcript amplification; feature elongation; feature truncation; inframe insertion; inframe deletion; missense variant; protein altering variant; splice donor 5th base variant; splice region variant; splice donor region variant; splice polypyrimidine tract variant; incomplete terminal codon variant; start retained variant; stop retained variant; synonymous variant; coding sequence variant; and mature miRNA variant.

Each tag's probability of being chosen as a causal SNP is therefore proportional to a weight that depends on whether it has the annotation in question or not, as discussed above, and on the parameter $\alpha$. Note also that each tag SNP's prior is proportional not only to its weight $v_{d,t}$ relative to the other SNPs tagging its driver but to all tag SNPs for all drivers, so that we sum over all $d$ and all $t \in \mathcal{T}_d$.

The 'data' whose likelihood we maximise under this model is the observed values of the phenotype, which we denote as a vector $\boldsymbol{y}$. We write the likelihood as follows, which corresponds to the product of likelihoods when considering one region at a time (we assume the regions are independent and will ensure that this is the case in practice as we explain below):

$$\mathcal{L}(\alpha) = \prod_{d \in \mathcal{D}} \sum_{t \in \mathcal{T}_d} \Pr(\boldsymbol{y} | t \text{ is causal}) \Pr(t \text{ is causal}).$$

By summing over the tags in each region, we account for the fact that we don't know which is causal.

The formula above can be simplified in our setting by noting that the Bayes Factor for a tag SNP $t$ is, by definition:

$$\mathrm{BF}_t = \frac{\Pr(\boldsymbol{y} | t \text{ is causal})}{\Pr(\boldsymbol{y} | t \text{ is not causal})}.$$

If $t$ is not causal, $\boldsymbol{y}$ does not depend on $t$ and therefore it also does not depend on $\alpha$. This implies that we can replace $\Pr(\boldsymbol{y}|t \text{ is causal})$ in the likelihood by $\mathrm{BF}_t$, as it is proportional to it.

Applying this substitution, inserting the expression for $\Pr(t \text{ is causal})$ above and taking the logarithm, we obtain the following:

$$\log\left(\mathcal{L}(\alpha)\right) \propto \sum_{d \in \mathcal{D}} \log\left( \sum_{t \in \mathcal{T}_d} \frac{v_{d,t}}{\sum_{d \in \mathcal{D}} \sum_{t \in \mathcal{T}_d} v_{d,t}} \mathrm{BF}_{d,t} \right). \tag{23}$$

To implement this in practice, we begin by ensuring that the regions defined by the drivers and their tags are independent:

3. Some tags may be assigned to multiple hit SNPs, creating dependence between different regions/LD blocks. To achieve a set of independent regions, assign tags of multiple hit SNPs to the hit SNP with the smallest number of tags. Then, keep only regions with no evidence of overlap with another region: if, for a region, the reassigned tags accounted for more than 5% of the sum of initial probabilities (which is 1 by construction), consider this as evidence of overlap and remove that region.

We then fit the model above separately for the three types of annotation, noting that we have a large number of datasets of H3K4me1/H3K4me3 methylation which for the moment we process as separate annotations:

4. For each annotation (coding and 264/306 annotations for H3K4me1/H3K4me3), find the value of $\alpha$ from among 79 possible values (from 0.1 to 4 in increments of 0.05) that maximises the log-likelihood in equation (23).

   For H3K4me1 and H3K4me3 separately, find the dataset for which the likelihood is maximised and use that same dataset and corresponding optimal $\alpha$ in all downstream analyses for this phenotype.

We now have everything we need to compute 'sub-PGSs' specific to each annotation:

5. Take the optimal values $\alpha_c^*, \alpha_{h_1}^*, \alpha_{h_3}^*$ for each of the three annotations and compute the weights $w_{d,t}$ for each tag using the formula in equation (22).
6. Assign each tag to a unique driver as before and redistribute the initial Bayes Factors within each driver so that they sum to one. Reweight the Bayes Factors of each tag by the weights computed above, ensuring that they sum to one for each driver at the end.
7. For each of the three annotations, take only the SNPs which have this annotation, sort them by their new weights in decreasing order and keep only those SNPs accounting for the first 80% of the cumulative distribution.
8. Multiply the weight $w_{d,t}$ of each tag that survived the previous step by its GWAS coefficient to obtain the final PGS coefficients.

##### 2.8.3 Finding SNPs that disrupt TF binding sites

To obtain TF-specific PGSs, we extend the approach of the previous section to incorporate information on TF binding sites in the selection of SNPs for the 'sub-PGS' and in the computation of their weights. The information on TF binding sites is obtained from the HOCOMOCO v13 CORE collection database [46], which contains 1611 DNA binding motifs for 1120 unique human TFs in the form of Position Count Matrices (PCMs).

We begin by assuming that we have available for each tag SNP $t$ of driver $d$ a score $\mathrm{OR}_{d,t}$ such that, for a SNP with no possibility of having an 'active' motif for the current TF (not indexed), $\mathrm{OR}_{d,t} = 1$ and, for a SNP with some possibility of having an 'active' motif, $\mathrm{OR}_{d,t} > 1$. Here, 'active' means that the TF binds to this motif and causally generates the association through this binding. We proceed under the assumption that such a score is available and then explain how we compute it.

Previously, we modelled the prior probability of causality for a tag SNP as being proportional to $(1 + a(\alpha - 1))$, where $a$ was a binary indicator of the presence of an annotation and $\alpha$ a parameter fit to data under a model of which tag SNPs are causal. Since determining whether there is an active motif is more complex than simply checking whether a SNP has a given functional annotation or overlaps certain methylation marks, we allow the annotation variable that contains this information to be a quantitative measure rather than a binary indicator and thus now model the prior probability of causality as being proportional to $(1 + (\mathrm{OR}_{d,t} - 1)(\alpha - 1))$, i.e., we redefine $v_{d,t}$ as:

$$v_{d,t} = 1 + (\mathrm{OR}_{d,t} - 1)(\alpha_m - 1), \tag{24}$$

where $m$ indexes the motif of interest. As before, we consider 1 to be the baseline term corresponding to no motif activity while the term $(\mathrm{OR}_{d,t} - 1)(\alpha_m - 1)$ corresponds to it being active. The model of the form $(1 + a(\alpha - 1))$ that we used for functional and methylation annotations above can be seen as a special case of this more general model where we simply take $\mathrm{OR}_{d,t} = 2$ if the annotation is present and $\mathrm{OR}_{d,t} = 1$ otherwise. Then, $\alpha$ is the prior weight on the SNP if the annotation is present and 1 the prior weight if it is not present; this makes the parameterisation in terms of $(\alpha - 1)$ natural for that case and we therefore use it for the general case also.

Note that, for the purposes of fitting $\alpha_m$ by MLE, we only need to specify the prior up to a constant of proportionality not depending on $\alpha_m$. When computing the posterior probability below, we will use as a prior $v_{d,t}$ as defined in equation (24) above multiplied by terms corresponding to functional/methylation annotations so that this additional information is also included in the prior and hence factors into the posterior weights. However, for simplicity we do not include it in the definition of $v_{d,t}$ used to fit $\alpha_m$ as it makes no difference to that inference step.

We also note that, while it may make sense statistically to have $\alpha_m < 1$, this would correspond to reducing the prior probability of causality for the SNP when the motif is present, which is of questionable biological relevance in this context. We will therefore exclude, for each trait, the motifs for which the inferred $\alpha_m$ is less than or equal to 1 (since if $\alpha_m = 1$ the motif carries no information under this model).

Having redefined $v_{d,t}$ in equation (24), we proceed as before and fit, for each motif $m$, the parameter $\alpha_m$ by MLE using the likelihood in equation (23).[25] We then redefine also the final weight variable $w_{d,}$, which now includes the new information on the prior probability of causality coming from the TF as well as the previous contributions from the functional/methylation annotations:

$$w_{d,t} = [1 + (\mathrm{OR}_{d,t} - 1)(\alpha_m - 1)] \left[1 + a^c_{d,t}(\alpha^*_c - 1)\right] \left[1 + a^{h_1}_{d,t}(\alpha^*_{h_1} - 1)\right] \left[1 + a^{h_3}_{d,t}(\alpha^*_{h_3} - 1)\right].$$

In practice, we simplify this expression by using the following adjusted formula:

$$w_{d,t} \approx \left[1 + (\mathrm{OR}_{d,t} - 1)(\alpha_m - 1) + a^c_{d,t}(\alpha^*_c - 1)\right] \left[1 + a^{h_1}_{d,t}(\alpha^*_{h_1} - 1)\right] \left[1 + a^{h_3}_{d,t}(\alpha^*_{h_3} - 1)\right],$$

which provides a slightly smaller boost for motif annotations compared to functional/methylation ones by removing a single cross-term, $a^c_{d,t}(\alpha^*_c - 1)(\mathrm{OR}_{d,t} - 1)(\alpha_m - 1)$ (assuming $\alpha$ values above 1). Biologically, this means we assume coding mutations do not in fact function by disrupting the motif, unless they coincide with H3K4me1 or H3K4me3 annotations (and otherwise, we allow annotations to operate multiplicatively).

Finally, we proceed analogously to steps 6–8 in the previous section. We first assign each tag to a unique driver and redistribute the initial Bayes Factors. Then, we multiply them by $w_{d,t}$ to obtain posterior probabilities of causality for each tag SNP and redistribute them so they sum to one for each driver.

Before, we kept only the tag SNPs with a particular annotation when making a PGS corresponding to that annotation. Since the TF binding annotations we built are quantitative rather than binary, we now multiply the newly obtained posterior probabilities by a quantity approximating the probability that the motif is active (for $\mathrm{OR}_{d,t} \geq 1$), which is conceptually analogous:[26]

$$\frac{(\mathrm{OR}_{d,t} - 1)(\alpha_m - 1)}{1 + (\mathrm{OR}_{d,t} - 1)(\alpha_m - 1)}.$$

[25] Now a greater range of values for $\alpha$ is considered: from 0.1 to 4 in increments of 0.05; and from 4.25 to 6 in increments of 0.25.

[26] This formula follows from the fact that, as discussed above, 1 corresponds to the baseline probability of causality if the motif is not active while $(\mathrm{OR}_{d,t} - 1)(\alpha_m - 1)$ to it being active. Note also that these weights take values between 0 and 1.

We make any negative weights zero, sort them in decreasing order and keep only those SNPs accounting for the first 80% of the cumulative distribution. Multiplying these weights by each surviving tag's GWAS coefficients, we obtain the TF-PGS's final coefficients.

Note that for many TFs there will be no surviving SNPs with a positive weight at the end of this process (we require that more than one such SNP survives, as otherwise this would be equivalent to a pairwise test which we have already undertaken). These are naturally excluded and therefore the list of TF-PGSs tested will be slightly different for each phenotype.

##### 2.8.4 Obtaining TF binding motif scores

Finally, we have not specified how we obtain a score $\mathrm{OR}_{d,t}$ for each tag and motif (not indexed) reflecting how likely the tag is to be located in an active binding site for that motif. In some sense, any approach is valid since we explore whether such an annotation is useful later by MLE. We choose to use properties of the presence or absence of the motif within the set of H3K4me1 peaks (likely enhancers in the relevant tissue type) when compared with regions with neither H3K4me1 nor H3K4me3 peaks to define this score.

For this step, we will make use of an additional external dataset: the results of running the ARG inference method Relate [47] on the 1000 Genomes Project (1kGP) Phase 3 dataset (2013 release) [48, 49] which contain information on which allele is ancestral and which is derived as well as allele frequencies in a sample from the same population that is used in this project ('GBR, British in England and Scotland').[27]

We begin by creating two sample sets of tag SNPs that fall in H3K4me1 marks, or neither in H3K4me1 nor in H3K4me3 marks, as follows:

1. Take the subset of SNPs in the Relate results for the GBR population that are present in our set of tags and obtain their allele count distribution: more specifically, these results show for each SNP the total count of derived alleles in a sample of 90 individuals of the GBR population in the 1kGP data, and we count the number of SNPs with each observed derived allele count.

   Then take the full set of SNPs in the Relate results for the GBR population (not only those in our set of tags as before) and draw a random sample (without replacement) of SNPs falling in H3K4me1 marks (using the 'best' ChIP-Seq dataset identified for the phenotype under analysis in step 4 of Section 2.8.3) so as to obtain the same derived allele count distribution as for the general set of tags that we obtained above. The number of SNPs sampled is equal to the number of tag SNPs in the Relate results.[28]

2. Repeat the previous step for SNPs that fall in neither methylation mark (referred to as the 'neither set' in what follows).

We now use these two sets of tags as approximating regions where a motif can be bound (enhancers) or not (regions that have neither methylation mark). We will score the SNPs in these

[27]We take information on ancestral/derived alleles from the results of running Relate on data from all 1kGP populations jointly (unpublished data). Allele frequency information is taken from the results for GBR samples only provided in ref. [47].

[28]If not enough SNPs are available for a given derived allele frequency bin, we take only those that are (i.e., we do not sample with replacement).

two sets for the binding of each TF (using a standard approach) and compute the odds ratio of SNPs in each of the two sets having a score above a certain percentile (for a number of upper percentiles, as described below) to approximate the empirical distribution of these scores. Lastly, we will use the scores and corresponding odds ratios, comparing them between the H3K4me1 set and the 'neither set', to obtain $\mathrm{OR}_{d,t}$ scores for the full set of tag SNPs by linear interpolation.

We proceed by scoring SNPs in the two sets for the binding of each TF using a moving window approach in a relatively large window around them:

3. For each tag SNP in the sample set of H3K4me1 SNPs, obtain a surrounding sequence of 150 bp (75 bp on either side of the tag SNP) from the reference sequence provided in the UCSC Genome Browser dataset. Then score each sequence for how closely each TF binding motif matches any portion of it:

   (a) For each motif, obtain its PCM, add a fifth column of 1s and replace any 0s with 1s. Compute row-wise frequencies, obtaining a Position Weight Matrix (PWM). The table below shows the original PCM and processed PWM for *ESR2* as an example.

**Table:** Position matrices for *ESR2* (alternative motif).

PCM as obtained from HOCOMOCO

| A | C | G | T |
|---|---|---|---|
| 676 | 53 | 288 | 4 |
| 25 | 1 | 972 | 23 |
| 7 | 10 | 1002 | 2 |
| 17 | 8 | 73 | 923 |
| 3 | 1005 | 2 | 11 |
| 967 | 1 | 51 | 2 |
| 89 | 465 | 361 | 106 |

PWM after processing

| A | C | G | T | N |
|---|---|---|---|---|
| 0.661 | 0.052 | 0.282 | 0.004 | 0.001 |
| 0.024 | 0.001 | 0.951 | 0.023 | 0.001 |
| 0.007 | 0.010 | 0.980 | 0.002 | 0.001 |
| 0.017 | 0.008 | 0.071 | 0.903 | 0.001 |
| 0.003 | 0.983 | 0.002 | 0.011 | 0.001 |
| 0.946 | 0.001 | 0.050 | 0.002 | 0.001 |
| 0.087 | 0.455 | 0.353 | 0.104 | 0.001 |

   (b) Then take a moving window approach, with the window having the same width as the length of the motif. Place the window at the start of the 150 bp sequence and compute a score for that window, then move it one base pair to the right and compute another score for that second window, and so on.

   Indexing the rows of the PWM by $i = 1, \ldots, L$, denoting the elements of each row of the matrix from left to right by $f_{i,A}, f_{i,C}, f_{i,G}, f_{i,T}, f_{i,N}$, and letting the sequence portion to be scored be $s_1, \ldots, s_L$, we compute its score as the product of the frequencies associated with each nucleotide in the sequence in the PWM:

$$\prod_{i=1}^{L} f_{i,A}^{\mathbb{I}(s_i=A)} f_{i,C}^{\mathbb{I}(s_i=C)} f_{i,G}^{\mathbb{I}(s_i=G)} f_{i,T}^{\mathbb{I}(s_i=T)}.$$

   We repeat this process for the complement sequences (i.e., we reverse the order of the sequence and swap each base for its complement). The final score of each 150 bp sequence is then the maximum of the scores between the normal and complement sequences across all the windows.

(c) To obtain a reference scale for these scores, we sample for each motif 10,000 sequences of the same length as the motif where each nucleotide is sampled according to the motif's PWM and score them using the approach above. We then compute the percentiles of these scores, which gives us a baseline distribution of the motif's scores.

   Having obtained this reference distribution, we turn the scores of each motif obtained above into their corresponding percentiles in the reference distribution, thus converting them into an intepretable scale.

4. We repeat the previous steps for the 'neither set' of SNPs.

5. For each motif and for a set of nine top-percentile thresholds $1 - \tau$, with $\tau \in \{0.001, 0.005, 0.01, 0.02, 0.05, 0.1, 0.15, 0.2, 0.25\}$, we count the number of tag SNPs whose score is above/below the threshold for the sample set of SNPs in H3K4me1 marks and in neither mark separately, and compute the odds (number of SNPs strictly above the threshold divided by number of SNPs at or below the threshold). We then compute the ratio of these odds ratios as follows:

$$\frac{\text{odds ratio of having score} > (1-\tau) \text{ in the H3K4me1 set}}{\text{odds ratio of having score} > (1-\tau) \text{ in the `neither set'}}. \tag{25}$$

6. For each motif and for the same set of nine top-percentile thresholds, compute the average of scores across tags that are above the corresponding threshold *in the 'neither set' only.* This set of nine average scores across tags above each threshold in the 'neither set' constitutes the 'x-values' that we will use when performing the interpolation, while the ratios above will be the 'y-values'. Moreover, we impose that the odds ratio be 1 at 0, i.e., if the SNP is a very poor match to the motif.

We are now ready to score the full set of tags, this time using a narrower window around each SNP:

7. For each tag, extract the 60 bp surrounding sequence (30 bp on either side); we chose this sequence length because the longest motif in HOCOMOCO has length 31 and the sequences must be long enough to accommodate windows of each motif's length that begin or end at the tag SNP. Make a reference sequence where the central position only is replaced with the ancestral allele from Relate where available (where not available we take the reference allele), and similarly for the alternative sequence.

8. For each motif, use again a moving window approach (with the width of the window being the length of the motif and where the window always includes the central position in the sequence, i.e., the tag SNP) and compute a score by multiplying the frequencies of each nucleotide taken from the PWM as before.

   Repeat this process for the complement sequences and take the highest score across all windows for normal and complement (for reference and alternative sequences separately). As before, replace the scores with their percentiles in the reference distribution of scores for that motif obtained above.

9. Finally, for each tag and each motif, take the highest score between reference and alternative sequences and *define $OR_{d,t}$ to be the linearly interpolated value of the ratio in equation* (25)

*given this score as the 'x-value'* (where, as mentioned previously, the average scores above the threshold in the 'neither set' from step 6 and the ratio of odds ratios from step 5 for the nine upper-percentile thresholds are used as the 'x' and 'y' interpolation vectors, respectively).

##### 2.8.5 Testing for interactions with TF-specific PGSs

Having computed TF-specific PGSs for a large number of TFs – as well as PGSs for 'coding'/H3K4me1/H3K4me3 SNPs – for the traits for which we identified at least one SNP×PGS interaction hit, we then run interaction tests between these (independent) hits and each sub-PGS. In more detail, for each independent trait–SNP×PGS hit pair, we fit separate regression models for the 'coding'/H3K4me1/H3K4me3 PGSs and all available TF-PGSs where the phenotype is the residuals after regressing out covariates and applying INT (the same used for the basic GWAS and for pairwise interaction testing) and the regressors are: an intercept term; the genotype of the SNP and its square;[29] the standard LOCO PGS (based on C+T with an LD threshold of 10% as mentioned at the beginning of this section) and its interaction with the SNP; and the 'coding'/H3K4me1/H3K4me3 or TF-PGS (which is also a LOCO PGS) and its interaction with the SNP. In including the standard LOCO PGS and its interaction with the SNP in addition to the TF-PGS and its interaction, we aim to only identify TFs whose interaction with the SNP in question is relevant even after the original interaction with the regular PGS has been accounted for.

We then filter these results into a set of independent interacting sub-PGSs by again using a stepwise regression procedure. Analogously to the stepwise procedures previously described, we begin with the same setup as for the GWAS for the most significant sub-PGS and then iteratively add (in decreasing order of significance) the sub-PGSs that showed a significant interaction signal (we add both a main effect and a term for interaction with the SNP), keeping those whose interaction is significant in the model until no significant hits remain. Since the number of tests being performed for each SNP×PGS hit is relatively small (fewer than 750 on average as, for each trait, many TFs are excluded because there are no SNPs with a positive weight after the procedure described in Section 2.8.3), we use $10^{-5}$ as a conservative significance threshold.

##### 2.8.6 Two robustness checks for interactions with TF-specific PGSs

We conclude our data analysis pipeline with two final robustness checks of the independent interactions with TF-specific PGSs[30] identified in the previous section. First, we consider the possibility that an interaction with a TF-PGS may be driven primarily by a pairwise interaction between the SNP in question and one of the driver SNPs in the TF-PGS and/or its tags. To check for this, we check whether each SNP included in the TF-PGS had a significant (at the same $10^{-5}$ level as the TF-PGS interaction) pairwise interaction with the independent interaction hit in the pairwise interaction GWAS and, if so, remove that SNP, its associated driver SNP if it is a tag, and all other tags of this driver SNP. For each TF interaction hit, we repeat the interaction

[29] Only if the correlation between the minor allele count/dosage and its square is strictly less than 0.999 as above.

[30] Or 'coding'/H3K4me1/H3K4me3 sub-PGSs; in this section, we include also these scores when discussing TF-PGSs.

test after removing possible pairwise targets and keep only those hits that remain significant. Of 24 independent TF interactions that we initially identified, this procedure discarded half, leaving the 12 hits reported in Table S14.

Second, since we included the standard LOCO PGS in all regressions when testing for interactions with TF-PGSs, it is conceivable that the part of the standard PGS corresponding to a particular TF does *not* in fact interact with the SNP in question if the two interactions have opposite signs. Indeed, hypothetically, if all the standard PGS SNPs except those corresponding to the TF interact with the SNP, then opposite signs between the standard and TF-PGS interaction terms could mean that the SNPs in the standard PGS corresponding to this TF-PGS were being 'removed' by the TF-PGS interaction, in which case this interaction would be a statistical artefact driven by non-interacting SNPs in the standard PGS.

To test for this, after identifying a set of independent interactions with TF-PGSs we remove for each one the TF-PGS from the standard PGS – so that the two form a partition of the original standard PGS – and rerun the regression. In greater detail, we first note that the set of SNPs in the TF-PGS is unlikely to be a subset of the SNPs in the standard PGS: this is because when building the TF-PGS we start with the set of SNPs in the standard PGS but then expand it to include the tags of those SNPs (which are not part of the standard PGS). To perform this test we therefore begin by making a TF-PGS whose component SNPs are a subset of the standard PGS: we do this by taking the final set of SNPs defining each TF-PGS and replacing the tag SNPs with their corresponding driver SNP and assigning to each driver a weight equal to the sum of those of its tag SNPs. We then recompute the TF-PGS using this new set of driver SNPs and weights and finally subtract it from the standard PGS. Reassuringly, the results of this alternative test are very similar to those of the original procedure (Table S15), suggesting that the issue that motivated this robustness check is not important in our context.

#### 2.A Proof of Proposition 2.2

We begin by plugging in the true model of $y_i$ given in equation (11) into the expression for the OLS estimator given in equation (13):

$$\hat{\alpha}_2 = \frac{\sum_i S_i^2 \sum_i x_{ij} S_i (S_i + \lambda_j x_{ij} R_i + \varepsilon_i)}{\sum_i S_i^2 \sum_i x_{ij}^2 S_i^2 - \left(\sum_i S_i^2 x_{ij}\right)^2} - \frac{\sum_i S_i^2 x_{ij} \sum_i S_i (S_i + \lambda_j x_{ij} R_i + \varepsilon_i)}{\sum_i S_i^2 \sum_i x_{ij}^2 S_i^2 - \left(\sum_i S_i^2 x_{ij}\right)^2}.$$

Replacing $R_i$ with $S_i - Q_i$ following equation (4), we obtain:

$$\begin{aligned}\hat{\alpha}_2 = & \frac{\sum_i S_i^2 \sum_i x_{ij} S_i [S_i + \lambda_j x_{ij} (S_i - Q_i) + \varepsilon_i]}{\sum_i S_i^2 \sum_i x_{ij}^2 S_i^2 - \left(\sum_i S_i^2 x_{ij}\right)^2} \\ & - \frac{\sum_i S_i^2 x_{ij} \sum_i S_i [S_i + \lambda_j x_{ij} (S_i - Q_i) + \varepsilon_i]}{\sum_i S_i^2 \sum_i x_{ij}^2 S_i^2 - \left(\sum_i S_i^2 x_{ij}\right)^2}.\end{aligned}$$

We now isolate $\lambda_j$:

$$\begin{aligned}\hat{\alpha}_2 = \lambda_j & \left[ \frac{\sum_i S_i^2 \sum_i x_{ij}^2 S_i^2 - \sum_i S_i^2 \sum_i x_{ij}^2 S_i Q_i}{\sum_i S_i^2 \sum_i x_{ij}^2 S_i^2 - \left(\sum_i S_i^2 x_{ij}\right)^2} \right. \\ & \left. - \frac{\sum_i S_i^2 x_{ij} \sum_i S_i^2 x_{ij} - \sum_i S_i^2 x_{ij} \sum_i S_i x_{ij} Q_i}{\sum_i S_i^2 \sum_i x_{ij}^2 S_i^2 - \left(\sum_i S_i^2 x_{ij}\right)^2} \right] \\ & + \frac{\sum_i S_i^2 \sum_i x_{ij} S_i (S_i + \varepsilon_i)}{\sum_i S_i^2 \sum_i x_{ij}^2 S_i^2 - \left(\sum_i S_i^2 x_{ij}\right)^2} \\ & - \frac{\sum_i S_i^2 x_{ij} \sum_i S_i (S_i + \varepsilon_i)}{\sum_i S_i^2 \sum_i x_{ij}^2 S_i^2 - \left(\sum_i S_i^2 x_{ij}\right)^2}.\end{aligned}$$

Rearranging the first two fractions and cancelling terms in the second two, we get:

$$\begin{aligned}\hat{\alpha}_2 = \lambda_j & \left[ 1 - \frac{\sum_i S_i^2 \sum_i x_{ij}^2 S_i Q_i - \sum_i S_i^2 x_{ij} \sum_i S_i x_{ij} Q_i}{\sum_i S_i^2 \sum_i x_{ij}^2 S_i^2 - \left(\sum_i S_i^2 x_{ij}\right)^2} \right] \\ & + \frac{\sum_i S_i^2 \sum_i x_{ij} S_i \varepsilon_i - \sum_i S_i^2 x_{ij} \sum_i S_i \varepsilon_i}{\sum_i S_i^2 \sum_i x_{ij}^2 S_i^2 - \left(\sum_i S_i^2 x_{ij}\right)^2}.\end{aligned}$$

We can now use the Weak Law of Large Numbers (since our samples are assumed to be i.i.d.) and Slutsky's Theorem to examine the estimator's behaviour as the sample size becomes large:

$$\begin{aligned}\hat{\alpha}_2 \xrightarrow{p} \lambda_j & \left[ 1 - \frac{\mathbb{E}(S_i^2)\mathbb{E}(x_{ij}^2 S_i Q_i) - \mathbb{E}(S_i^2 x_{ij})\mathbb{E}(S_i x_{ij} Q_i)}{\mathbb{E}(S_i^2)\mathbb{E}(x_{ij}^2 S_i^2) - \left[\mathbb{E}(S_i^2 x_{ij})\right]^2} \right] \\ & + \frac{\mathbb{E}(S_i^2)\mathbb{E}(x_{ij} S_i \varepsilon_i) - \mathbb{E}(S_i^2 x_{ij})\mathbb{E}(S_i \varepsilon_i)}{\mathbb{E}(S_i^2)\mathbb{E}(x_{ij}^2 S_i^2) - \left[\mathbb{E}(S_i^2 x_{ij})\right]^2}.\end{aligned}$$

Since, by assumption, $\varepsilon_i$ is independent of $x_{ij} S_i$ and $S_i$, we have:

$$\mathbb{E}(x_{ij} S_i \varepsilon_i) = \mathbb{E}(S_i \varepsilon_i) = 0,$$

which makes the second term zero.

Moreover, we assumed $\mathbb{E}(x_{ij}^2) = 1$ and that $x_{ij}^2$ is uncorrelated with $S_iQ_i$ and $S_i^2$, which gives:

$$\begin{aligned}\mathbb{E}(x_{ij}^2 S_i Q_i) &= \mathbb{E}(x_{ij}^2)\mathbb{E}(S_iQ_i) = \mathbb{E}(S_iQ_i), \text{ and} \\ \mathbb{E}(x_{ij}^2 S_i^2) &= \mathbb{E}(x_{ij}^2)\mathbb{E}(S_i^2) = \mathbb{E}(S_i^2).\end{aligned}$$

Removing null terms and substituting in these simplified expressions, we obtain:

$$\begin{aligned}\hat{\alpha}_2 &\xrightarrow{\text{p}} \lambda_j \left[1 - \frac{\mathbb{E}(S_i^2)\mathbb{E}(S_iQ_i)}{[\mathbb{E}(S_i^2)]^2}\right] \\ &\xrightarrow{\text{p}} \lambda_j \left[1 - \frac{\mathbb{E}(S_iQ_i)}{\mathbb{E}(S_i^2)}\right].\end{aligned}$$

Finally, since $S_i = R_i + Q_i$ and all variables have mean zero, we can write:

$$\begin{aligned}1 - \frac{\mathbb{E}(S_iQ_i)}{\mathbb{E}(S_i^2)} &= \frac{\mathbb{E}(S_i(S_i - Q_i))}{\mathbb{E}(S_i^2)} \\ &= \frac{\text{Cov}(R_i, S_i)}{\text{Var}(S_i)},\end{aligned}$$

which gives the desired result. □

#### 2.B Proof of Proposition 2.3

We plug the true model of $y_i$ given in equation (14) into the expression for the OLS estimator given in equation (16):

$$\hat{\alpha}_2 = \frac{\sum_i \hat{S}_i^2 \sum_i x_{ij}\hat{S}_i(S_i + \lambda_j x_{ij} S_i + \varepsilon_i)}{\sum_i \hat{S}_i^2 \sum_i x_{ij}^2 \hat{S}_i^2 - \left(\sum_i \hat{S}_i^2 x_{ij}\right)^2} - \frac{\sum_i \hat{S}_i^2 x_{ij} \sum_i \hat{S}_i(S_i + \lambda_j x_{ij} S_i + \varepsilon_i)}{\sum_i \hat{S}_i^2 \sum_i x_{ij}^2 \hat{S}_i^2 - \left(\sum_i \hat{S}_i^2 x_{ij}\right)^2}.$$

Since $S_i$ is now observed with error, we use equation (7) to replace it:

$$\begin{aligned}\hat{\alpha}_2 = &\frac{\sum_i \hat{S}_i^2 \sum_i x_{ij}\hat{S}_i[\hat{S}_i - \zeta_i + \lambda_j x_{ij}(\hat{S}_i - \zeta_i) + \varepsilon_i]}{\sum_i \hat{S}_i^2 \sum_i x_{ij}^2 \hat{S}_i^2 - \left(\sum_i \hat{S}_i^2 x_{ij}\right)^2} \\ &- \frac{\sum_i \hat{S}_i^2 x_{ij} \sum_i \hat{S}_i[\hat{S}_i - \zeta_i + \lambda_j x_{ij}(\hat{S}_i - \zeta_i) + \varepsilon_i]}{\sum_i \hat{S}_i^2 \sum_i x_{ij}^2 \hat{S}_i^2 - \left(\sum_i \hat{S}_i^2 x_{ij}\right)^2}.\end{aligned}$$

Isolating $\lambda_j$:

$$\begin{aligned}\hat{\alpha}_2 = \lambda_j &\left[\frac{\sum_i \hat{S}_i^2 \sum_i x_{ij}^2 \hat{S}_i^2 - \sum_i \hat{S}_i^2 \sum_i x_{ij}^2 \hat{S}_i \zeta_i}{\sum_i \hat{S}_i^2 \sum_i x_{ij}^2 \hat{S}_i^2 - \left(\sum_i \hat{S}_i^2 x_{ij}\right)^2}\right. \\ &\left. - \frac{\sum_i \hat{S}_i^2 x_{ij} \sum_i \hat{S}_i^2 x_{ij} - \sum_i \hat{S}_i^2 x_{ij} \sum_i \hat{S}_i x_{ij} \zeta_i}{\sum_i \hat{S}_i^2 \sum_i x_{ij}^2 \hat{S}_i^2 - \left(\sum_i \hat{S}_i^2 x_{ij}\right)^2}\right] \\ &+ \frac{\sum_i \hat{S}_i^2 \sum_i x_{ij}\hat{S}_i(\hat{S}_i - \zeta_i + \varepsilon_i)}{\sum_i \hat{S}_i^2 \sum_i x_{ij}^2 \hat{S}_i^2 - \left(\sum_i \hat{S}_i^2 x_{ij}\right)^2} \\ &- \frac{\sum_i \hat{S}_i^2 x_{ij} \sum_i \hat{S}_i(\hat{S}_i - \zeta_i + \varepsilon_i)}{\sum_i \hat{S}_i^2 \sum_i x_{ij}^2 \hat{S}_i^2 - \left(\sum_i \hat{S}_i^2 x_{ij}\right)^2}.\end{aligned}$$

Rearranging the first two fractions and cancelling terms in the second two, we get:

$$\hat{\alpha}_2 = \lambda_j \left[ 1 - \frac{\sum_i \hat{S}_i^2 \sum_i x_{ij}^2 \hat{S}_i \zeta_i - \sum_i \hat{S}_i^2 x_{ij} \sum_i \hat{S}_i x_{ij} \zeta_i}{\sum_i \hat{S}_i^2 \sum_i x_{ij}^2 \hat{S}_i^2 - \left(\sum_i \hat{S}_i^2 x_{ij}\right)^2} \right] \\ + \frac{\sum_i \hat{S}_i^2 x_{ij} \sum_i \hat{S}_i (\zeta_i - \varepsilon_i) - \sum_i \hat{S}_i^2 \sum_i x_{ij} \hat{S}_i (\zeta_i - \varepsilon_i)}{\sum_i \hat{S}_i^2 \sum_i x_{ij}^2 \hat{S}_i^2 - \left(\sum_i \hat{S}_i^2 x_{ij}\right)^2}.$$

Using the Weak Law of Large Numbers and Slutsky's Theorem as before:

$$\hat{\alpha}_2 \xrightarrow{\text{p}} \lambda_j \left[ 1 - \frac{\mathbb{E}(\hat{S}_i^2)\mathbb{E}(x_{ij}^2 \hat{S}_i \zeta_i) - \mathbb{E}(\hat{S}_i^2 x_{ij})\mathbb{E}(\hat{S}_i x_{ij} \zeta_i)}{\mathbb{E}(\hat{S}_i^2)\mathbb{E}(x_{ij}^2 \hat{S}_i^2) - [\mathbb{E}(\hat{S}_i^2 x_{ij})]^2} \right] \\ + \frac{\mathbb{E}(\hat{S}_i^2 x_{ij})\mathbb{E}(\hat{S}_i(\zeta_i - \varepsilon_i)) - \mathbb{E}(\hat{S}_i^2)\mathbb{E}(x_{ij} \hat{S}_i (\zeta_i - \varepsilon_i))}{\mathbb{E}(\hat{S}_i^2)\mathbb{E}(x_{ij}^2 \hat{S}_i^2) - [\mathbb{E}(\hat{S}_i^2 x_{ij})]^2}.$$

Since, by assumption, $x_{ij}$ is uncorrelated with $\hat{S}_i, \hat{S}_i^2, \zeta_i$, $\varepsilon_i$ and any products of these variables, and $x_{ij}^2$ is uncorrelated with $\hat{S}_i, \hat{S}_i^2$ and $\zeta_i$, the expression above simplifies to:

$$\hat{\alpha}_2 \xrightarrow{\text{p}} \lambda_j \left[ 1 - \frac{\mathbb{E}(\hat{S}_i \zeta_i)}{\mathbb{E}(\hat{S}_i^2)} \right].$$

And given that $\hat{S}_i = S_i + \zeta_i$ by construction, we can write:

$$\hat{\alpha}_2 \xrightarrow{\text{p}} \lambda_j \frac{\text{Cov}(\hat{S}_i, S_i)}{\text{Var}(\hat{S}_i)}.$$

□

#### 3 Simulation details

Finally, in this section we describe the construction of simulated traits without interactions (to assess propensity for false positives) and with interactions (to evaluate power to detect real effects).

##### 3.1 Null simulations

To build simulated traits without any interactions, we select a set of SNPs, indexed by $k = 1, \ldots, K$, attribute them effect sizes $\beta_k$ and compute phenotypes $y_i$ for each observation $i = 1, \ldots, N$, given genotypes $x_{ik}$, as follows:

$$y_i = \sum_{k=1}^{K} \beta_k x_{ik} + \varepsilon_i,$$

where $\varepsilon_i$ is a stochastic error term drawn independently for each observation from a zero-centred Normal distribution whose variance is set so as to obtain a desired level of (additive) heritability.

Since we do not want to mistake a global non-linear functional form of the genetics–trait mapping (as described in Section 2.4.4) for a 'local' interaction, we also build two additional versions of each trait in which we apply a non-linear function $f : \mathbb{R} \to \mathbb{R}$ to the additive genetic component. This will enable us to check whether this type of transformation causes our method to detect false-positive interactions. The two transformations are as follows:

$$y_i = f\left(\sum_{k=1}^{K} \beta_k x_{ik}\right) + \varepsilon_i, \quad \text{or} \tag{26}$$

$$y_i = f\left(\sum_{k=1}^{K} \beta_k x_{ik} + \varepsilon_i\right), \tag{27}$$

i.e., we either transform the additive genetic effects and then add Normal noise, or we first add noise and then apply the transformation. We expect the latter transformation not to be a challenge to our approach since this non-linearity is not affected by noise and should thus be easily reversed by normalisation (INT) of the trait. We set $f$ to be a scaled sigmoidal function with scaling factor given by the standard deviation of the additive genetic effect: denoting the observed standard deviation of $\sum_{k=1}^{K} \beta_k x_{ik}$ across observations by $\hat{\sigma}_A$, we set $f(x) = 1/[1 + \exp(-x/\hat{\sigma}_A)]$.

We make a series of simulated phenotypes by varying their genetic architecture along five axes, aiming to capture the heterogeneity seen in real phenotypes. These five dimensions are:

1. Number of causal SNPs;
2. Positioning of causal SNPs relative to one another;
3. Possible transformation of the effect sizes of these SNPs;
4. Possible global, non-linear transformation of the additive genetic effects; and
5. Level of heritability,

as we now describe.

*Number of causal SNPs:*

We choose 100, 1000 or 10 000 causal SNPs.

*Positioning of causal SNPs:*

For each possible number of causal SNPs, we either place these uniformly at random along the genome (i.e., any of the 12.7 million SNPs that survived variant filtering is equally likely to be chosen) – we term this a *'regular architecture'* – or we place half of them into clusters while the remaining half are placed uniformly at random again – we term this a *'clustered architecture'.*

More precisely, a clustered architecture is obtained as follows:

1. We first place (# causal SNPs)/(5 × 2) regions of physical length 1000 kb uniformly at random along the genome into which we will put a total of (# causal SNPs)/2 variants, resulting in an average of five causal SNPs per region;
2. We then draw the number of causal variants to place into each region from a Multinomial distribution with equal probabilities for each region and thus place half the number of causal variants into these regions;
3. Finally, we place the remaining half of the causal variants at random anywhere on the genome.

*Effect sizes and their possible transformation:*

All effect sizes are drawn independently from a standard Normal distribution. However, sampling the effect sizes of different SNPs in this manner – without considering their allele frequencies – does not take into account the expected negative relationship between effect size and frequency whereby rarer SNPs tend to have larger effects.

To make simulated architectures more realistic in this respect, it is common practice to multiply the effect sizes obtained from a standard Normal distribution by $[2p_k(1-p_k)]^{-0.5}$, a transformation that completely nullifies the positive relationship between allele frequency and heritability contribution that results from sampling effect sizes independently of frequency.[31] This was the approach adopted in the simulation setups used for testing important methods in the field such as GCTA [50], BOLT-LMM [51] or REGENIE [52].

However, a recent empirical assessment of variant-level contributions to heritability that modelled these as being proportional to $[2p_k(1-p_k)]^{1+\alpha}$ suggests that a value of $\alpha = -0.25$ provides the best fit to real phenotypes [53].[32] We therefore choose to set two possible values for $\alpha$ of 0 (in which case we do not transform the effect sizes) and -0.5 (where we multiply the effect sizes by $[2p_k(1-p_k)]^{-0.25}$) with the aim of capturing a realistic range for this parameter.

For the three axes of architectural variation discussed so far, we proceed as follows. For each combination of number of causal SNPs and their positioning, we draw a set of SNPs and corresponding effect sizes. Then, we either transform these effect sizes or not, thus obtaining for

[31] Under a fully additive architecture, the heritability contribution of a particular SNP $x_k$ with effect size $\beta_k$ is proportional to $\mathrm{Var}(\beta_k x_k) = \beta_k^2 \mathrm{Var}(x_k) = 2\beta_k^2 p_k(1-p_k)$, where $p_k$ is the MAF of $x_k$, since $\beta_k$ is fixed and $x_k$ is binomial. Multiplying $\beta_k$ by $[2p_k(1-p_k)]^{-0.5}$, the heritability contribution becomes proportional to $\mathrm{Var}(\beta_k[2p_k(1-p_k)]^{-0.5}x_k) = \beta_k^2[2p_k(1-p_k)]^{-1}2p_k(1-p_k) = \beta_k^2$, which does not depend on $p_k$.

[32] In their paper, Speed et al. [53] model heritability as depending also on the level of LD between a SNP and its neighbours and the confidence in the accuracy of its imputation, factors which we ignore in this exposition.

each number–positioning combination two vectors of effect sizes varying only in whether or not they were multiplied by $[2p_k(1-p_k)]^{-0.25}$.

*Possible non-linear transformation of the additive genetic effects:*

We then take each set of SNPs and effect sizes obtained so far and generate two 'additive genetic scores' (these will be turned into final phenotypes by adding noise as described in the next paragraph): one where we do not apply any transformation to the combined additive genetic effects, one where the transformation is applied before adding noise as in equation (26), and one where it is applied after adding noise as in equation (27). Where a transformation is applied, the transformation function $f$ is a scaled sigmoid as described above.

*Heritability:*

Heritability is set to either 30% or 60%. To achieve this, we simply add to each individual's 'total genetic score' (i.e., the total additive genetic effect with a possible sigmoidal transformation) an error term drawn from a zero-mean Normal distribution whose variance is set so that the ratio of the variance of this PGS to that of the resulting final phenotype is 30% or 60%.[33]

To summarise, we have *three* possible numbers of causal SNPs, positioned in *two* possible ways, with *two* possible sets of effect sizes,[34] with either no transformation or one of two alternative transformations (*three* possibilities) and *two* possible levels of heritability: this yields 72 null traits in total. Once generated, these simulated phenotypes are processed through exactly the same pipeline as the real ones.

##### 3.2 Simulations with interactions

Our primary goal in building simulated traits with interactions is to understand which types of interaction this method is more likely to be able to detect. To this end, we start from one of the simplest frameworks in our null simulations and extend it by adding interactions with different characteristics to its architecture.

We begin with the set of causal SNPs chosen above for phenotypes with 1000 causal loci and a regular architecture, which we consider the 'PGS' for these new simulated traits. We then choose 22 'left-hand side' (LHS) SNPs at random to interact with different subsets of 'right-hand side' (RHS) SNPs in this PGS. When evaluating the power of our method, we will assess whether these 22 focal SNPs – which differ in a number of characteristics of interest – are detected. All 22 SNPs are required to have MAF greater than or equal to 5% and to be at least 500 kb away from one another.

Having chosen these 22 LHS SNPs, we select their corresponding RHS targets. We then fix the resulting set of LHS-RHS interacting SNP pairs and create *eight different traits* varying only in whether the effect sizes of both additive effects and interactions are transformed (i.e., whether $\alpha$ is set to 0 or -0.5 as in the null simulations) and in the magnitude of the effect sizes of the interactions (for which there are four possibilities as explained below).

[33]These Normal distributions are i.i.d. across individuals.

[34]These vary only in whether or not the original $\beta_k$ were multiplied by $[2p_k(1-p_k)]^{-0.25}$ as explained above.

Interactions *within each trait* will vary along the following three dimensions:

1. Whether or not the LHS SNP has a direct effect on the trait;
2. How many RHS SNPs a LHS SNP interacts with; and
3. The sign of the interactions,

while variation *across traits* (which will share the same set of 22 LHS SNPs and corresponding RHS targets, with the same characteristics according to the previous three dimensions) will be along two axes:

4. The $\alpha$ parameter controlling the relationship between effect size and allele frequency (for both additive effects and interactions); and
5. The magnitude of the interaction effect sizes.

*Presence of a direct effect on the trait:*

An important question when analysing the interactions we detect in real data is whether or not the interacting SNPs have a main effect: do they affect the trait in an additive manner in addition to their interaction with another SNP (or SNPs) or do they only modulate the effect of other variants?

To assess the effect of these two possibilities on our ability to identify instances of epistatis, we simulate interactions that vary in whether or not the LHS SNP also has a direct effect on the trait beyond its interactions. Of the 22 LHS interacting SNPs, we choose: 8 from among the 1000 PGS SNPs that have a direct effect on the trait; 8 from among the other SNPs in the genome that do not have such an effect;[35] and, for a middle-ground scenario, 6 SNPs that, while not having a direct effect on the trait, interact primarily – at a 4-to-1 proportion as described below – with SNPs with a positive direct effect.[36] SNPs with a 'biased interaction' of this kind may be estimated to have a positive main effect even though this is in fact zero, since the least-squares estimator of a main effect in a model not containing interaction terms will partly capture such interactions if these exist.

*Number of RHS SNPs that each LHS SNP interacts with:*

We have noted before than an important motivation for our approach is the possibility that some SNPs may interact with several or even many others, and how our method will be especially powerful to capture this type of effect.

To evaluate this in practice, we vary the number of RHS SNPs that each LHS SNP interacts with. More specifically, each LHS SNP may interact with either one other SNP with an additive effect; with 2% of the SNPs with an additive effect (i.e., those in the set of 1000 causal SNPs, which we denote by PGS in this context); with 10% of these SNPs; or with the full PGS (i.e., 100% of its SNPs).

[35]These are chosen to be at least 500 kb away from any SNP with a direct effect.

[36]This is without loss of generality: an interaction primarily with SNPs with a negative effect would have a symmetric effect.

The presence of a direct effect and the number of RHS SNPs that each LHS SNP interacts with are jointly distributed as follows:

- The 8 LHS SNPs with a direct effect are divided into four groups of 2, each affecting either one SNP, or 2%, 10% or 100% of SNPs in the PGS;
- The same is true for the 8 LHS SNPs that do not have a direct effect;
- The 6 LHS SNPs that do not have a direct effect but will have a 'biased interaction' are divided into three groups of 2 SNPs, each interacting with: one other SNP, which must have a positive direct effect; 2% of the 1000 PGS SNPs (i.e., 20 SNPs), in which case 16 are chosen from among those with a positive direct effect and the remaining 4 from among those with a negative effect; or 10% of PGS SNPs (i.e., 100 SNPs), in which case 80 are chosen to have a direct positive effect and the remaining 20 a negative effect.[37]

*Sign of the interaction:*

Each LHS SNP will affect all the SNPs with which it interacts with the same sign: it is chosen to have either a positive or a negative effect (at random and with equal probability), where in the former case it increases (in absolute value) the main effect of the SNP(s) with which it interacts while in the latter case it reduces the magnitude of the effects of the RHS SNP(s).

The process just described determines the 22 LHS SNPs and their RHS pairs, as well as the sign of the interactions; these will be identical for all simulated phenotypes. We now specify how we generate eight different phenotypes that vary only in the effect sizes of these interactions.

*Effect sizes (possible transformation and magnitude):*

We will have *four traits* for each level of $\alpha$ (0 or -0.5) which arise from four different magnitudes for the interaction effect size: the interaction pairs defined above will have an effect that is 1%, 10%, 50% or 100% of the effect size *of the RHS SNP* for each trait.[38] We may therefore conceive of the interactions we simulate as rotating the increasing line connecting the additive PGS to the trait: if their sign is negative, they decrease its slope (with this change being more pronounced the greater the interaction magnitude and the proportion of PGS SNPs affected), while if it is positive they increase it.

Finally, all traits are constructed to have an *additive* heritability of 60%.

[37]Note that it is not possible to have a biased interaction effect if a LHS SNP affects 100% of the PGS SNPs as these are equally likely to have positive or negative main effects and therefore we expect there to be approximately 500 of each.

[38]For the traits whose magnitude of the interaction effect sizes is 100%, the effect size of the four SNPs that interact with 100% of PGS SNPs is reduced by 60%. This is to ensure that the additive heritability can still be set to 60%, as otherwise the heritability contribution of the interactions would be too large relative to that of the additive genetic component.

#### Supplementary tables

**Table S1:** List of 97 quantitative traits analysed. The 'UKB ID' column shows the UKB 'data-field' number; the five phenotypes that were computed manually as combinations of other phenotypes do not have such an ID. The 'N' column shows the number of samples (out of the set of 343 969 White British individuals) for which each phenotype is available. The 'Herit.' column shows the estimates of SNP heritability from ref. [2]. The 'PGS perf.' column shows the performance of each phenotype's PGS measured as the $R^2$ (coefficient of determination) of a regression of the trait (after regressing out covariates and applying INT) on this score in the held-out test set. *(continued on the next two pages)*

| Category | UKB ID | Description | N | Herit. | PGS perf. |
|---|---|---|---|---|---|
| Body size measures | 48 | Waist circumference | 343 419 | 20.6% | 8.5% |
| | 49 | Hip circumference | 343 380 | 22.3% | 10.0% |
| | – | Waist-to-hip ratio | 343 354 | – | 5.4% |
| | 50 | Standing height | 343 257 | 48.5% | 33.6% |
| | 20015 | Sitting height | 343 266 | 34.9% | 21.7% |
| | – | Sitting height, relative | 343 194 | – | 14.3% |
| | – | Leg length | 343 194 | – | 28.2% |
| | – | Leg length, relative | 343 194 | – | 14.3% |
| | 21001 | Body mass index (BMI) | 342 912 | 24.8% | 11.3% |
| | 21002 | Weight | 343 031 | 26.5% | 12.7% |
| Impedance measures | 23099 | Body fat percentage | 338 274 | 23.0% | 9.9% |
| | 23100 | Whole body fat mass | 337 940 | 23.9% | 10.5% |
| | 23101 | Whole body fat-free mass | 338 420 | 30.3% | 15.3% |
| | 23102 | Whole body water mass | 338 450 | 30.2% | 15.2% |
| | 23105 | Basal metabolic rate | 338 133 | 29.6% | 14.6% |
| | 23115 | Leg fat percentage | 338 414 | 22.1% | 9.9% |
| | 23116 | Leg fat mass | 338 411 | 23.4% | 10.0% |
| | 23117 | Leg fat-free mass | 338 401 | 28.2% | 13.2% |
| | 23118 | Leg predicted mass | 338 397 | 28.1% | 13.1% |
| | 23123 | Arm fat percentage | 338 346 | 22.5% | 9.5% |
| | 23124 | Arm fat mass | 338 317 | 23.2% | 9.6% |
| | 23125 | Arm fat-free mass | 338 308 | 27.0% | 13.0% |
| | 23126 | Arm predicted mass | 338 297 | 26.9% | 13.3% |
| | 23127 | Trunk fat percentage | 338 274 | 22.1% | 9.1% |
| | 23128 | Trunk fat mass | 338 260 | 23.9% | 10.5% |
| | 23129 | Trunk fat-free mass | 338 209 | 29.8% | 15.7% |
| | 23130 | Trunk predicted mass | 338 179 | 29.6% | 15.7% |
| Blood pressure | 102 | Pulse rate | 326 079 | 15.7% | 5.4% |
| | 4079 | Diastolic blood pressure | 326 079 | 14.3% | 4.2% |
| | 4080 | Systolic blood pressure | 326 077 | 15.1% | 4.7% |
| Urine assays | 30510 | Creatinine in urine | 334 537 | 6.6% | 0.9% |
| | 30530 | Sodium in urine | 333 854 | 7.3% | 1.1% |

| | | | | | |
|---|---|---|---|---|---|
| Blood count | 30000 | White blood cell count | 334 549 | 19.1% | 10.4% |
| | 30010 | Red blood cell count | 334 552 | 23.4% | 14.1% |
| | 30020 | Haemoglobin concentration | 334 551 | 17.4% | 9.8% |
| | 30030 | Haematocrit percentage | 334 552 | 16.3% | 8.3% |
| | 30040 | Mean corpuscular volume | 334 551 | 26.7% | 19.1% |
| | 30050 | Mean corpuscular haemoglobin | 334 548 | 25.3% | 18.2% |
| | 30060 | Mean corpuscular haemoglobin conc. | 334 546 | 5.3% | 1.1% |
| | 30070 | Red blood cell distribution width | 334 551 | 21.7% | 12.1% |
| | 30080 | Platelet count | 334 549 | 30.8% | 20.5% |
| | 30090 | Platelet crit | 334 393 | 25.6% | 16.7% |
| | 30100 | Mean platelet volume | 334 545 | 40.6% | 32.7% |
| | 30110 | Platelet distribution width | 334 392 | 20.3% | 17.8% |
| | 30120 | Lymphocyte count | 333 978 | 21.0% | 10.9% |
| | 30130 | Monocyte count | 333 978 | 23.0% | 14.4% |
| | 30140 | Neutrophil count | 333 978 | 16.4% | 7.8% |
| | 30150 | Eosinophil count | 333 978 | 18.4% | 10.6% |
| | 30180 | Lymphocyte percentage | 333 981 | 16.3% | 7.8% |
| | 30190 | Monocyte percentage | 333 981 | 21.3% | 13.0% |
| | 30200 | Neutrophil percentage | 333 981 | 15.3% | 7.0% |
| | 30210 | Eosinophil percentage | 333 981 | 21.5% | 11.1% |
| | 30220 | Basophil percentage | 333 981 | 4.9% | 1.9% |
| | 30240 | Reticulocyte percentage | 329 338 | 22.5% | 10.9% |
| | 30250 | Reticulocyte count | 329 338 | 22.7% | 11.3% |
| | 30260 | Mean reticulocyte volume | 329 339 | 20.0% | 12.8% |
| | 30270 | Mean sphered cell volume | 329 154 | 20.0% | 13.2% |
| | 30280 | Immature reticulocyte fraction | 329 154 | 16.4% | 7.9% |
| | 30290 | High light scatter reticulocyte % | 329 154 | 24.8% | 11.1% |
| | 30300 | High light scatter reticulocyte count | 329 153 | 24.8% | 11.2% |
| Blood biochemistry | 30600 | Albumin | 301 906 | 14.5% | 5.6% |
| | 30610 | Alkaline phosphatase | 328 791 | 30.9% | 17.8% |
| | 30620 | Alanine aminotransferase | 328 663 | 13.2% | 4.4% |
| | 30630 | Apolipoprotein A | 300 089 | 28.5% | 11.9% |
| | 30640 | Apolipoprotein B | 327 226 | 9.2% | 11.7% |
| | 30650 | Aspartate aminotransferase | 327 645 | 14.3% | 6.0% |
| | 30660 | Direct bilirubin | 280 815 | 43.8% | 21.0% |
| | 30670 | Urea | 328 567 | 11.9% | 4.2% |
| | 30680 | Calcium | 301 800 | 13.5% | 6.0% |
| | 30690 | Cholesterol | 328 780 | 11.2% | 5.3% |

| | | | | | |
|---|---|---|---|---|---|
| Blood biochemistry | 30700 | Creatinine | 328 611 | 21.1% | 10.7% |
| | 30710 | C-reactive protein | 328 092 | 19.3% | 6.2% |
| | 30720 | Cystatin C | 328 746 | 32.1% | 15.0% |
| | 30730 | Gamma glutamyltransferase | 328 613 | 22.8% | 7.7% |
| | 30740 | Glucose | 301 587 | 8.9% | 3.8% |
| | 30750 | Glycated haemoglobin | 328 819 | 19.6% | 12.7% |
| | 30760 | HDL cholesterol | 301 778 | 33.0% | 15.1% |
| | 30770 | IGF-1 | 327 065 | 25.3% | 11.8% |
| | 30780 | LDL direct | 328 199 | 8.2% | 6.2% |
| | 30790 | Lipoprotein(a)[39] | 262 231 | 92% | 49.6% |
| | 30810 | Phosphate | 301 343 | 13.4% | 5.4% |
| | 30830 | SHBG | 299 052 | 23.0% | 12.4% |
| | 30840 | Total bilirubin | 327 481 | 54.3% | 27.8% |
| | 30850 | Testosterone | 298 630 | 7.7% | 2.3% |
| | 30860 | Total protein | 301 577 | 16.7% | 8.1% |
| | 30870 | Triglycerides | 328 513 | 21.8% | 9.2% |
| | 30880 | Urate | 328 378 | 21.4% | 11.5% |
| | 30890 | Vitamin D | 314 652 | 10.0% | 3.3% |
| Other traits | 46 | Hand grip strength | 342 646 | 11.7% | 3.8% |
| | 78 | Heel bone mineral density T-score | 198 441 | 31.5% | 14.0% |
| | 399 | No. of incorrect matches in round | 343 969 | 5.4% | 0.7% |
| | 3143 | Ankle spacing width | 198 441 | 30.2% | 12.2% |
| | 20016 | Fluid intelligence score | 132 271 | 22.3% | 5.1% |
| | 20023 | Mean time to correctly ID matches | 341 944 | 7.8% | 2.0% |
| | 20150 | FEV1 | 260 249 | 21.3% | 8.6% |
| | 20151 | FVC | 260 249 | 23.7% | 10.6% |
| | – | FEV1/FVC ratio | 260 249 | – | 8.9% |

[39] The heritability estimate of 12.7% provided in ref. [2] is likely innacurate as estimates provided in the literature are consistently much higher. We therefore report instead a recent estimate of approximately 92% from ref. [54] (the authors' model explains 90% of heritable variance or 83% of total variance, which implies that heritable variance is approximately 92%).

**Table S2:** False positives in null simulations with $\alpha = 0$.

Each row shows data for one of 18 traits, with the first three columns containing trait information (number of causal SNPs, architecture – regular or clustered – and transformation – none, before adding noise or after adding noise). The 'GWS' columns show the number of genome-significant hits in the GWAS, the 'FPR' columns the FPR (number of genome-wide significant hits divided by the total number of SNPs tested, 12.7 million) and the 'Ind.' columns the number of independent hits. The combined FPR across all 18 traits is shown in the last row.

| SNPs | Arc. | Tr. | Heritability = 0.3 | | | Heritability = 0.6 | | |
|---|---|---|---|---|---|---|---|---|
| | | | GWS | FPR | Ind. | GWS | FPR | Ind. |
| 100 | Reg. | – | 2 | $1.58 \times 10^{-7}$ | 1 | 0 | 0 | 0 |
| 100 | Reg. | Bef. | 4 | $3.15 \times 10^{-7}$ | 2 | 0 | 0 | 0 |
| 100 | Reg. | Aft. | 0 | 0 | 0 | 0 | 0 | 0 |
| 100 | Cl. | – | 0 | 0 | 0 | 0 | 0 | 0 |
| 100 | Cl. | Bef. | 0 | 0 | 0 | 0 | 0 | 0 |
| 100 | Cl. | Aft. | 1 | $7.88 \times 10^{-8}$ | 1 | 1 | $7.88 \times 10^{-8}$ | 1 |
| 1K | Reg. | – | 0 | 0 | 0 | 1 | $7.88 \times 10^{-8}$ | 1 |
| 1K | Reg. | Bef. | 0 | 0 | 0 | 0 | 0 | 0 |
| 1K | Reg. | Aft. | 0 | 0 | 0 | 1 | $7.88 \times 10^{-8}$ | 1 |
| 1K | Cl. | – | 0 | 0 | 0 | 0 | 0 | 0 |
| 1K | Cl. | Bef. | 0 | 0 | 0 | 0 | 0 | 0 |
| 1K | Cl. | Aft. | 0 | 0 | 0 | 0 | 0 | 0 |
| 10K | Reg. | – | 0 | 0 | 0 | 1 | $7.88 \times 10^{-8}$ | 1 |
| 10K | Reg. | Bef. | 0 | 0 | 0 | 1 | $7.88 \times 10^{-8}$ | 1 |
| 10K | Reg. | Aft. | 0 | 0 | 0 | 0 | 0 | 0 |
| 10K | Cl. | – | 1 | $7.88 \times 10^{-8}$ | 1 | 0 | 0 | 0 |
| 10K | Cl. | Bef. | 1 | $7.88 \times 10^{-8}$ | 1 | 0 | 0 | 0 |
| 10K | Cl. | Aft. | 1 | $7.88 \times 10^{-8}$ | 1 | 0 | 0 | 0 |
| Combined FPR | | | | $4.38 \times 10^{-8}$ | | | $2.19 \times 10^{-8}$ | |

**Table S3:** False positives in null simulations with $\alpha = -0.5$.

| SNPs | Arc. | Tr. | Heritability = 0.3 | | | Heritability = 0.6 | | |
|---|---|---|---|---|---|---|---|---|
| | | | GWS | FPR | Ind. | GWS | FPR | Ind. |
| 100 | Reg. | – | 1 | $7.88 \times 10^{-8}$ | 1 | 0 | 0 | 0 |
| 100 | Reg. | Bef. | 2 | $1.58 \times 10^{-7}$ | 1 | 0 | 0 | 0 |
| 100 | Reg. | Aft. | 0 | 0 | 0 | 0 | 0 | 0 |
| 100 | Cl. | – | 0 | 0 | 0 | 4 | $3.15 \times 10^{-7}$ | 1 |
| 100 | Cl. | Bef. | 0 | 0 | 0 | 3 | $2.36 \times 10^{-7}$ | 2 |
| 100 | Cl. | Aft. | 0 | 0 | 0 | 0 | 0 | 0 |
| 1K | Reg. | – | 0 | 0 | 0 | 0 | 0 | 0 |
| 1K | Reg. | Bef. | 1 | $7.88 \times 10^{-8}$ | 1 | 0 | 0 | 0 |
| 1K | Reg. | Aft. | 0 | 0 | 0 | 0 | 0 | 0 |
| 1K | Cl. | – | 0 | 0 | 0 | 0 | 0 | 0 |
| 1K | Cl. | Bef. | 0 | 0 | 0 | 1 | $7.88 \times 10^{-8}$ | 1 |
| 1K | Cl. | Aft. | 0 | 0 | 0 | 1 | $7.88 \times 10^{-8}$ | 1 |
| 10K | Reg. | – | 0 | 0 | 0 | 0 | 0 | 0 |
| 10K | Reg. | Bef. | 1 | $7.88 \times 10^{-8}$ | 1 | 3 | $2.36 \times 10^{-7}$ | 2 |
| 10K | Reg. | Aft. | 1 | $7.88 \times 10^{-8}$ | 1 | 0 | 0 | 0 |
| 10K | Cl. | – | 0 | 0 | 0 | 0 | 0 | 0 |
| 10K | Cl. | Bef. | 0 | 0 | 0 | 0 | 0 | 0 |
| 10K | Cl. | Aft. | 0 | 0 | 0 | 0 | 0 | 0 |
| Combined FPR | | | | $2.63 \times 10^{-8}$ | | | $5.25 \times 10^{-8}$ | |

**Table S4:** Detection of interacting left-hand side SNPs by their characteristics in simulated traits with interactions with $\alpha = 0$.

Each row shows one of 22 left-hand side SNPs with their MAF, coefficient, sign of interaction terms, type of effect and number or proportion of SNPs in PGS affected in the first five columns. Remaining columns correspond to the effect size of the interaction terms as a fraction of the effect size of the right-hand side SNP and show log-transformed p-values with sign of corresponding coefficient obtained from SNP $\times$ PGS interaction testing. Genome-wide significant p-values are highlighted in bold.

| MAF | Coeff. | Sign | Type | # SNPs affected | *Trait 1* | *Trait 2* | *Trait 3* | *Trait 4* |
|---|---|---|---|---|---|---|---|---|
| | | | | | Effect: 1% | Effect: 10% | Effect: 50% | Effect: 100% |
| 0.38 | 0.7 | - | Direct effect | 1 | 0.2 | -0.9 | -1.7 | -2.6 |
| 0.42 | -0.1 | + | Direct effect | 1 | -0.3 | 0.1 | 0.6 | 0.9 |
| 0.28 | 0.84 | - | Direct effect | 2% | 0 | 0.5 | -0.8 | **-8.1** |
| 0.46 | 1.09 | - | Direct effect | 2% | -0.2 | -1.4 | -3.6 | **-8.2** |
| 0.13 | -1.04 | + | Direct effect | 10% | 3.7 | 4.2 | **61.2** | **216.9** |
| 0.34 | -1.86 | + | Direct effect | 10% | 0.5 | 0.9 | **34.3** | **127.5** |
| 0.12 | -0.18 | - | Direct effect | 100% | -1 | **-142.8** | **-3242.4** | **-2089.4** |
| 0.42 | -0.01 | + | Direct effect | 100% | 4.4 | **341.4** | **7425.4** | **5055.9** |
| 0.11 | 0 | - | No direct effect | 1 | -0.1 | 0.1 | -1.3 | -0.2 |
| 0.45 | 0 | + | No direct effect | 1 | 1 | -0.4 | 1 | 1.5 |
| 0.08 | 0 | - | No direct effect | 2% | -0.6 | -1.3 | -3.6 | -6.1 |
| 0.29 | 0 | - | No direct effect | 2% | -1.5 | -0.7 | -3.7 | **-22.6** |
| 0.25 | 0 | + | No direct effect | 10% | 0.1 | 1.8 | **36.5** | **150.3** |
| 0.39 | 0 | - | No direct effect | 10% | 0.5 | -2.7 | **-34.9** | **-147.2** |
| 0.08 | 0 | + | No direct effect | 100% | 3.5 | **110** | **2000.6** | **1380.4** |
| 0.29 | 0 | + | No direct effect | 100% | 3.8 | **296.2** | **5660.4** | **3869.2** |
| 0.05 | 0 | - | Biased inter. sign | 1 | 0.6 | -1.3 | -2.7 | -1.1 |
| 0.1 | 0 | - | Biased inter. sign | 1 | 0.2 | 0 | 0.1 | 0.1 |
| 0.08 | 0 | + | Biased inter. sign | 2% | 0.3 | 0.4 | 3.4 | **8.8** |
| 0.08 | 0 | + | Biased inter. sign | 2% | 0.3 | 0.5 | 0.6 | 2 |
| 0.14 | 0 | + | Biased inter. sign | 10% | 0 | 4 | **33.5** | **135.5** |
| 0.3 | 0 | + | Biased inter. sign | 10% | 0.7 | 0.9 | **22** | **128.9** |

**Table S5:** Detection of interacting left-hand side SNPs by their characteristics in simulated traits with interactions with $\alpha = -0.5$.

| MAF | Coeff. | Sign | Type | # SNPs affected | *Trait 1* | *Trait 2* | *Trait 3* | *Trait 4* |
|---|---|---|---|---|---|---|---|---|
| | | | | | Effect: 1% | Effect: 10% | Effect: 50% | Effect: 100% |
| 0.38 | 0.7 | - | Direct effect | 1 | -0.1 | 0.6 | -1.7 | -2 |
| 0.42 | -0.1 | + | Direct effect | 1 | 0.3 | 0.3 | 1 | 0.1 |
| 0.28 | 0.84 | - | Direct effect | 2% | 0.3 | -0.6 | -3.1 | **-11.3** |
| 0.46 | 1.09 | - | Direct effect | 2% | 0.1 | -0.3 | -4.6 | **-8.3** |
| 0.13 | -1.04 | + | Direct effect | 10% | 1.3 | 3.6 | **48** | **201** |
| 0.34 | -1.86 | + | Direct effect | 10% | -0.1 | 3.2 | **34** | **157.8** |
| 0.12 | -0.18 | - | Direct effect | 100% | -3.2 | **-152.8** | **-3434.4** | **-2099.1** |
| 0.42 | -0.01 | + | Direct effect | 100% | 4.9 | **351.7** | **7856.3** | **4973.7** |
| 0.11 | 0 | - | No direct effect | 1 | -0.2 | 0.4 | -0.5 | 0 |
| 0.45 | 0 | + | No direct effect | 1 | 0.2 | -0.7 | 1.5 | 1.2 |
| 0.08 | 0 | - | No direct effect | 2% | -0.1 | -0.2 | -3.8 | -7.1 |
| 0.29 | 0 | - | No direct effect | 2% | -0.4 | -1.3 | -3.3 | **-16.3** |
| 0.25 | 0 | + | No direct effect | 10% | 0.1 | 1.9 | **37.2** | **147** |
| 0.39 | 0 | - | No direct effect | 10% | -0.2 | -4.7 | **-42.1** | **-159.8** |
| 0.08 | 0 | + | No direct effect | 100% | 3.5 | **115.9** | **2094.4** | **1355** |
| 0.29 | 0 | + | No direct effect | 100% | 3.5 | **289.9** | **6050.5** | **3833.2** |
| 0.05 | 0 | - | Biased inter. sign | 1 | -0.2 | 0 | -1.1 | -0.9 |
| 0.1 | 0 | - | Biased inter. sign | 1 | -0.6 | -0.3 | 0.3 | -0.2 |
| 0.08 | 0 | + | Biased inter. sign | 2% | 0.2 | 0.6 | 2.2 | 6.6 |
| 0.08 | 0 | + | Biased inter. sign | 2% | -0.4 | -0.1 | 1.3 | 2.5 |
| 0.14 | 0 | + | Biased inter. sign | 10% | 0.4 | 0.7 | **32.3** | **149.6** |
| 0.3 | 0 | + | Biased inter. sign | 10% | -0.6 | 1.3 | **32.4** | **148.4** |

#### Supplementary figures

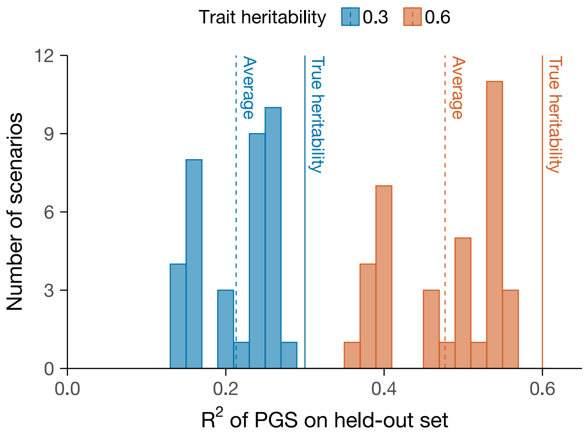

**Figure S2:** Null simulations: histogram of PGS performance (each simulation scenario is one observation) by heritability level measured by the coefficient of determination in a regression of the phenotype (from which covariates were regressed out and to which INT was subsequently applied) on the PGS in a held-out test set.

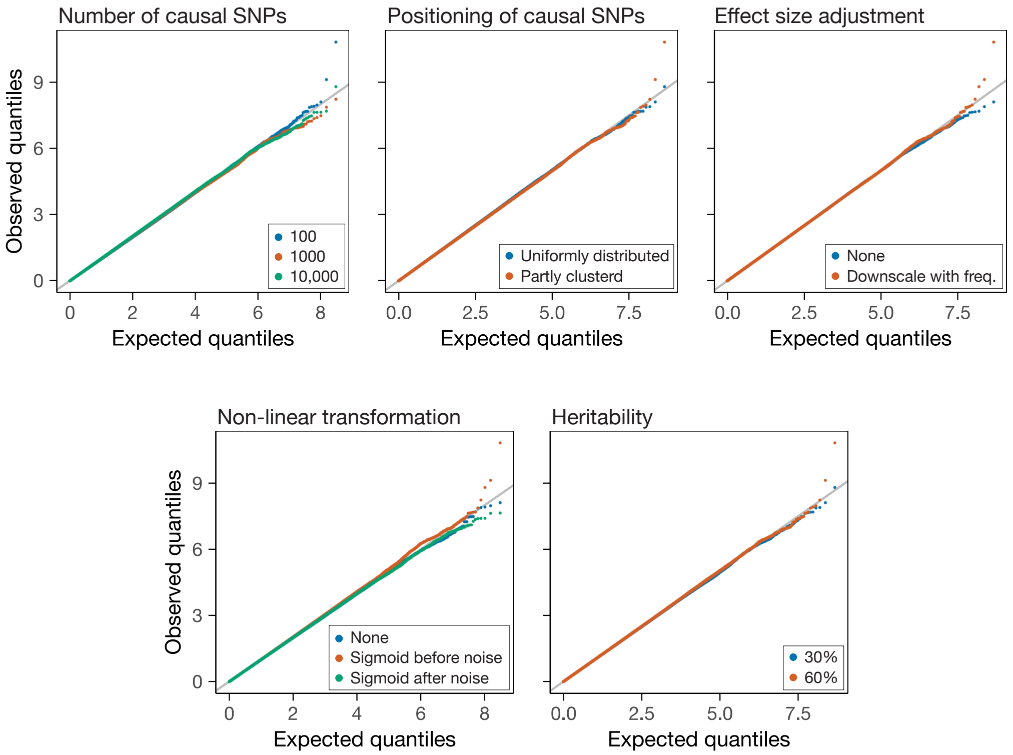

**Figure S3:** Null simulations: Q-Q plots for the SNP-by-PGS interaction term in an interaction GWAS for the different simulation parameters across the five dimensions of genetic architecure defined for these simulations.

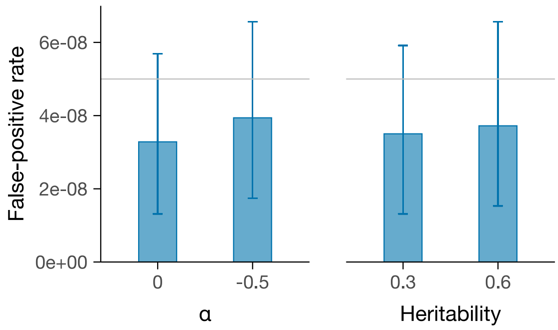

**Figure S4:** Null simulations: false-positive rates by effect size transformation and heritability parameters. The horizontal grey lines are set at $5 \times 10^{-8}$ which is the rate expected under the null hypothesis.

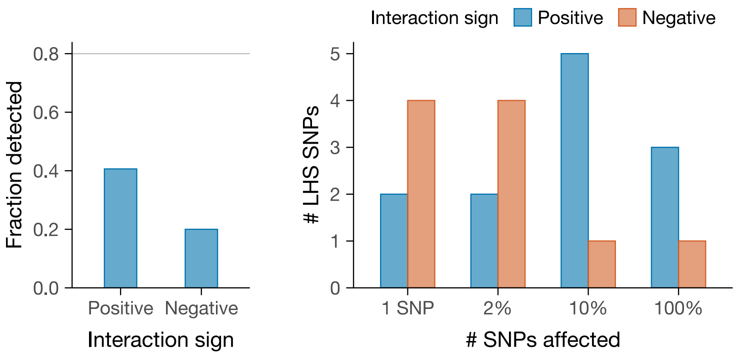

**Figure S5:** Positive simulations: fraction of interactions detected by interaction sign (left). Lower power to detect interactions with a negative sign is likely driven by the fact that, by chance, these interactions affect fewer SNPs on average (right).

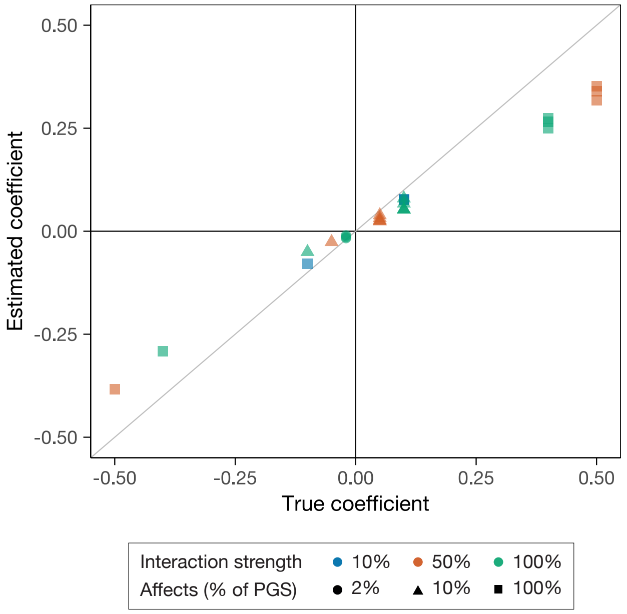

**Figure S6:** Positive simulations: accuracy of estimated interaction coefficients for phenotypes with $\alpha = -0.5$. For the 27 interactions detected, scatter plot of the estimated coefficients against their true value, with colour and shape varying by interaction strength and number of SNPs affected. The diagonal solid grey line has slope one, representing perfectly accurate estimation.

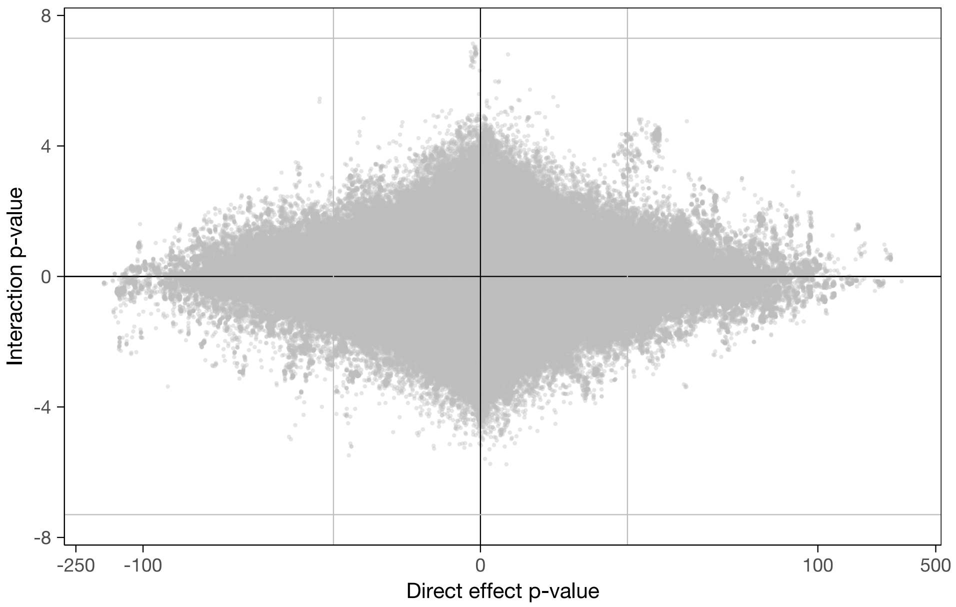

**(a)** Standing height. The interaction GWAS for this phenotype did not yield genome-wide significant associations.

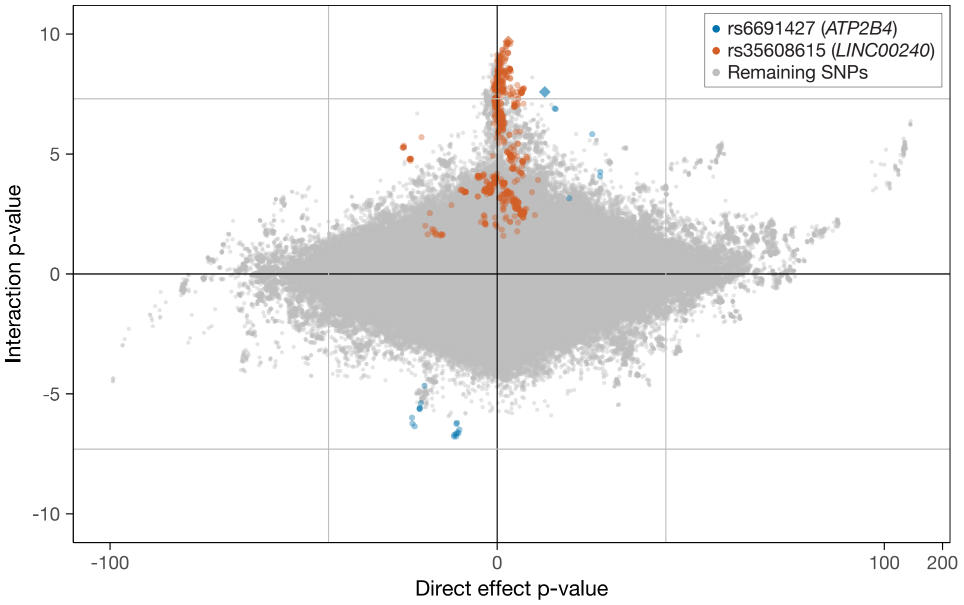

**(b)** Waist circumference.

**Figure S7:** Scatter plots of signed (by the sign of the coefficient) log-transformed p-values in the standard (vertical axis) and interaction (horizontal axis) GWASs for several phenotypes. Independent hits are plotted as diamonds coloured according to the legend. SNPs in LD (squared Pearson correlation between unphased genotypes) of 30% and higher with, and within 1 Mb of, each of the independent hits are shown as circles with the same colour as the hit they tag. The solid grey lines correspond to the genome-wide significance threshold.

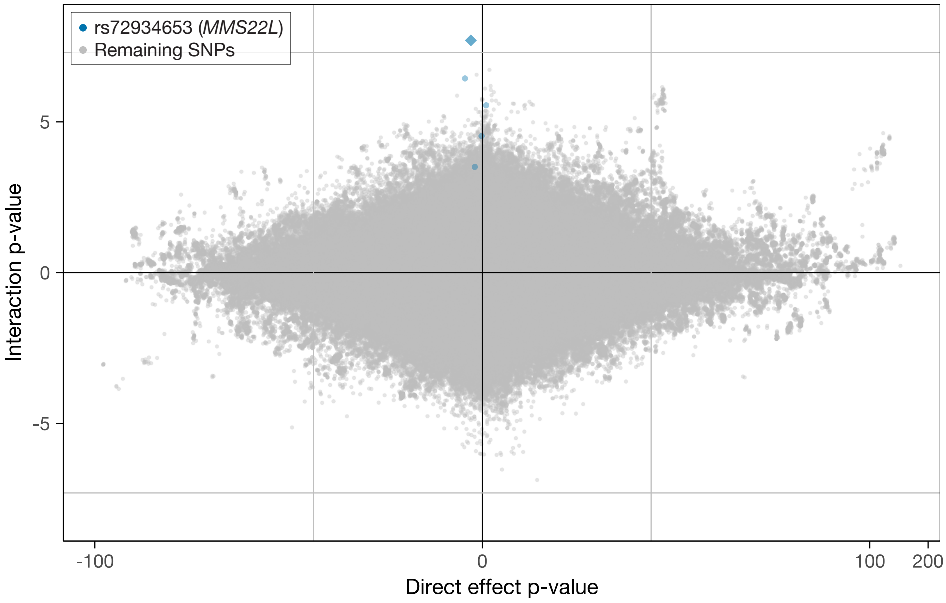

**(c)** Whole body fat-free mass.

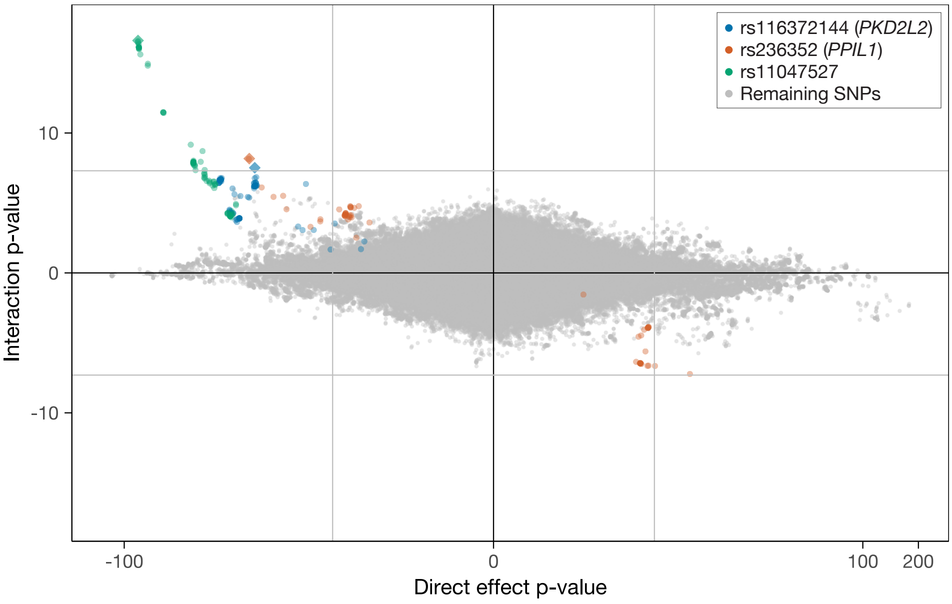

**(d)** Pulse rate.

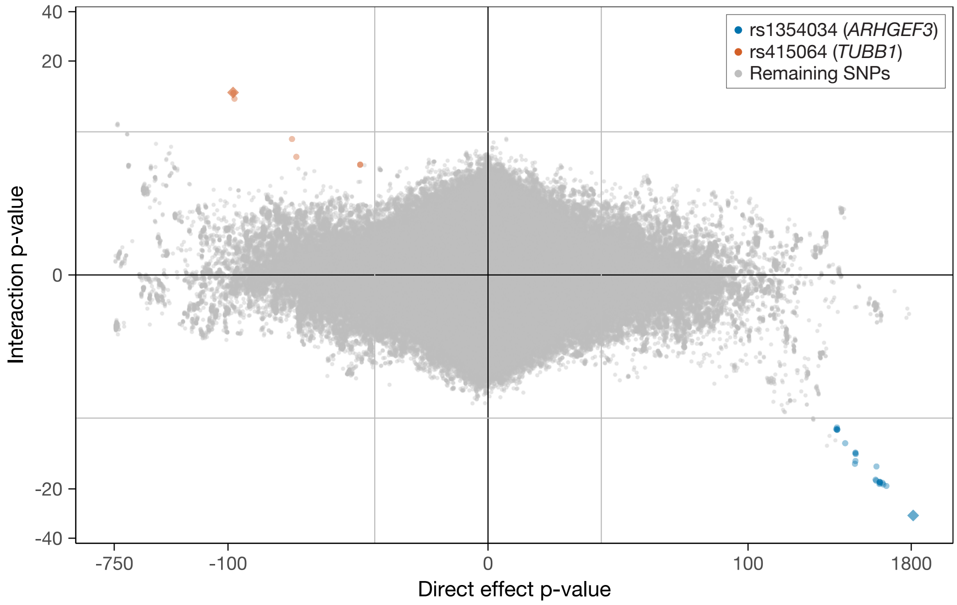

**(e)** Mean platelet volume.

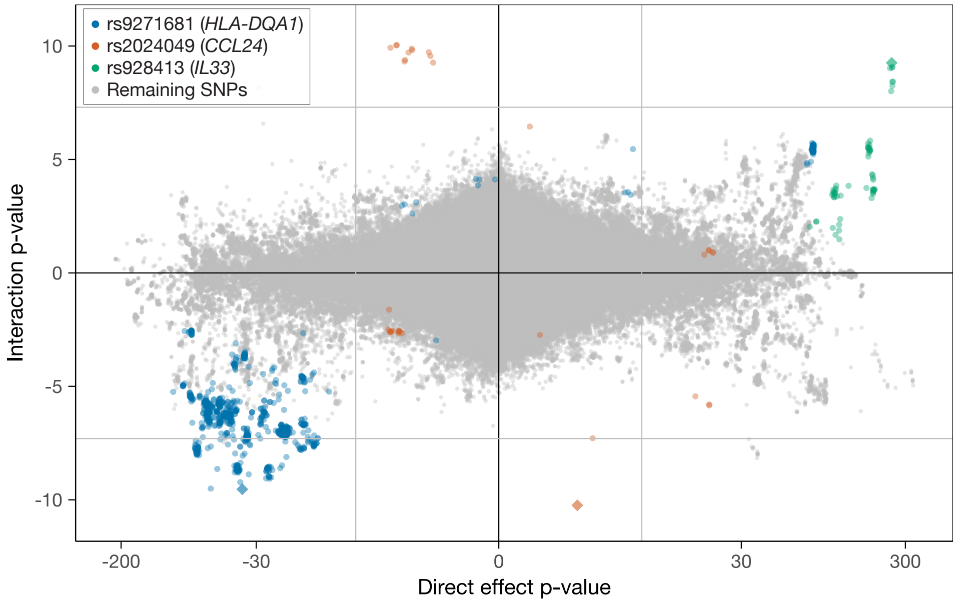

**(f)** Eosinophil count.

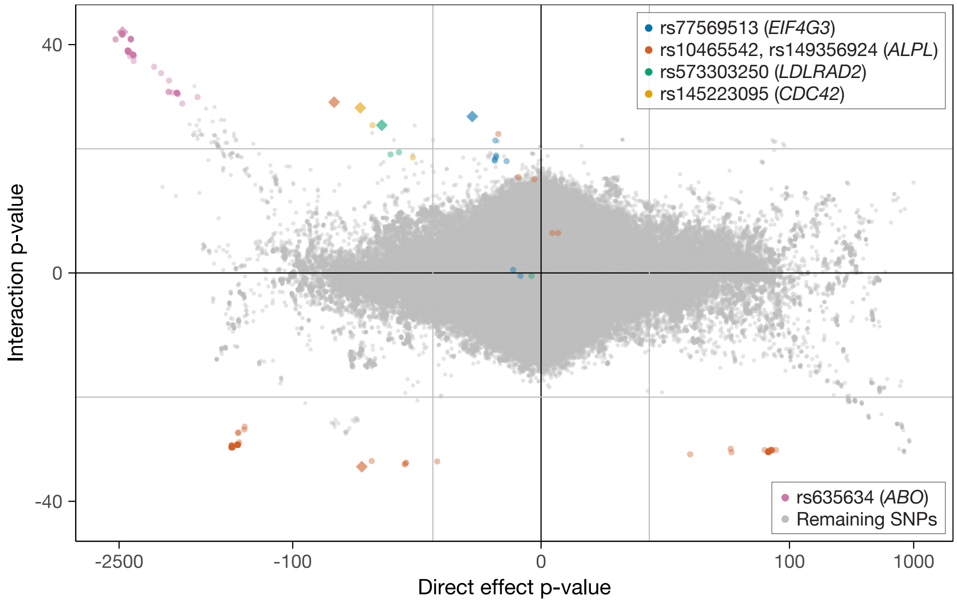

**(g)** Alkaline phosphatase.

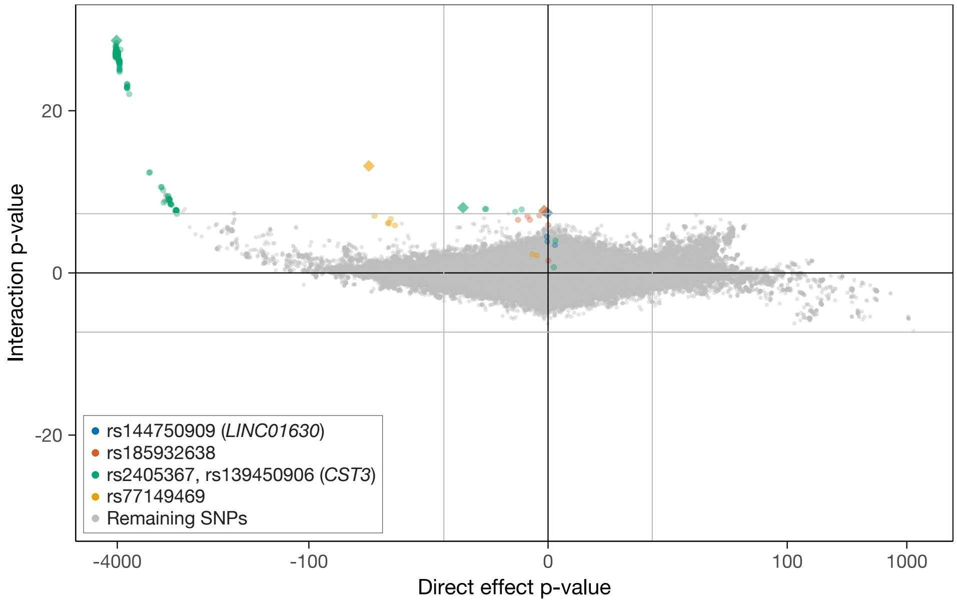

**(h)** Cystatin C.

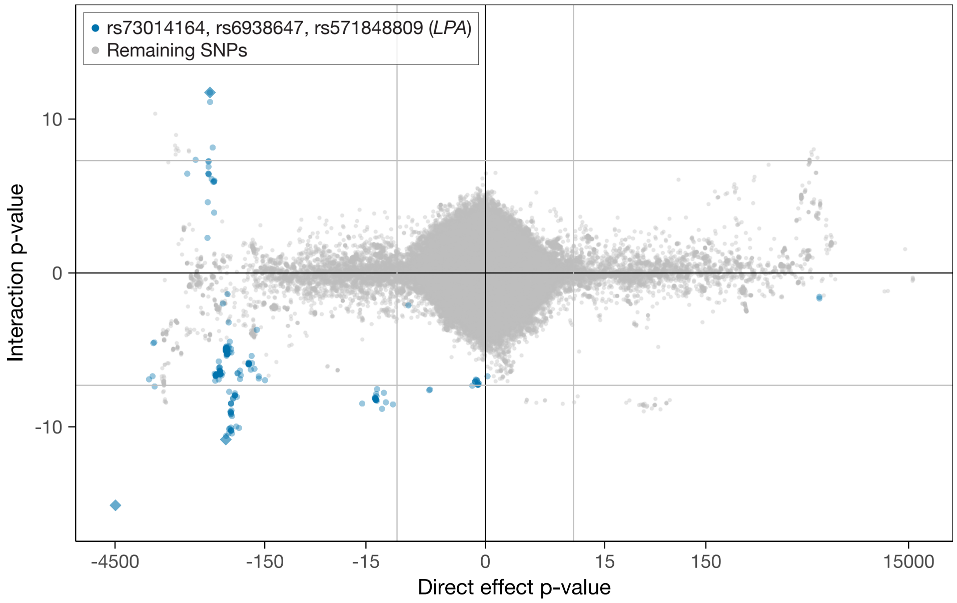

**(i)** Lipoprotein(a).

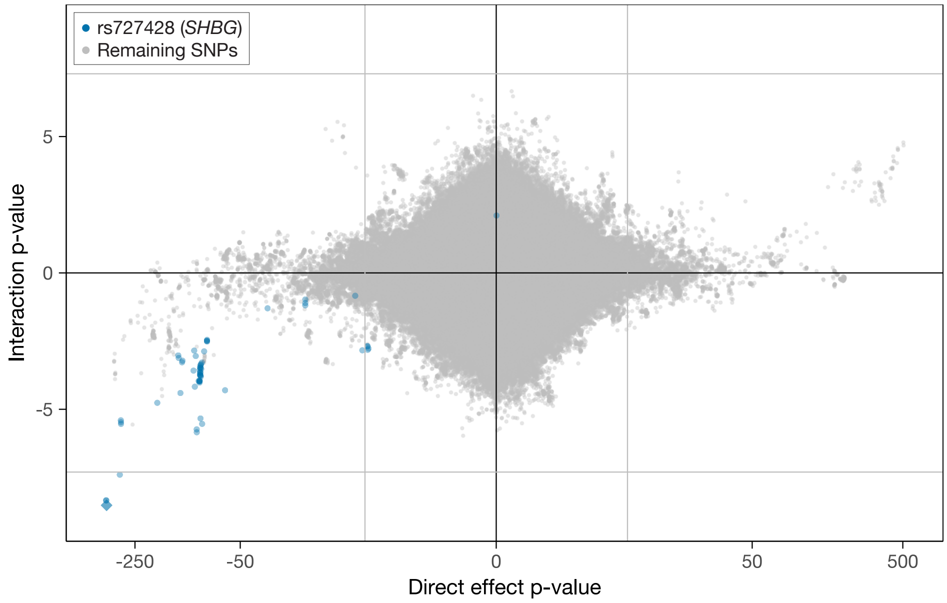

**(j)** Testosterone.

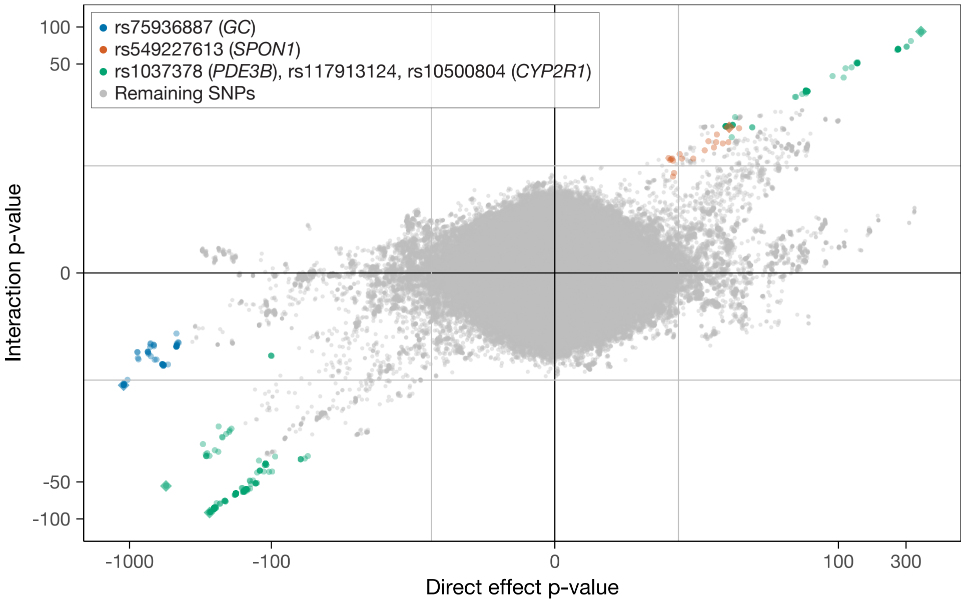

**(k)** Vitamin D.

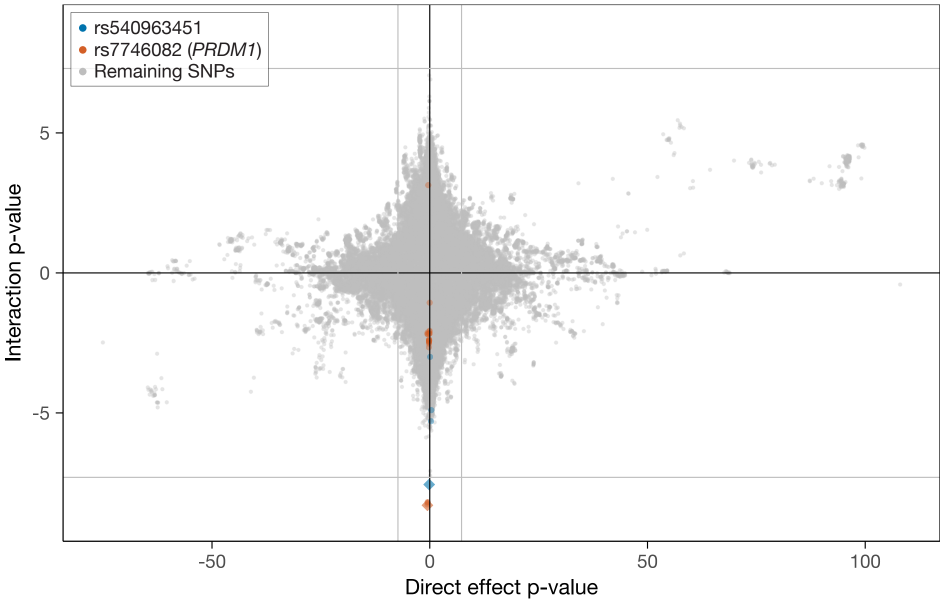

**(l)** Ankle spacing width.

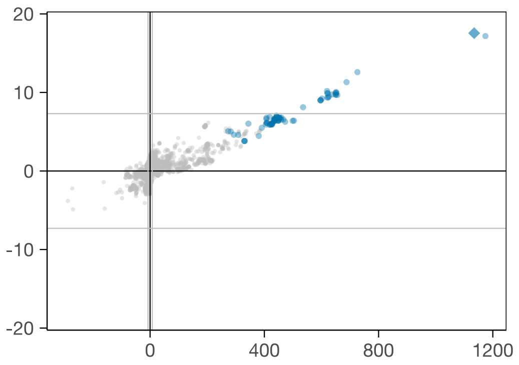

**(a)** Triglycerides: rs66505542 (*APOA5*).

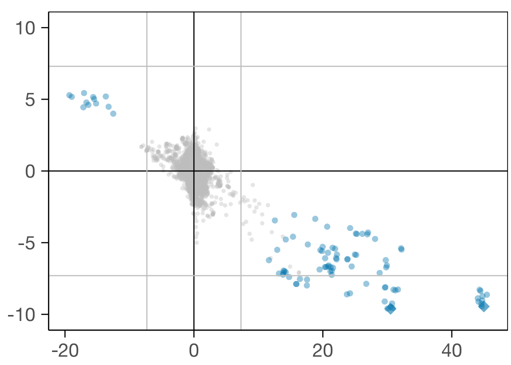

**(b)** Triglycerides: rs58719229, rs2070895 (*LIPC*).

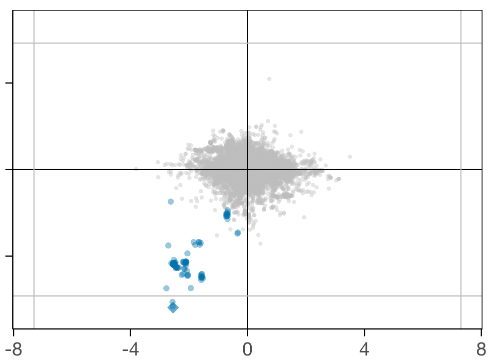

**(c)** Triglycerides: rs738408 (*PNPLA3*).

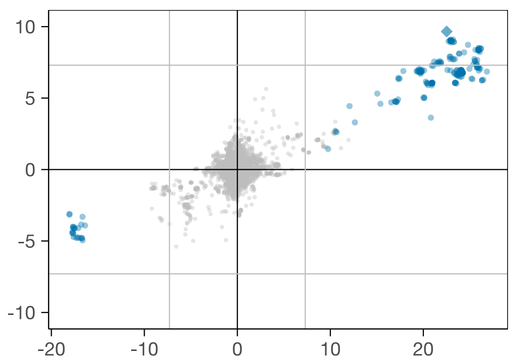

**(d)** Body fat percentage: rs34927463 (*LYPLAL1-AS1*).

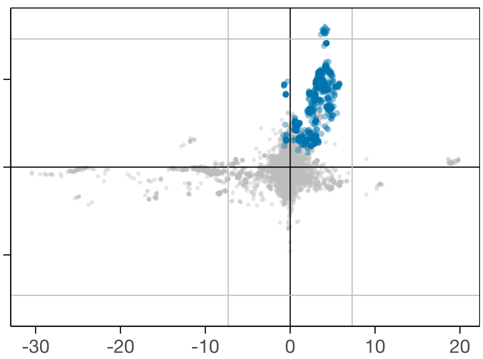

**(e)** Body fat percentage: rs35526527 (*LINC00240*).

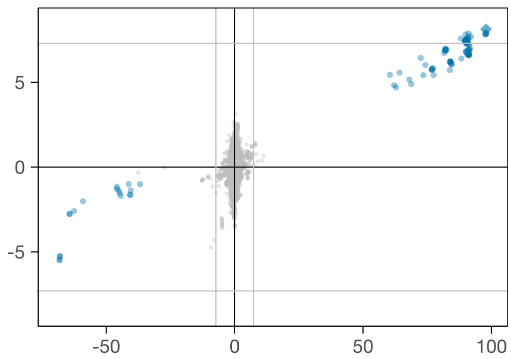

**(f)** Body fat percentage: rs56094641 (*FTO*).

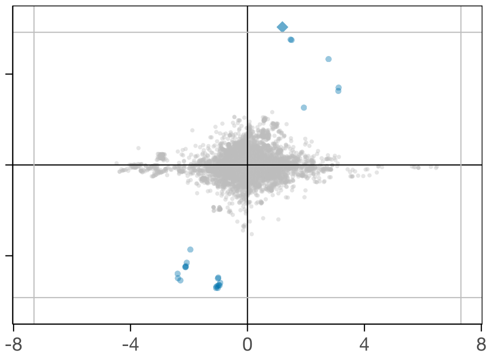

**(g)** Waist circumference: rs6691427 (*ATP2B4*).

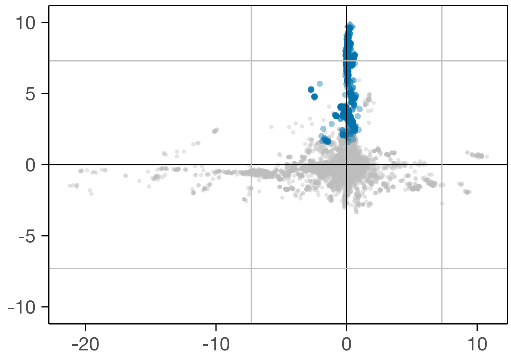

**(h)** Waist circumference: rs35608615 (*LINC00240*).

**Figure S8:** As Fig. S7 but only regions of 2 Mb around each of the loci in that figure are plotted. Independent hits are shown as diamonds; they and their tags and coloured in blue.

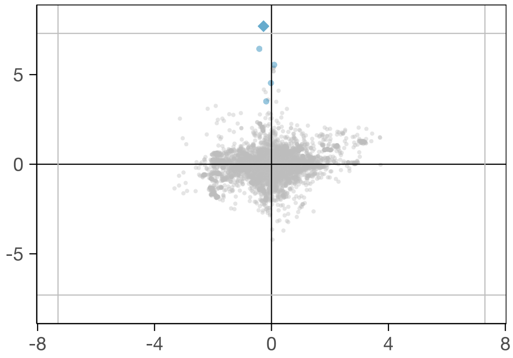

**(i)** Whole body fat-free mass: rs72934653 (*MMS22L*).

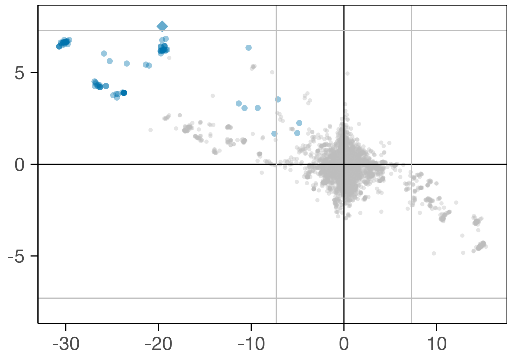

**(j)** Pulse rate: rs116372144 (*PKD2L2*).

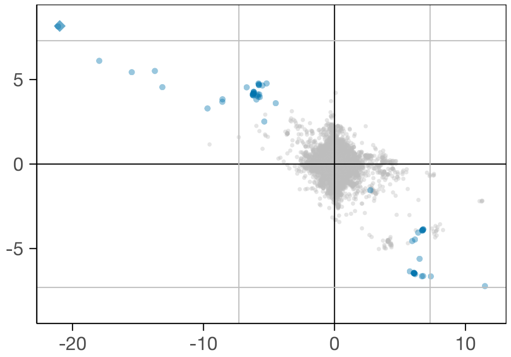

**(k)** Pulse rate: rs236352 (*PPIL1*).

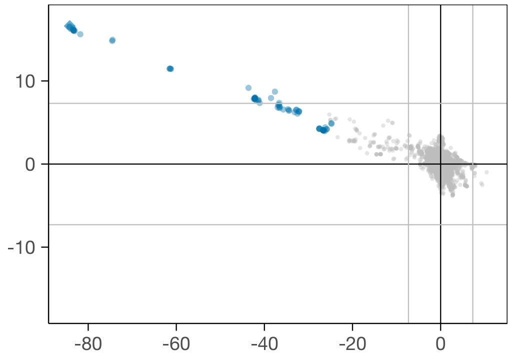

**(l)** Pulse rate: rs11047527.

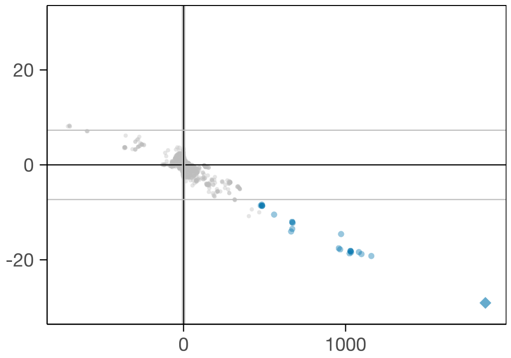

**(m)** Mean platelet volume: rs1354034 (*ARHGEF3*).

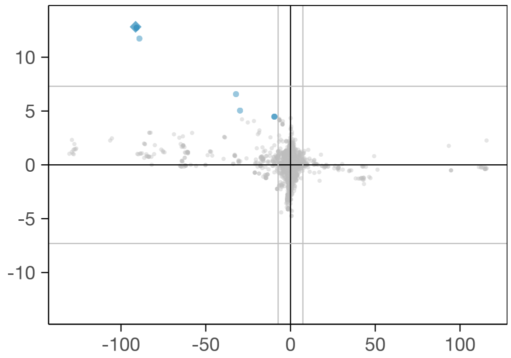

**(n)** Mean platelet volume: rs415064 (*TUBB1*).

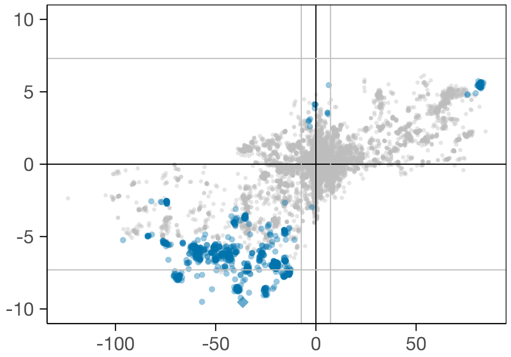

**(o)** Eosinophil count: rs9271681 (*HLA-DQA1*).

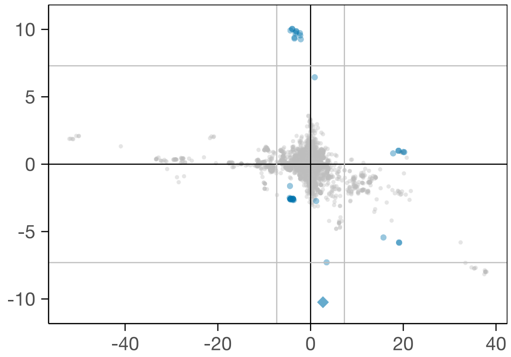

**(p)** Eosinophil count: rs2024049 (*CCL24*).

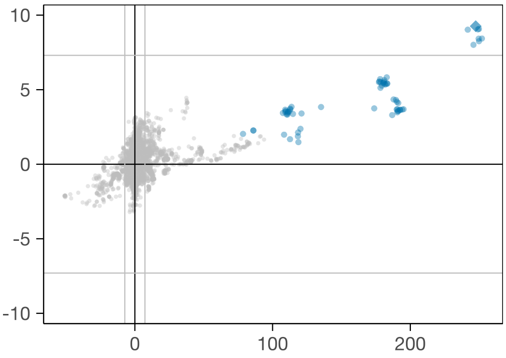

**(q)** Eosinophil count: rs928413 (*IL33*).

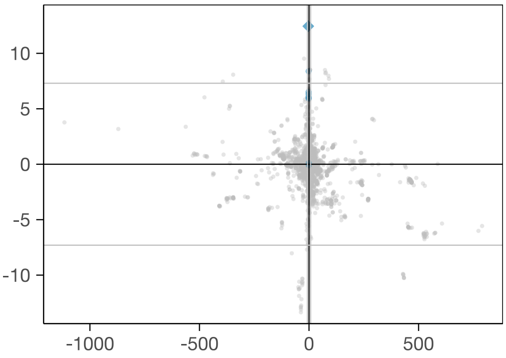

**(r)** Alkaline phosphatase: rs77569513 (*EIF4G3*).

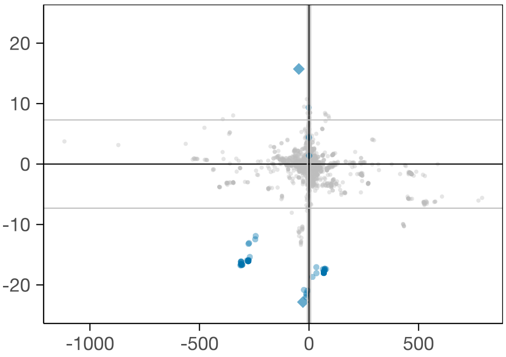

**(s)** Alkaline pho.: rs10465542, rs149356924 (*ALPL*).

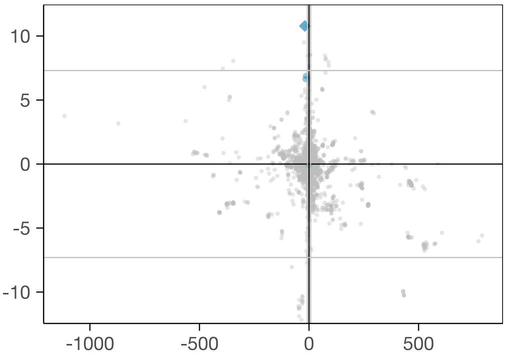

**(t)** Alkaline phosphatase: rs573303250 (*LDLRAD2*).

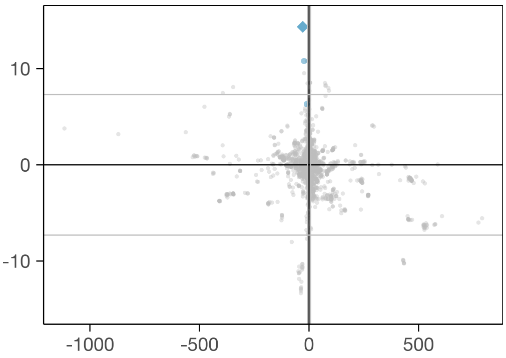

**(u)** Alkaline phosphatase: rs145223095 (*CDC42*).

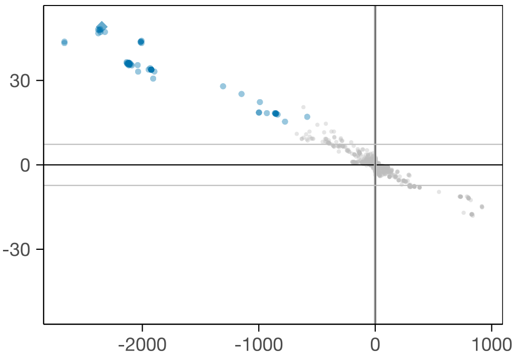

**(v)** Alkaline phosphatase: rs635634 (*ABO*).

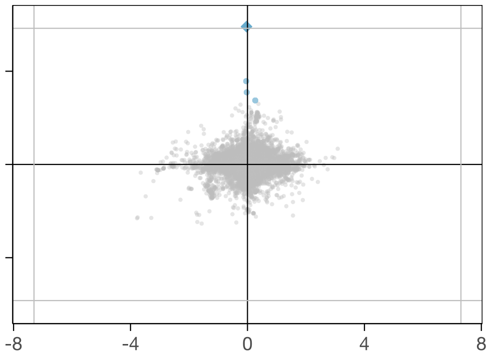

**(w)** Cystatin C: rs144750909 (*LINC01630*).

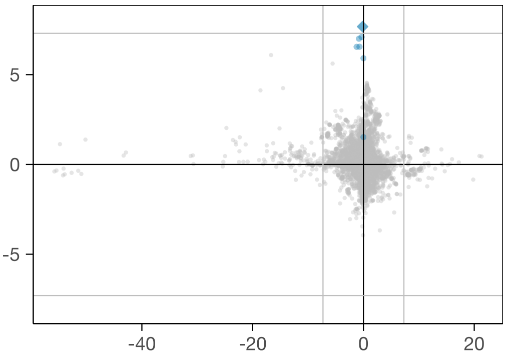

**(x)** Cystatin C: rs185932638.

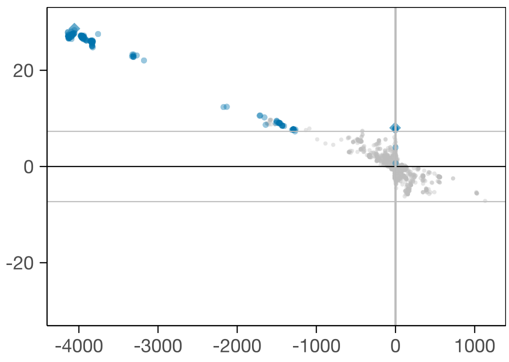

**(y)** Cystatin C: rs2405367, rs139450906 (*CST3*).

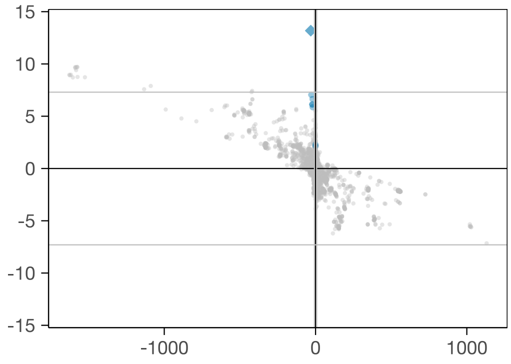

**(z)** Cystatin C: rs77149469.

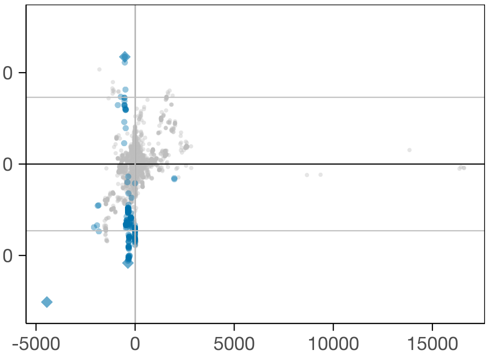

**(aa)** Lp(a): rs73014164, rs6938647, rs571848809 (*LPA*).

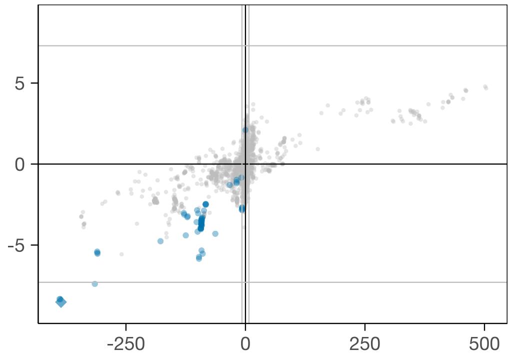

**(ab)** Testosterone: rs727428 (*SHBG*).

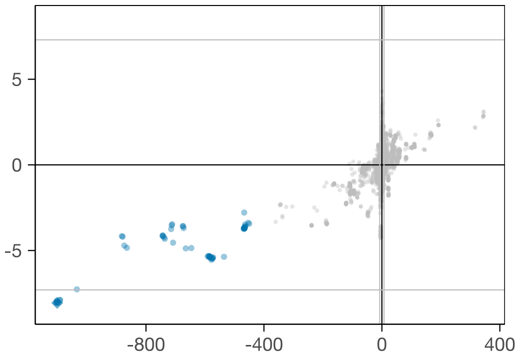

**(ac)** Vitamin D: rs75936887 (*GC*).

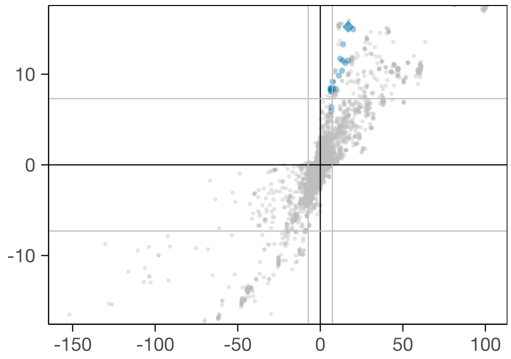

**(ad)** Vitamin D: rs549227613 (*SPON1*).

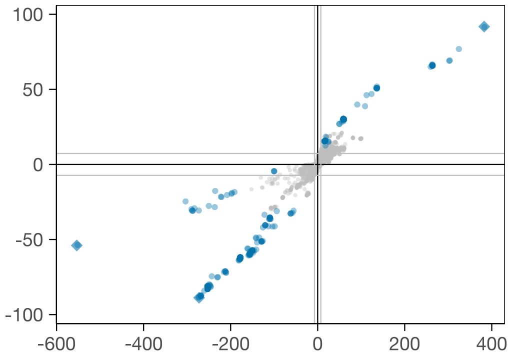

**(ae)** Vitamin D: rs1037378 (*PDE3B*), rs117913124, rs10500804 (*CYP2R1*).

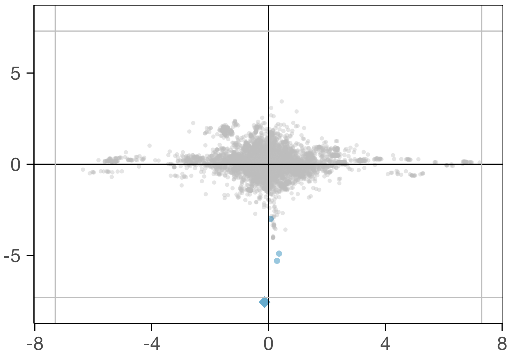

**(af)** Ankle spacing width: rs540963451.

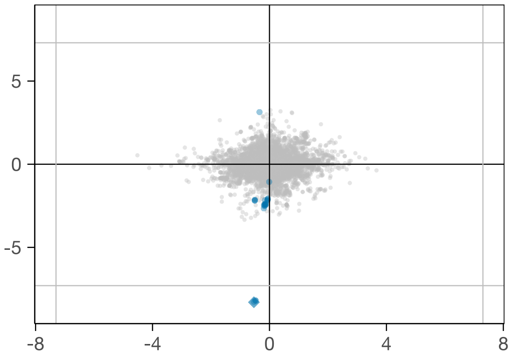

**(ag)** Ankle spacing width: rs7746082 (*PRDM1*).

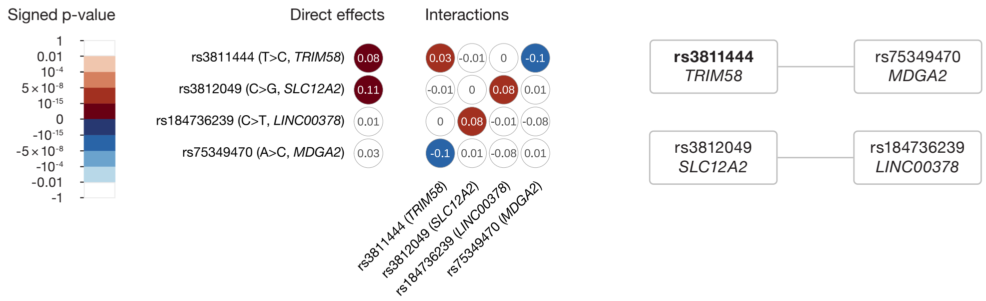

**(a)** Red blood cell distribution width.

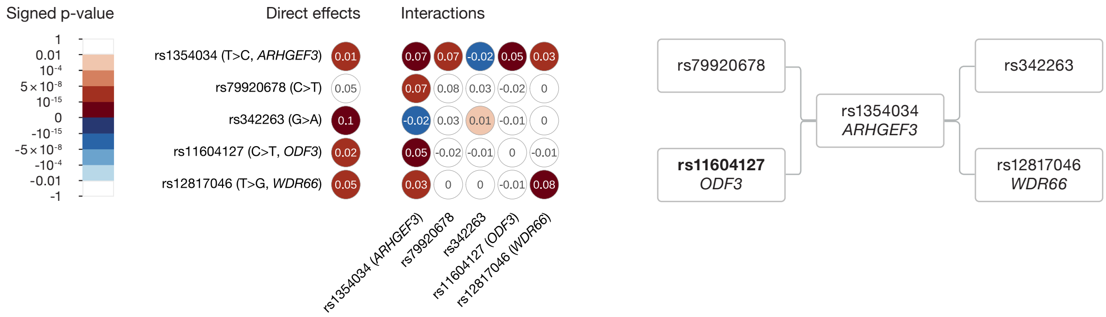

**(b)** Platelet distribution width.

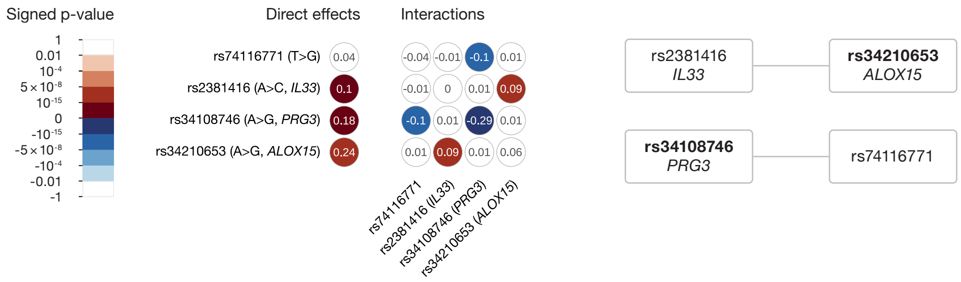

**(c)** Eosinophil percentage.

**Figure S9:** Diagrams of interaction networks for all traits with at least two independent pairwise interactions. *Left:* Coefficients (shown as numbers within each circle) and p-values (shown through colour, with sign given by coefficient) from joint linear regression model with main effects (left-most column; the effect allele was chosen so these coefficients are positive), squared allele count terms to account for possible dominance/recessiveness (diagonal elements in the matrix), and every pairwise interaction between all SNPs that are part of the trait's set of independent pairwise interactions (off-diagonal elements). *Right:* Network of pairwise interactions obtained from the regression model results. Solid connecting lines denote genome-wide significant interactions and dashed lines non-significant interactions with p-value $\leq 10^{-4}$. SNPs typed in bold are non-synonymous or tag (with $r^2 \geq 80\%$ within a 1 Mb window) a non-synonymous SNP; these are defined as SNPs with a predicted consequence at least as severe as 'protein altering variant' in the VEP database.

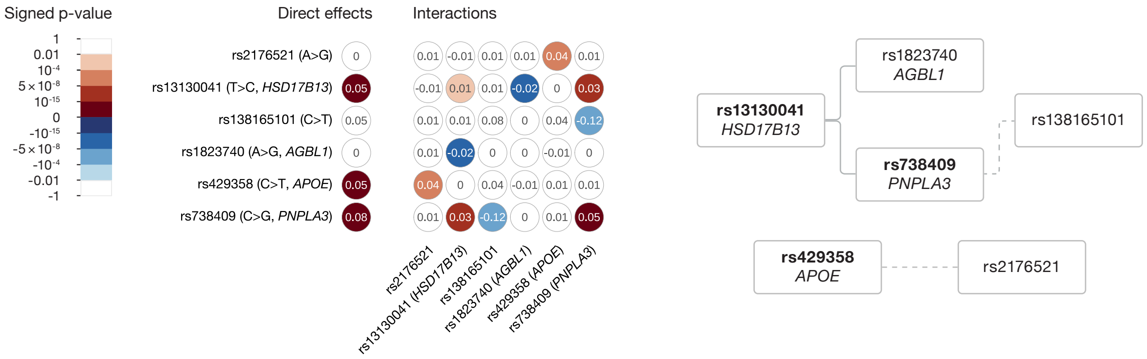

**(d)** Alanine aminotransferase. In Table S11, two neighbouring chromosome 4 variants are listed: rs13141441 (interacting with rs1823740) and rs13130041 (interacting with rs738409). These two SNPs are almost certainly a single signal (their LD is 99.5%) and so when fitting the joint regression model we include only one. We chose to include rs13130041 as it has a more severe functional consequence (intronic variant of *HSD17B13* vs. intergenic variant according to VEP). This SNP tags (with LD of 94.4%) rs72613567, a splice donor variant of the same gene.

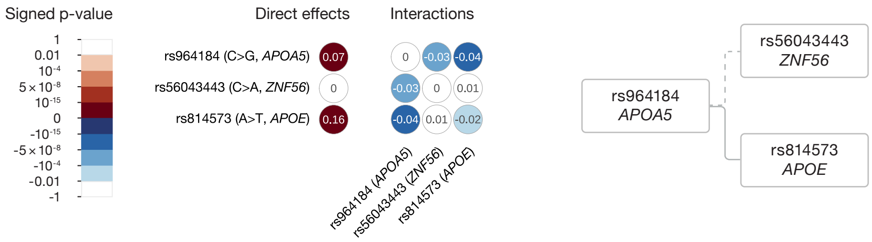

**(e)** Cholesterol. In Table S11, two neighbouring chromosome 11 variants are listed: rs66505542 (interacting with rs56043443) and rs964184 (interacting with rs814573). These two SNPs appear to represent the same signal (their LD is 90%; fitting joint regression models as in this figure with either variant in turn yields very similar results, while including both variants renders one non-significant) and so when fitting the joint regression model shown in this figure we include only one. We chose rs964184 for consistency with Fig. 4 (where it is a hit for both cholesterol and triglycerides) and as it has many more reported associations in the literature, including with cholesterol-related traits.

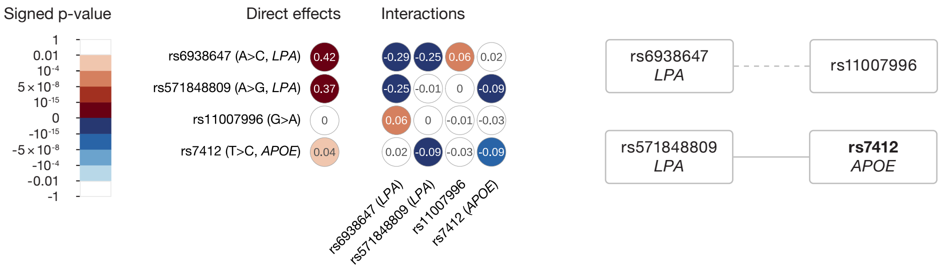

**(f)** Lipoprotein(a). In the network plot on the right, we do not show the apparent interaction between the two *LPA* variants since these are on the same chromosome and in close proximity, which raises the possibility that this putative interaction is a false positive which is in effect tagging an unobserved third variant with an additive effect (i.e., that this is an instance of 'phantom epistasis').
